# A semi-empirical Bayes approach for calibrating weak instrumental bias in sex-specific Mendelian randomization studies

**DOI:** 10.1101/2025.01.02.25319889

**Authors:** Yu-Jyun Huang, Nuzulul Kurniansyah, Daniel F. Levey, Joel Gelernter, Jennifer E. Huffman, Kelly Cho, Peter W.F. Wilson, Daniel J. Gottlieb, Kenneth M. Rice, Tamar Sofer, VA Million Veteran Program

**Affiliations:** Department of Medicine, Harvard Medical School, Boston, MA, USA; CardioVascular Institute (CVI), Beth Israel Deaconess Medical Center, Boston, MA, USA; Department of Medicine, Brigham and Women’s Hospital, Boston, MA, USA; Division of Human Genetics, Department of Psychiatry, Yale University School of Medicine, New Haven, CT, USA; Department of Psychiatry, Veterans Affairs Connecticut Healthcare Center, West Haven, CT, USA; Massachusetts Veterans Epidemiology Research and Information Center, VA Healthcare System, Boston, MA, USA; VA Palo Alto Health Care System, Palo Alto, CA, USA; Palo Alto Veterans Institute for Research, Palo Alto, CA, USA; Division of Aging, Department of Medicine, Brigham and Women’s Hospital and Harvard Medical School, Boston, MA, USA; Atlanta VA Healthcare System, Decatur, GA, USA; Department of Biostatistics, University of Washington, Seattle, WA, USA; Department of Biostatistics, Harvard T.H. Chan School of Public Health, Boston, MA, USA

## Abstract

Strong sex differences exist in sleep phenotypes and also cardiovascular diseases (CVDs). However, sex-specific causal effects of sleep phenotypes on CVD-related outcomes have not been thoroughly examined. Mendelian randomization (MR) analysis is a useful approach for estimating the causal effect of a risk factor on an outcome of interest when interventional studies are not available. We first conducted sex-specific genome-wide association studies (GWASs) for suboptimal-sleep phenotypes (insomnia, obstructive sleep apnea (OSA), short and long sleep durations, and excessive daytime sleepiness) utilizing the Million Veteran Program (MVP) dataset. We then developed a semi-empirical Bayesian framework that (i) calibrates variant-phenotype effect estimates by leveraging information across sex groups, and (ii) applies shrinkage sex-specific effect estimates in MR analysis, to alleviate weak instrumental bias when sex groups are analyzed in isolation. Simulation studies demonstrate that the causal effect estimates derived from our framework are substantially more efficient than those obtained through conventional methods. We estimated the causal effects of sleep phenotypes on CVD-related outcomes using sex-specific GWAS data from the MVP and All of Us. Significant sex differences in causal effects were observed, particularly between OSA and chronic kidney disease, as well as long sleep duration on several CVD-related outcomes. By applying shrinkage estimates for instrumental variable selection, we identified multiple sex-specific significant causal relationships between OSA and CVD-related phenotypes.

## Introduction

Investigating sex differences in health and disease mechanisms is a leading public health research priority (1–3). Sex differences are evident in various health conditions, including suboptimal-sleep phenotypes and cardiovascular diseases (CVD). For example, there is a higher prevalence of insomnia in women (4,5), whereas obstructive sleep apnea (OSA) is more common in men (6,7). Cardiovascular-related diseases, such as myocardial infarction and hypertension, generally present with a higher incidence in male adults compared to females (8– 10). Increasing numbers of genome-wide association studies (GWASs), which like other genomic studies often analyze only autosomal chromosomes, have identified strong signals of sex differences (11–13). Examples include, but are not limited to, sex differences in genetic variant effect sizes (14–16), sex-specific genetic risk associations (17–19), and sex-biased gene/protein expression level (20–22). Sex-specific causal effects of modifiable risk exposures on outcomes can, under some conditions, be obtained via Mendelian randomization (MR) analysis (23). But because of the paucity of sex-specific interventional studies, or studies with sufficient sex-stratified sample sizes, much remains unknown about how sex-specific causal effects may inform targeted disease treatments or interventions, ultimately limiting efforts to reduce sex-disparities in health (24–26).

MR analysis is widely used in genetic epidemiology because it can, using GWAS summary statistics alone (27–29), estimate causal effects from observational data. Sex-specific MR analysis, however, is limited in comparison: not only is each sex group’s sample size smaller than total in any one study, but GWAS reporting is not always sex-specific. This problem can be worse where GWAS participation is biased for structural or other reasons. For example, in the Million Veteran Program (MVP) of the U.S. Department of Veterans Affairs (VA) healthcare system, which collected genetic data and extensive phenotypes from U.S. veterans, only 10% of participants are female, in line with the proportion of female veterans. In sex-specific MR analysis, the small sample size of the female MVP population particularly limits the strength of instrumental variables (IVs) identified from female-specific GWAS, which may make causal effect estimates unstable, due to the corresponding weak IV bias (30,31).

To address these challenges, we (i) performed sex-specific GWASs of sleep phenotypes in the MVP, (ii) developed a novel statistical approach to enhance the precision of sex-specific variant-phenotype effect estimates by leveraging information across sex groups, and (iii) integrated our new shrinkage estimator into MR analyses to improve causal effect estimates, particularly for the sex group with smaller sample sizes. Motivation for this approach comes from recent findings of high correlation, between females and males, of the genetic components of multiple traits, (32,33), suggesting that many (though not all) variant associations are similar between sexes. Thus, focusing on MVP where the female population is small, our approach is to borrow information from the male population in an adaptive manner to improve female-specific variant effect estimates and, ultimately, exposure-outcome causal effect estimates.

The proposed approach, incorporating the spirit of both transfer learning and empirical Bayes, uses male-specific effect size estimates to specify prior distributions on the female-specific variant-exposure effect sizes (i.e., using information from the larger sample to improve power in the smaller). The inverse variance-weighted meta-analysis estimator and the adaptive weight estimator (proposed for analyzing secondary outcome in case-control studies (34–36)) can both be derived as the posterior mean in the proposed framework. In simulation studies, compared to standard use of variant-exposure summary statistics in MR analysis, our approach improves efficiency of exposure-outcome causal effect estimates. Finally, using sex-specific data from the MVP, along with genetic association results from the All of Us (AoU) study, we applied a two-sample MR approach to estimate the causal effects of sleep phenotypes on CVD-related outcomes. Our method identified several sex-specific causal associations. Specifically, insomnia was causally associated with an increased risk of chronic kidney disease in females, long sleep was linked to a higher risk of hypertension in females, and short sleep was associated with an increased risk of coronary artery disease in males. A statistically significant sex difference in the causal effect of OSA on chronic kidney disease was also identified. In addition, using shrinkage estimates for IV selection, we detected several statistically significant causal effects of OSA on CVD-related outcomes, as well as distinct sex differences in the causal patterns of long sleep on CVD-related outcomes, with higher risks observed in female populations

## Results

### Overview of semi-empirical Bayesian method for calibrating genetic variant effect size estimates utilizing information across groups

While our method is general, we focus on the need to improve the estimation of variant effect sizes in the relatively small MVP female population, and do so by borrowing information from the male population. Another simplification we make in the exposition is to focus on the “exposure” GWAS, even though the same framework can be applied to any trait GWAS, regardless of its role in an MR analysis. Throughout this paper, we use *γ* to represent “variant-exposure” effect size and Γ to represent “variant-outcome” effect size. To motivate our method, we consider a Bayesian prior on *γ*_*jF*_, the female-specific effect size of the *j*th SNP on the sleep phenotype, specifically

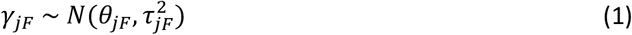

where *θ*_*jF*_,*τ*_*jF*_ are the prior mean and standard deviation (SD). The approximate distribution of the “raw” female-specific effect size estimate (i.e., an estimate that relies on female data only) is given by

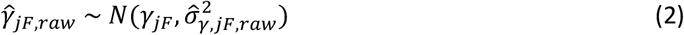

where 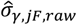 is the estimated standard error of 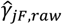.The normality assumption here is appropriate due to large GWAS sample sizes (30,31), regardless of the specific method used for estimation (maximum likelihood, method of moments, etc.).

### Potential specification of the prior distribution of female-specific SNP effect sizes and resulting posterior estimates

An intuitive way to borrow information from the male to the female population is to specify the prior mean and variance 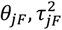 in equation (1) as the male-specific effect size estimate and its estimated variance. Formally the prior is

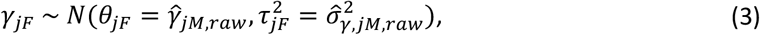

leading to posterior

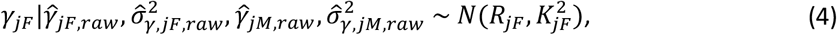

where the posterior mean 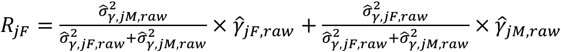 and the posterior variance 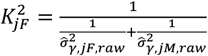. The posterior mean is a weighted average of the prior mean and sample mean, with the weight on the group-specific estimates being proportional to their precisions (i.e., the inverse of their variances). The posterior mean and variance are exactly identical to those from conventional fixed-effects inverse-variance meta-analysis of the sex-specific estimates (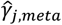,FE meta estimate henceforth). A more detailed illustration of the Bayesian approach to meta-analysis is given by Dominguez and Rice (37). The FE meta estimate has been shown to be as efficient as pooling individual level data when effects are identical across combined studies (38), so in that setting there is no penalty to using meta-analysis over any standard competing method.

However, the estimated strength of sex differences, i.e., 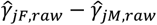, is not considered in the FE meta estimate, or standard competing methods, making their use unappealing when there are strong sex differences. To incorporate information on sex differences we consider the prior with:

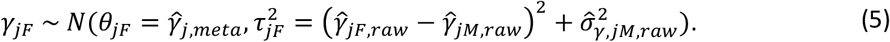

Here, the prior mean is the efficient FE meta estimate 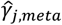,while the sex differences strength is incorporated in the prior variance. The posterior distribution of *γ*_*jF*_ is Normal, with mean 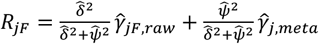, where 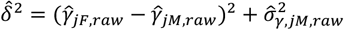, is related to both the difference in effect size estimate *and* the variance of male-specific estimate. Here, 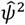 is 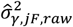.The posterior variance can be written as 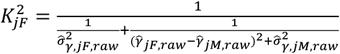.

Because the posterior mean adapts 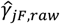 to the observed differences in sex-specific estimates, we call it the *adaptive posterior mean* (APM) estimator 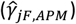.From equation (5), the prior mean and variance depend not only on sex-specific groups but also on sex-combined effect estimates and sex differences. This is why we refer to our framework as a “semi-empirical” Bayes approach.

The APM estimator is related to the adaptive weight (AW) estimator, initially proposed for gene-environment interactions or gene-secondary outcome associations in case-control studies (34– 36). In both of those contexts, the AW estimator was developed to provide a population-level estimate (i.e., not specific to either cases or controls) by adaptively combining information from both groups using weighting. In our approach, we instead use shrinkage estimators to obtain group-specific estimates. The APM estimator also differs from the original AW in its weighting parameters. APM incorporates the variance of group-specific effect estimates into its prior variance for *γ*_*jF*_, to avoid underestimating posterior variance due to smaller estimated sex differences (i.e.,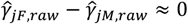). In other words, if 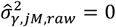 in equation (5), then the APM estimator may reduce to AW estimator. Table 1 summarizes the derivation of the FE meta and APM estimates under the Bayesian Normal-Normal modeling scheme. As stated earlier, we focus on the calibration of the female effects 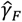,but the framework is general and can be similarly applied to the male population.

**Table 1:** The proposed Bayesian framework for calibrating variant-trait effect size estimates.

| Likelihood | Prior distribution of $\gamma_{jF}$ | Posterior mean of $\gamma_{jF}$ | Estimator name |
| --- | --- | --- | --- |
| $\hat{\gamma}_{jF,raw} \sim N(\gamma_{jF}, \hat{\sigma}_{\gamma,jF,raw}^2)$ | $\gamma_{jF} \sim N(\hat{\gamma}_{jM,raw}, \hat{\sigma}_{\gamma,jM,raw}^2)$ | $\frac{\hat{\sigma}_{\gamma,jM,raw}^2}{\hat{\sigma}_{\gamma,jF,raw}^2 + \hat{\sigma}_{\gamma,jM,raw}^2} \hat{\gamma}_{jF,raw} + \frac{\hat{\sigma}_{\gamma,jF,raw}^2}{\hat{\sigma}_{\gamma,jF,raw}^2 + \hat{\sigma}_{\gamma,jM,raw}^2} \hat{\gamma}_{jM,raw}$ | FE meta estimate<br>$\hat{\gamma}_{j,meta}$ |
| | $\gamma_{jF} \sim N(\hat{\gamma}_{j,meta}, (\hat{\gamma}_{jF,raw} - \hat{\gamma}_{jM,raw})^2 + \hat{\sigma}_{\gamma,jM,raw}^2)$ | $\frac{(\hat{\gamma}_{jF,raw} - \hat{\gamma}_{jM,raw})^2 + \hat{\sigma}_{\gamma,jM,raw}^2}{(\hat{\gamma}_{jF,raw} - \hat{\gamma}_{jM,raw})^2 + \hat{\sigma}_{\gamma,jM,raw}^2 + \hat{\sigma}_{\gamma,jF,raw}^2} \hat{\gamma}_{jF,raw} + \frac{\hat{\sigma}_{\gamma,jF,raw}^2}{(\hat{\gamma}_{jF,raw} - \hat{\gamma}_{jM,raw})^2 + \hat{\sigma}_{\gamma,jM,raw}^2 + \hat{\sigma}_{\gamma,jF,raw}^2} \hat{\gamma}_{j,meta}$ | APM estimate<br>$\hat{\gamma}_{jF,APM}$ |

The table summarizes the semi-empirical Bayesian model parameters behind the two proposed variant-trait effect size estimators. The “raw” effect size estimates are obtained from an analysis of a single stratum (here we focus on the female stratum). By specifying prior distributions on the variant-trait effect sizes (second column), the raw estimates are shrunk toward the prior means to become the estimates provided in the posterior mean column. The last column, estimator name, provides the name and notation of the resulting estimators (FE meta estimate and adaptive posterior mean (APM) estimate).

### Exposure-outcome causal effect estimation

Two-sample MR approaches estimate causal effects from two independent sets of summary statistics. These describe variant associations with an exposure and with the outcome phenotype, where the variants are selected to be valid IVs for the exposure of interest (30,39– 42). A causal effect *β*_*j*_ of exposure on outcome can then be estimated using estimated associations of variant *j* with exposure 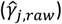 and outcome 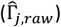 via the Wald ratio estimate 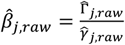. Using multiple IVs, the causal effect *β* can then be estimated by aggregating estimates from all valid IVs through various weighting approaches. Here, we assume all *p* variants are valid IVs. Our proposed Bayesian framework, therefore, is conceptualized as a preliminary step before applying MR methods. More specifically, we first calibrate the 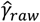 estimates using the proposed Bayesian models. Then, the shrinkage estimates, i.e., the posterior means and SDs in equation (4), are used as inputs in existing MR algorithms. The causal effect is then estimated using the newly-estimated 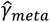 and 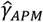 (with corresponding posterior SD) coupled with the (raw) variant-outcome estimated effect sizes. A schematic overview of the sex-specific MR analysis and the proposed Bayesian framework is illustrated in Figure 1.

**Figure 1:**
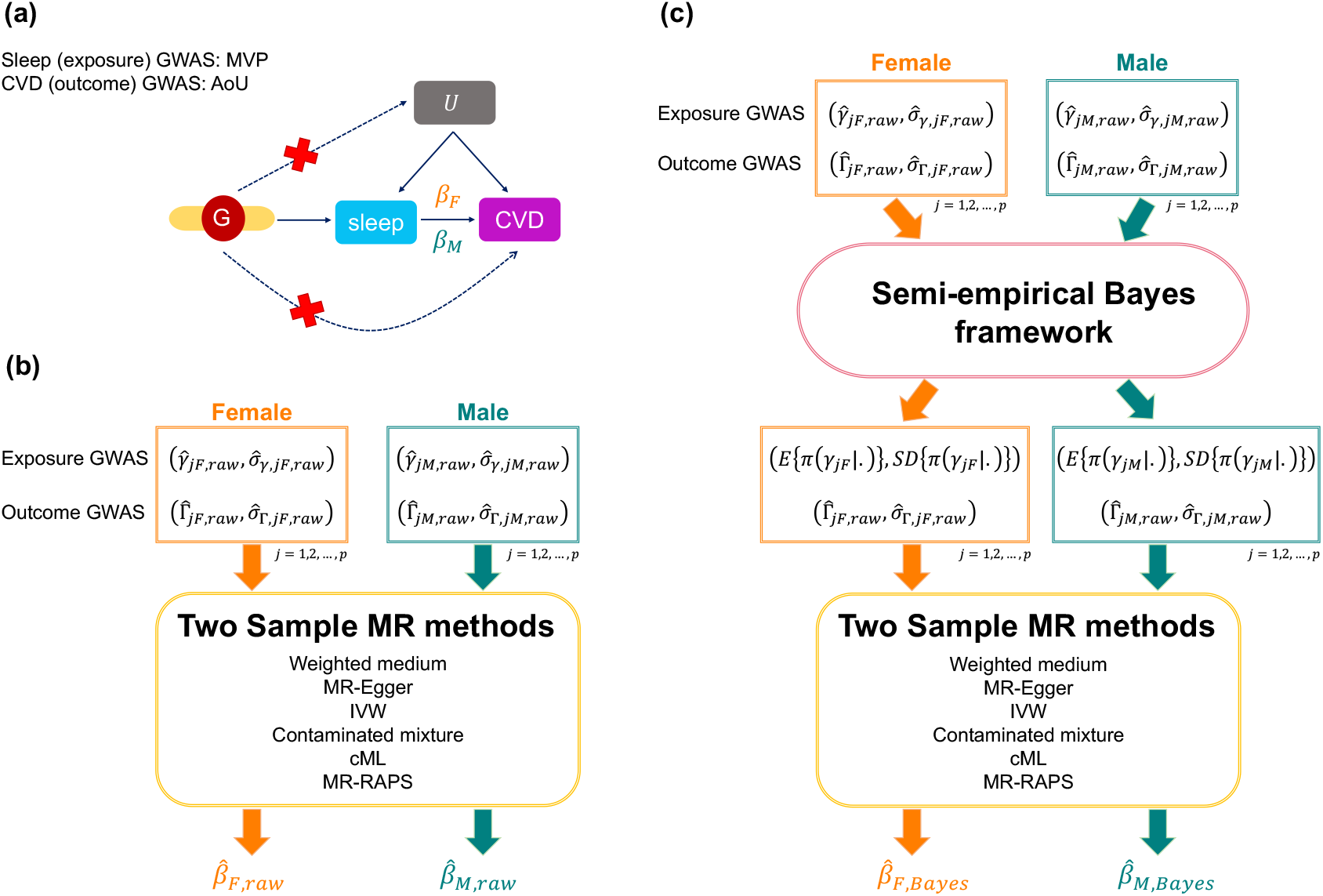
Schematic overview of the sex-specific MR analysis Panel a: A causal diagram underlying the MR framework in this manuscript. We consider sleep-related phenotypes to be the exposure factors and cardiovascular-related measures as outcome variables. *β*_*F*_ denotes the underlying female-specific exposure-outcome causal effect; *β*_*M*_ represents the underlying male-specific exposure-outcome causal effect. In our analysis, *β*_*F*_ and *β*_*M*_ are estimated separately. The GWAS summary statistics for sleep phenotypes were derived from the MVP dataset, and the summary statistics for cardiovascular-related diseases were computed in AoU; Panel b: Estimation of sex-specific causal effects using the two-sample MR approaches. The inputs for the two-sample MR methods are raw sex-specific exposure (female: 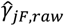 and 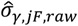;male: 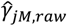 and 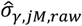) and outcome GWAS summary statistics (female: 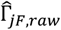 and 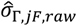;male: 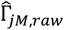;and 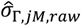) from independent samples. The outputs are: estimated female-specific causal effect 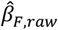 and estimated male-specific causal effect 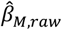;Panel c: The proposed Bayesian method. We first calibrate the raw sex-specific exposure effects by borrowing information from one sex group to the other, or across both sex groups. Two-sample MR analysis then uses the shrinkage exposure summary statistics, i.e., the posterior mean and posterior SD of *γ* (female: *E*{*π*(*γ*_*jF*_|.)} and *SD*{*π*(*γ*_*jF*_|.)}; male: *E*{*π*(*γ*_*jM*_|.)} and *SD*{*π*(*γ*_*jM*_|.)}), with the raw outcome summary statistics to provide a more robust basis for causal effect estimation. We use *π*(*γ*|.) to denote the posterior distribution of *γ*. The outputs are the estimated female-specific causal effect 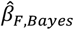,and estimated male-specific causal effect 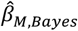,both use shrinkage exposure effect estimates in their construction. Abbreviations. MR: Mendelian randomization; F: female; M: male; CVD: cardiovascular disease; GWAS: genome-wide association study; MVP: million veteran program; AoU: All of Us; SD: standard deviation; cML: constrained maximum likelihood.

### Simulation studies

We used simulation primarily to evaluate and compare the performance of exposure-outcome causal effect (*β*) estimation using raw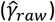 and shrinkage variant-exposure effect (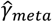and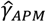) with respective SD estimates, combined with each selected MR method, particularly focusing on the population with a smaller sample size (female population in our analysis). For our Bayesian methods, the posterior SDs of *γ* are treated as standard error estimates. We also incorporated the AW estimator 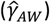 as another approach for calibrating female-specific variant-exposure effect size estimates. The two-sample MR methods considered in the analyses are summarized in Table 4 (Method section).

To mimic the structure of the MVP dataset, we generated 2,000 female individuals and 20,000 male individuals for the exposure GWAS, maintaining a similar proportion of females to males as in the MVP. For the outcome GWAS, we generated balanced datasets of 10,000 individuals for both female and male populations. We generated 100 independent SNPs as IVs in all simulations, with all allele frequencies set at 0.3. We use *D*_*jγ*_ = *γ*_*jF*_ − *γ*_*jM*_, *j* = 1,2, …, *p* to denote the sex differences strength in variants associations with the exposure, *γ*. We write *D*_*γ*_ = {*j*|*D*_*jγ*_ = *γ*_*jF*_ − *γ*_*jM*_ ≠ 0} and | *D*_*γ*_ | to represent the set of variants and the number of variants having sex-differences in variant-exposure effect size, respectively. We considered three simulation scenarios: (i) fixed *D*_*jγ*_ = 0.05 if variant *j* ∈ *D*_*γ*_ (ii) random *D*_*jγ*_, and (iii) using MVP OSA GWAS summary statistics to guide the simulated differences *D*_*jγ*_, which consists of strong *D*_*jγ*_ patterns with weak IVs in the female population (average *F*-statistic < 10). In simulation (ii) and (iii), all variants *j* ∈ *D*_*γ*_ (i.e., | *D*_*γ*_ | = 100). Within each simulation, we also considered a few levels of sex differences in “causal effect” settings (i.e., *β*_*F*_ ≠ *β*_*M*_). A more detailed description of the simulation studies is provided in Method section, Supplementary Note 1, Supplementary Tables 1 and 2.

We summarized the estimation performance of the 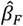 and 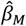 in the simulation studies using two metrics: (i) the mean squared error (MSE) of the estimated effect and (ii) the 95% confidence interval’s actual coverage of the true effect, where these 95% confidence intervals for *β* under each MR method were computed using standard asymptotic normality properties. The female results from simulations with no sex-differences in causal effect are presented in Figure 2 and summarized below. The full simulation results, including the male-specific causal effect estimation and sex differences in causal effect simulations are summarized in Supplementary Note 1 and Supplementary Figures 1, 2 and 3.

**Figure 2:**
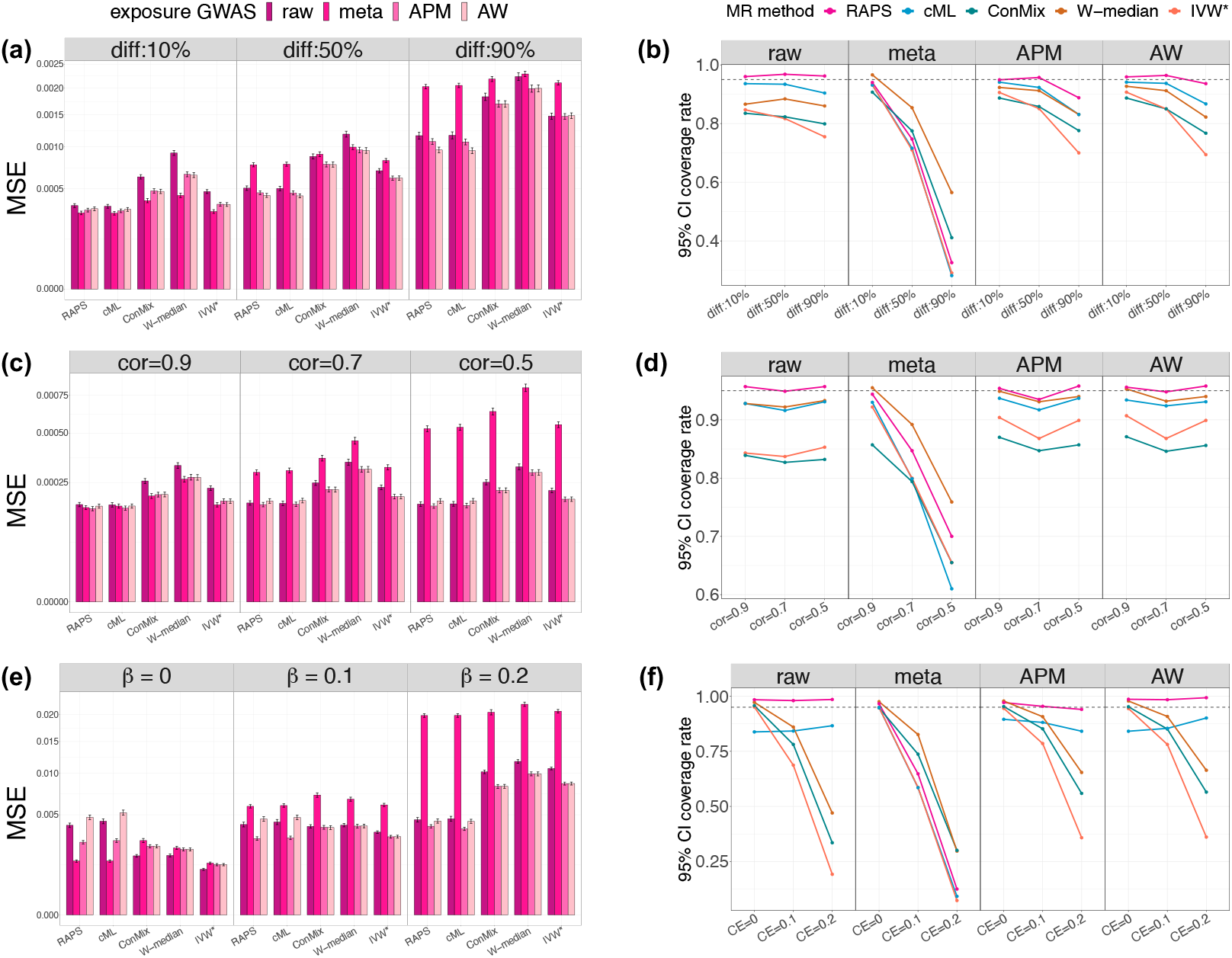
MSE and 95% confidence interval coverage rate for female-specific causal effect estimation Results from estimating female-specific causal effects *β*_*F*_ under three simulation scenarios where there are no sex differences in causal effects (i.e., *β*_*F*_ = *β*_*M*_) across sex groups. The left panel shows the MSEs of the estimated female-specific causal effects, while the right panel presents the 95% confidence interval coverage rates of the true underlying causal effects. We considered five two-sample MR methods for estimating the causal effect, which are: MR-RAPS (RAPS), constrained maximum likelihood (cML), contaminated mixture (ConMix), weighted median (W-median), penalized and robust IVW (IVW*). In the MSE results, the un-calibrated approach 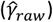 is represented by the bars with darker colors. The other three shrinkage approaches 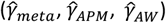 are shown in gradient colors. For the coverage rate results, the un-calibrated approach is shown in the first column, and other three shrinkage approaches are shown in the second to forth column. The results of fixed sex differences in variant-exposure effect simulation are presented in panels a and b. The results of random sex differences in variant-exposure effect simulations are shown in panels c and d. The results of using MVP OSA GWAS summary statistics to guide sex-specific variant-exposure effect simulation are shown in panels e and f. The underlying true causal effect is set at *β*_*F*_ = 0.1 in panels a, b, c, and d. The underlying true causal effects are shown in the top of panel e and bottom of panel f. MSEs were computed over 1000 simulation replicates. Intervals around the estimated MSE correspond to the MSE +/-one estimated standard error. Abbreviation: MSE: mean square error; MR: Mendelian randomization; APM: adaptive posterior mean; AW: adaptive weight; diff: different level of sex differences in variant-exposure effects; Cor: correlation between female and male variant-exposure effect; CE: causal effect; RAPS: MR-RAPS; cML: constrained maximum likelihood; ConMix: contaminated mixture; W-median: weighted medium; IVW*: penalized and robust IVW

### Results from simulation settings (i): fixed sex differences in variant-exposure effect sizes

These results are provided in Figures 2a and 2b. The 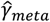 estimator performs best when *D*_*γ*_ included only 10% of the variants (i.e., |*D*_*γ*_ | = 10), compared to other shrinkage methods and to the raw 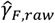.The estimators 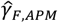 and 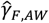 had similar performance, and both gave better causal effect estimates than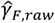,regardless of the proportion of variants with sex differences. We found that 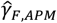 performed better than 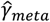 when more than 10% of variants were being selected into *D*_*γ*_. The coverage rate of *β*_*F*_ using 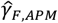 was lower than that of 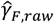 when many variants were included in *D*_*γ*_. However, 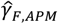 always achieved a smaller MSE for *β*_*F*_ compared with 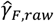.

### Results from simulation settings (ii): random sex differences in variant-exposure effect sizes

For random sex differences in *γ* (Figures 2c and 2d), when every variant has a sex difference, i.e., all *j* ∈ *D*_*γ*_, and most sex-difference *D*_*jγ*_ were strong, the estimate 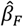 that relies on the 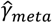 had a higher MSE than the one relying on raw 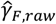.In contrast, using 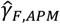 improved *β*_*F*_ estimation performance in terms of MSE and performed similarly to 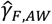.In these simulations, the APM estimator 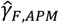 resulted in similar 95% confidence interval coverage compared to the raw approach, but achieved a smaller MSE for *β*_*F*_.

### Results from simulation settings (iii): using OSA GWAS summary statistics to guide the simulated variant-exposure effect sizes

Here, all variants have substantial sex differences in *γ*, while the selected variants are all weak IVs, meaning that the *F*-statistic < 10 in the female population. Using 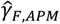 and 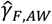 improved *β*_*F*_ estimation as demonstrated by improved MSE (Figures 2e and 2f). In most cases, 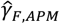 performed better 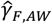.Both 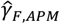 and 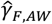 approaches improved the 95% confidence interval coverage rate compared to using 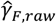.These results highlight that even though less information could be transferred from the male to the female population in these simulations (due to the strong sex differences in *γ*), adaptive estimates still improved causal effect estimation. Moreover, the estimates 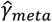 had a larger MSE than the raw approach under most settings and performed poorly when the female causal effect *β*_*F*_ was non-null.

In summary, borrowing power from the stratum with the larger to the stratum with the lower sample size, using adaptive (i.e., shrinkage) variant-exposure estimates (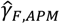 and 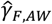) improves *β*_*F*_ estimates. Using estimators 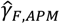 and 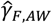 performed well in most simulation studies, regardless of the degree of sex differences *D*_*γj*_. The estimator 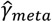 performs best when the underlying true *γ* were similar in the two groups. Among the two-sample MR methods considered, MR-RAPS, known to perform well when weak instruments are used, demonstrated at least no worse performance than other methods.

### Secondary simulation studies

Supplementary Note 2 and Supplementary Figures 4-14 provide results from secondary simulation studies. We expanded upon simulation settings (i), and further examined two scenarios: (a) estimation at the presence of pleiotropic effects of some IVs and (b) calibration of both *γ* and Γ effect estimates using the proposed framework. We also considered an increased sample size of the female population in the exposure GWAS to simulate a scenario where borrowing information from the male group may be less useful. Lastly, we evaluated the performance of a test for sex differences, i.e., a test of the null hypothesis *H*_0_: *β*_*F*_ = *β*_*M*_, using the estimated *β*_*F*_ and *β*_*M*_ based on various MR approaches (but always using the raw variant-exposure 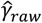 and variant-outcome associations 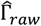).

In brief, 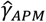 and 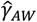 substantially improved the estimation accuracy of *γ* (Supplementary Figures 4-6) compared to the 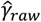 in all settings and, under substantial sex differences, performed better than 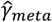.In the simulations where pleiotropy was present, 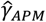 still produced a lower MSE when estimating *β*_*F*_ compared to 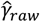 (Supplementary Figure 7). The penalized and robust IVW, contaminated mixture, cML, and MR-RAPS performed similarly in this analysis. Additional calibration of Γ improved the estimation of the *β*_*F*_ when males and females had the same causal effect, but not otherwise (Supplementary Figures 9-10). When increasing female sample sizes, the shrinkage approaches resulted nearly the same MSE for *β*_*F*_ estimation as the 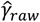,indicating that no estimation efficiency is lost when using the shrinkage approaches, even though potentially less information is transferred from the male population (Supplementary Figures 11-12). The test of sex differences in the causal effect between groups showed that MR-RAPS controls Type I error rate close to the nominal level under most settings while maintaining similar power compared to other MR methods for detecting sex differences in causal effects (Supplementary Figures 13-14).

### Sex-specific causal estimates of the effect of sleep phenotypes on cardiovascular-related outcomes

We estimated sex-specific causal effects of sleep phenotypes on CVD-related outcomes. The sample sizes for MVP’s sex-specific sleep GWAS are reported in Supplementary Table 4.

Specifically, we considered five binary sleep traits: OSA, insomnia, short sleep duration, long sleep duration, and excessive daytime sleepiness (sleepiness). The outcomes were six binary CVD-related phenotypes from All of Us: atrial fibrillation (AF), coronary artery disease (CAD), chronic kidney disease (CKD), heart failure (HF), hypertension (HTN), and type 2 diabetes mellitus (T2DM). Characteristics of AoU individuals in the variant-phenotype association analysis are summarized in Supplementary Table 5. The MVP sex-specific GWAS of sleep phenotypes were conducted using the same procedure described elsewhere (16). We only included individuals from the White Harmonized race/ethnicity and genetic ancestry (HARE) group, as the large HARE group. Based on these results, we selected variants and performed variant-outcome associations in the AoU dataset, focusing on the group of White individuals. For each trait, two types of analyses were performed: one adjusting for body mass index (BMI) and the other not adjusting for BMI. We only used common variants (minor allele frequency ≥ 0.01) with imputation quality score ≥ 0.8. More details are summarized in Method section. The results of sex-specific sleep GWASs are presented in Supplementary Figures 23-32.

### IV selection strategies

We applied p-value thresholding and clumping procedures on exposure GWASs using the “clump_data” function from the “TwoSampleMR” R package (version 0.6.8). The clumping window was set to 10,000 kb, correlation threshold to 0.001, and the European population reference panel was used. Due to having a limited number of variants (or even no variants) with p-value<5×10^−8^ and p-value<10^−7^ in the female sleep GWASs, p-value threshold of 10^−5^ was selected for IV selection in all analyses. The number of variants remaining after p-value thresholding (p-value<10^−5^) and clumping is given in Supplementary Table 3. We then matched the list of variants selected as targeted IVs from exposure GWAS to the variants in AoU datasets. The exposure and outcome dataset were harmonized using the “harmonise_data” function from the TwoSampleMR R package (version 0.6.8) to remove variants with unmatched allele frequency and palindromic patterns. We employed two strategies for IV selection in the primary analyses: (i) sex-specific IVs were selected based on sex-specific GWAS results 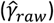,potentially resulting in different variants for male and female analyses; (ii) IVs were selected using APM shrinkage estimates 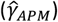,where additional “potential” IVs that could not be selected using 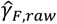 estimates (due to the smaller female sample size) were included by borrowing information from the male population. In the secondary analysis, we applied the 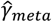 estimate for IV selection in sex-combined analyses.

### Sex-specific causal estimates

The primary results use the 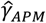 estimates with the MR-RAPS method, specifically developed for handling weak instruments and demonstrating superior performance in simulation studies. Figure 3 shows sex-specific causal effect estimates of sleep-related phenotypes on CVD-related outcomes, employing two different IV selection strategies: IVs selected based on sex-specific 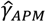 (primary, proposed) and sex-specific 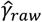.Three pairs of causal effect estimates showed statistically significant associations (p < 0.05) when IVs were selected using 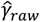:insomnia on CKD in females (OR: 1.23, 95% CI: 1.01–1.49), long sleep on HTN in females (OR: 1.03, 95% CI: 1.00–1.06), and short sleep on CAD in males (OR: 1.32, 95% CI: 1.03–1.69). In contrast, when 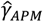 was used for IV selection, enabling more potential IVs to be selected by borrowing information across sex groups, additional statistically significant causal estimates were identified. For instance, in females, we found a significant effect of OSA on T2DM (OR: 1.32, 95% CI: 1.05–1.66) in BMI-unadjusted analyses, and also an effect on HTN (OR: 1.14, 95% CI: 1.03– 1.25) in BMI-adjusted analyses. Notably, the causal relationships between insomnia and CKD (OR: 1.38, 95% CI: 1.07–1.79) and long sleep and HTN (OR: 1.04, 95% CI: 1.00–1.09) were replicated in the APM selection analyses (both from BMI-adjusted analyses). Among males, multiple causal associations were observed between OSA and CVD-related outcomes, including OSA on CKD (BMI-unadjusted: OR: 1.23, 95% CI: 1.03–1.47; BMI-adjusted: OR: 1.27, 95% CI: 1.02–1.59) and OSA on HF in the BMI-unadjusted analysis (OR: 1.30, 95% CI: 1.04–1.63).

**Figure 3:**
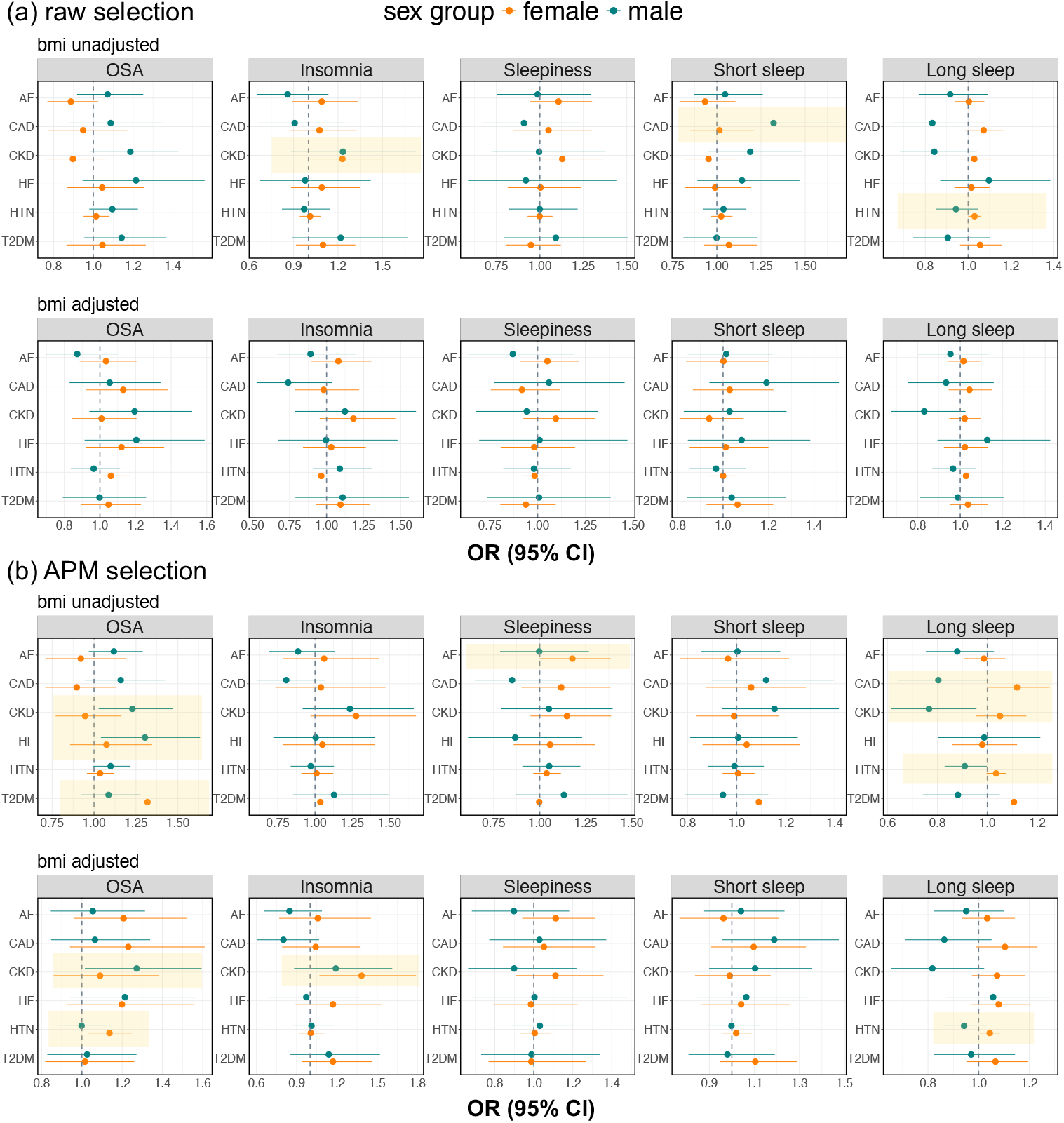
Results from sex-specific causal effect estimation Panel a provides sex-specific causal effect estimates with the corresponding 95% CIs based on IVs selected by using 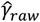,while panel b shows result using 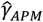 for IV selection. The estimated causal effects (from MR-RAPS) with 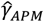 variant-exposure effect estimates are displayed on an OR scale. In each panel, variant-phenotype estimates without BMI adjustment are displayed at the top, and those with BMI adjustment are shown at the bottom. Female-specific results are indicated in the color orange, while results from male-specific are shown in color green. Vertical dashed lines indicate the null causal effects. The exposure variables are shown as the titles of the sub-panels, while row names of the sub-panels provide the outcome variables. Statistically significant results (p-value < 0.05) for either the female or male-specific analysis are highlighted with yellow background. Abbreviations: CI: confidence interval; APM: adaptive posterior mean; IV: instrumental variable; MR-RAPS: MR using robust adjusted profile score method; MR: Mendelian randomization; OR: odds ratio; BMI: body mass index; OSA: obstructive sleep apnea; AF: atrial fibrillation; CAD: coronary artery disease; CKD: chronic kidney disease; HF: heart failure; HTN: hypertension; T2DM: type 2 diabetes mellitus.

Sex-differences tests identified statistically significant difference in the causal effect of OSA on CKD, with a stronger effect in males (Table 3 and Supplementary Figure 15). Also, there were sex differences in the causal effects of long sleep on several CVD-related outcomes: long sleep increased risk of CAD, CKD, HTN, and T2DM in females, but was protective in males. These sex differences were statistically significant in the APM IV selection analysis (Table 3 and Supplementary Figures 17-18).

The key findings from sex-specific causal effect estimation and sex-difference tests are summarized in Tables 2 and 3. Full results, including the causal estimation using 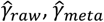,and 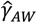,as well as the sex-differences in causal estimates tests, are summarized in Supplementary Figures 15-18. Causal estimates from other considered MR methods are summarized in Supplementary Data 1 (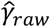 IV selection) and Supplementary Data2 (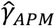 IV selection).

**Table 2:**
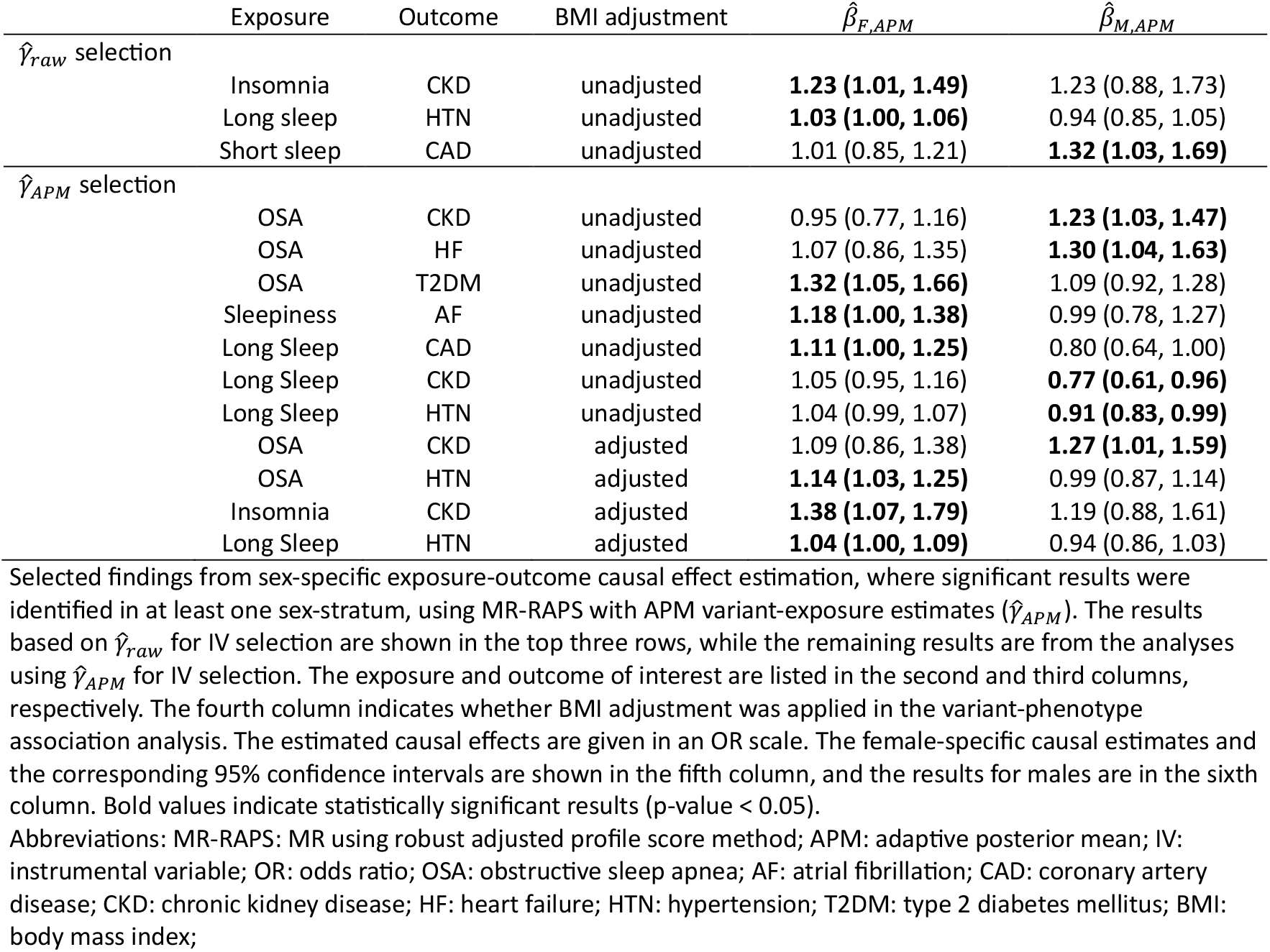
Top findings from sex-specific exposure-outcome causal effect estimation.

| | Exposure | Outcome | BMI adjustment | $\hat{\beta}_{F,APM}$ | $\hat{\beta}_{M,APM}$ |
| --- | --- | --- | --- | --- | --- |
| $\hat{\gamma}_{raw}$ selection | | | | | |
|  | Insomnia | CKD | unadjusted | **1.23 (1.01, 1.49)** | 1.23 (0.88, 1.73) |
|  | Long sleep | HTN | unadjusted | **1.03 (1.00, 1.06)** | 0.94 (0.85, 1.05) |
|  | Short sleep | CAD | unadjusted | 1.01 (0.85, 1.21) | **1.32 (1.03, 1.69)** |
| $\hat{\gamma}_{APM}$ selection | | | | | |
|  | OSA | CKD | unadjusted | 0.95 (0.77, 1.16) | **1.23 (1.03, 1.47)** |
|  | OSA | HF | unadjusted | 1.07 (0.86, 1.35) | **1.30 (1.04, 1.63)** |
|  | OSA | T2DM | unadjusted | **1.32 (1.05, 1.66)** | 1.09 (0.92, 1.28) |
|  | Sleepiness | AF | unadjusted | **1.18 (1.00, 1.38)** | 0.99 (0.78, 1.27) |
|  | Long Sleep | CAD | unadjusted | **1.11 (1.00, 1.25)** | 0.80 (0.64, 1.00) |
|  | Long Sleep | CKD | unadjusted | 1.05 (0.95, 1.16) | **0.77 (0.61, 0.96)** |
|  | Long Sleep | HTN | unadjusted | 1.04 (0.99, 1.07) | **0.91 (0.83, 0.99)** |
|  | OSA | CKD | adjusted | 1.09 (0.86, 1.38) | **1.27 (1.01, 1.59)** |
|  | OSA | HTN | adjusted | **1.14 (1.03, 1.25)** | 0.99 (0.87, 1.14) |
|  | Insomnia | CKD | adjusted | **1.38 (1.07, 1.79)** | 1.19 (0.88, 1.61) |
|  | Long Sleep | HTN | adjusted | **1.04 (1.00, 1.09)** | 0.94 (0.86, 1.03) |

Selected findings from sex-specific exposure-outcome causal effect estimation, where significant results were identified in at least one sex-stratum, using MR-RAPS with APM variant-exposure estimates ( $\hat{\gamma}_{APM}$ ). The results based on $\hat{\gamma}_{raw}$ for IV selection are shown in the top three rows, while the remaining results are from the analyses using $\hat{\gamma}_{APM}$ for IV selection. The exposure and outcome of interest are listed in the second and third columns, respectively. The fourth column indicates whether BMI adjustment was applied in the variant-phenotype association analysis. The estimated causal effects are given in an OR scale. The female-specific causal estimates and the corresponding 95% confidence intervals are shown in the fifth column, and the results for males are in the sixth column. Bold values indicate statistically significant results (p-value < 0.05).
Abbreviations: MR-RAPS: MR using robust adjusted profile score method; APM: adaptive posterior mean; IV: instrumental variable; OR: odds ratio; OSA: obstructive sleep apnea; AF: atrial fibrillation; CAD: coronary artery disease; CKD: chronic kidney disease; HF: heart failure; HTN: hypertension; T2DM: type 2 diabetes mellitus; BMI: body mass index;

**Table 3:** Top findings from sex-differences test in causal effect estimation.

| | Exposure | Outcome | $\hat{\beta}_{F,APM}$ | $\hat{\beta}_{M,APM}$ | $\hat{\beta}_{F,raw} - \hat{\beta}_{M,raw}$ | Sex difference p-value | Sex difference FDR p-value |
| --- | --- | --- | --- | --- | --- | --- | --- |
| $\hat{\gamma}_{raw}$ selection | | | | | | | |
| BMI-unadjusted |  |  |  |  |  |  |  |
|  | OSA | CKD | -0.108 (-0.277, 0.060) | 0.171 (-0.015, 0.357) | -0.275 (-0.521, -0.029) | 0.028 | 0.500 |
| $\hat{\gamma}_{APM}$ selection | | | | | | | |
| BMI-unadjusted |  |  |  |  |  |  |  |
|  | Long Sleep | CAD | 0.112 (0.001, 0.223) | -0.219 (-0.445, 0.008) | 0.328 (0.078, 0.578) | 0.009 | 0.118 |
|  | Long Sleep | CKD | 0.049 (-0.046, 0.146) | -0.267 (-0.489, -0.044) | 0.317 (0.073, 0.560) | 0.011 | 0.118 |
|  | Long Sleep | HTN | 0.035 (-0.003, 0.072) | -0.095 (-0.188, -0.002) | 0.128 (0.028, 0.228) | 0.011 | 0.118 |
|  | Long Sleep | T2DM | 0.101 (-0.022, 0.225) | -0.125 (-0.299, 0.049) | 0.223 (0.014, 0.433) | 0.036 | 0.273 |
| $\hat{\gamma}_{APM}$ selection | | | | | | | |
| BMI-adjusted |  |  |  |  |  |  |  |
|  | Long Sleep | CKD | 0.070 (-0.027, 0.168) | -0.202 (-0.425, 0.022) | 0.272 (0.028, 0.516) | 0.029 | 0.407 |
|  | Long Sleep | CAD | 0.099 (-0.011, 0.209) | -0.146 (-0.341, 0.049) | 0.241 (0.019, 0.463) | 0.033 | 0.407 |
|  | Long Sleep | HTN | 0.043 (0.004, 0.082) | -0.059 (-0.148, 0.029) | 0.101 (0.004, 0.197) | 0.041 | 0.407 |

Selected findings from tests of sex-difference in causal effects. The first column describes the IV selection procedure and whether the exposure GWAS was adjusted for BMI or not. The exposure and outcome of interest are provided in the second and third columns, respectively. Sex-specific causal estimates, obtained using MR-RAPS with APM variant-exposure estimates ( $\hat{\gamma}_{APM}$ ), along with their corresponding 95% confidence intervals, are displayed in the fourth (female-specific) and fifth (male-specific) columns. The estimated causal effects are reported on the log scale, as used in the test of sex differences. The sixth column provides the estimated sex differences in causal effects, calculated from raw variant-exposure estimates ( $\hat{\gamma}_{raw}$ ), with 95% confidence intervals. P-values and FDR-adjusted p-values for the statistical tests of sex differences are shown in the seventh and eighth columns. The FDR p-values were computed using the Benjamini-Hochberg procedure. All the sex-difference tests are significant in the nominal threshold (p-value < 0.05).
Abbreviations: IV: instrumental variable; GWAS: genome-wide association study; BMI: body mass index; BMI-unadj: BMI unadjusted; BMI-adj: BMI adjusted; MR-RAPS: MR using robust adjusted profile score method; APM: adaptive posterior mean; OR: odds ratio; OSA: obstructive sleep apnea; CAD: coronary artery disease; CKD: chronic kidney disease; HTN: hypertension; T2DM: type 2 diabetes mellitus; FDR p-value: false discovery rate adjusted p-value.

**Table 4:** Two-sample MR methods used in both simulation studies and real data analyses.

| MR method | Description | Software used | Reference |
| --- | --- | --- | --- |
| **MR-RAPS**<br>**(primary**<br>**analysis)** | Estimation: adjusted profile likelihood estimation with down-weighting of outliers via a robust loss function<br>Assumptions: InSIDE, pleiotropic effects follow normal distribution with mean zero | R package: mr.raps<br>R function: mr.raps.overdispersed.robust() | (30) |
| **IVW** | Estimation: aggregate multiple Wald-ratio estimates using fixed effect inverse-variance weighting<br>Assumptions: InSIDE, zero average pleiotropy effect | R package: MendelianRandomization<br>R function: mr_allmethods() | (27) |
| **Constrained**<br>**ML** | Estimation: maximum likelihood estimation with a constraint on the number of invalid IVs<br>Assumptions: plurality valid | R package: MendelianRandomization<br>R function: mr_cML() | (42) |
| **Contamination**<br>**mixture** | Estimation: profile likelihood estimation assuming that the ratio | R package: MendelianRandomization | (41) |
|  | estimates follow two normal distributions for valid and invalid IVs, respectively. | R function: `mr_conmix()` |  |
|  | Assumption: plurality valid |  |  |
| **Weighted median** | Estimation: compute the median of the causal effect estimated from multiple IVs, weighted by the inverse of the estimate's sampling variance | R package: `MendelianRandomization`<br>`mr_allmethods()` | (47) |
|  | Assumption: majority valid |  |  |
| **MR Egger** | Estimation: weighted meta-regression with an intercept term to capture the average horizontal pleiotropy effect | R package: `MendelianRandomization`<br>R function: `mr_allmethods()` | (39) |
|  | Assumption: InSIDE |  |  |
| **Robust IVW and MR-Egger** | Estimation: apply robust regression to down-weight or exclude variants with heterogenous causal estimates | R package: `MendelianRandomization`<br>R function: `mr_allmethods()` | (40) |

Summary of the two-sample MR methods used in the simulation studies and real data analyses. The names of the methods are listed in the first column. A brief description of the estimation approach and the underlying assumptions for each method are provided in the second column. The corresponding software for applying these methods is listed in the third column, and the references are provided in the last column Abbreviations: InSIDE: instrument strength independent of direct effect.

#### Results from secondary analyses

We compared the causal effect estimates of MR-RAPS, which we used in the primary analyses, with those of MR-PRESSO (43), a widely used approach for detecting IVs with pleiotropic effects, and removing them, in MR analysis. The results from two methods are similar and are summarized in Supplementary Note 3 and Supplementary Figures 19-20.

A comparison of causal effect estimates 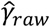 and 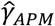 for IV selection is presented in Supplementary Figure 21. In the male population, more consistent results were observed across different IV selection methods. For the analysis of OSA phenotype, the 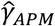 selection strategy identified more variants as IVs in both male and female populations. Several significant causal effects between OSA and CVD-related outcomes were only detected when 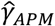 was used for IV selection. These findings highlight an additional advantage of our proposed shrinkage estimate: it not only helps correct for weak IV bias in causal effect estimation but also enhances the IV selection process, increasing the potential for identifying novel causal effects.

Lastly, we applied 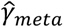 for IV selection in the sex-combined analysis. In this analysis, the variant-exposure effect estimates were based on 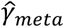,and the variant-outcome effect estimates were computed from a sex-combined analysis in the AoU study. The results are shown in Supplementary Figure 22. Significant causal relationships between OSA and several CVD-related outcomes were identified, consistent with the findings from the primary analysis using 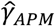 for IV selection. However, some of the significant sex-specific findings, such as the female-specific causal effect of long sleep on HTN, were not identified in sex-combined analysis, likely because the 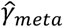 is closed to 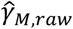 due to the predominantly male sample size in MVP.

## Discussion

We performed sex-specific analysis of the causal associations of sleep-related phenotypes on CVD-related outcomes. These are biologically important relationships with immediate clinical relevance. The primary rationale in our work was to ameliorate the weak instrumental bias in MR analysis. This can be done by incorporating information from auxiliary datasets. In our case, we used a male-specific dataset to improve female-specific statistics. Acknowledging likely sex differences between male and female individuals, this led to the need for an adaptive estimator, that will intelligently utilize information across the two sex groups. Female-specific causal estimation is limited by smaller sample sizes (relative to males), leading to large variability in *γ*_*F*_ estimates, also known as weak IV bias. Thus, we introduced a framework to calibrate the 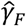 estimates by borrowing information from the male group. We first demonstrated that the FE meta estimate 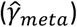 is a special case of our proposed framework. We then proposed the APM estimate, which adaptively transfers information across sex groups by considering the strength of sex difference in *γ*, in a data-driven manner. Simulation studies demonstrated that: (i) employing the shrinkage estimates can substantially improve the efficiency of causal estimation for the population with smaller sample size; (ii) using 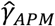 estimate is less sensitive to the existence of sex differences in *γ* compared to the use of 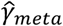;(iii) no estimation efficiency is lost by applying the 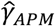 to the population with larger sample sizes. In real data analyses, we identified several sex-difference patterns between the causal association of sleep phenotypes and CVD-related outcomes, including OSA on CKD as well as long sleep duration on several CVD-related outcomes, offering potential implications for research in sex-specific cardiovascular medicine. The method itself has broader applicability, and could be used to address sex differences for a large set of complex traits.

We also applied the proposed framework to the male population, which had a larger sample size in our analysis. The results indicate that less information could be borrowed from the smaller female population. Although the shrinkage approaches did not improve causal effect estimation for the male population in our simulations, using them did not result in a loss of estimation efficiency compared to using the raw 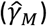.This supports the usefulness of incorporating shrinkage approaches (especially the adaptive approach) into MR analysis when relevant summary statistics are available, regardless of the corresponding sample size.

Due to the low number of IVs that could be used when applying a genome-wide significance threshold (5×10^−8^) for selecting IVs, we considered a lower p-value threshold (p-value < 10^−5^), as suggested by (27) as the minimal threshold value for selecting IVs in two-sample MR analysis. However, this strategy may increase the risk of including several weak instruments in MR analysis. Therefore, we applied advanced MR methods that address weak instrumental bias, including MR-RAPS (30), cML (42), the contaminated mixture model (41), and robust MR methods (40), in simulations and in real data analyses. We used MR-RAPS, which demonstrated superior performance in our simulation studies, as the primary method. The consistent findings between MR-RAPS and cML, the top two methods with the best estimation performance in simulation studies, also increase confidence in the data analyses findings. Additionally, to examine the influence of violating horizontal pleiotropy assumption due to using a lower p-value threshold to select IVs, we compared the causal effect estimated by MR-RAPS with MR-PRESSO (43), another widely used approach developed for detecting horizontal pleiotropy effects, in our primary and secondary analyses. The results from MR-RAPS and MR-PRESSO are highly consistent, increasing the expected reliability of our findings using MR-RAPS.

Motivated by the need to address low female sample size in MVP data and the use of its summary statistics as exposure GWAS in MR analysis, our focus has been calibrating the “variant-exposure” effect estimates. However, we did not apply such calibrations to the outcome GWASs, as the balanced sample sizes across sex-specific GWAS in AoU suggest that further calibrations might not substantially improve the efficiency of sex-specific summary statistics, compared to the exposure GWASs. Simultaneously incorporating the proposed Bayesian framework into exposure and outcome GWASs will be of interest if both GWASs were performed with limited sample sizes.

Some limitations of this work should be discussed. First, the lack of sex-specific sleep GWASs limits the selection of IVs using independent datasets. Instead, we directly utilized the MVP sleep GWASs for the IV selection. Various IV selection strategies were implemented, including sex-specific selection, sex-specific APM estimates selection in primary analysis, and sex-combined selection by using FE meta estimate in secondary analysis. The results, however, are only sometimes consistent across different selection methods, indicating that the estimation of causal effects is sensitive to the IV selection process. This sensitivity could be due to the strength of the IVs and differences in the underlying genetic architecture captured by the different IVs. The second limitation arises from the test of sex differences in exposure-outcome causal effects. We applied the conventional two-sample t-test, performed under the assumption of independence between the two compared samples. Therefore, we included only the causal estimates from the raw estimates (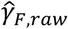 and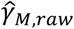) in our sex-differences test. We did not use any shrinkage estimates, because they may result in correlated causal effect estimates between sexes. A correlated version, which includes a covariance correction between estimates (32), can be exploited. However, quantifying the covariance between causal estimates across sexes using shrinkage approaches is challenging and a topic of future work. Lastly, we used only European ancestry individuals, due to the larger sample sizes available. Assessing the transferability of our findings to other populations, and potentially applying a similar shrinkage framework to improve IVs in non-European ancestry populations, is of interest for future research.

## Methods

### Sex-specific sleep GWAS in MVP

We performed sex-specific GWAS for four sleep phenotypes (insomnia, long sleep duration, short sleep duration, and excessive daytime sleepiness), and used GWAS summary statistics from previously published GWAS of OSA (16), using MVP participants. Sleep phenotypes are described below. We only used the European (White) harmonized race/ethnicity and genetic ancestry (HARE) group to match the genetic ancestry of the available outcome sex-stratified GWAS from the AoU. We removed related individuals based on kinship coefficient ≥ 0.0884, where the kinship coefficients were estimated using KING v.2.0 (44), so that all analyses were conducted using an unrelated set of individuals (including, participants from across the female and male strata were unrelated). Sex chromosome checks confirmed biological sex. Variants were filtered based on imputation quality, requiring INFO score of at least *R*^2^ ≥ 0.6, and minor allele frequency ≥ 0.01. The sample sizes of each sleep GWAS are summarized in Supplementary Table 4. Across analyses, sample sizes ranged from ∼15,000 to 30,000 (female stratum), and from ∼204,000 to 380,000 (male stratum). All analyses were adjusted for age and the first 10 PCs of genetic data. For each trait, two GWASs were performed: with and without BMI adjustment. BMI-adjusted models had BMI as a covariate using both linear and squared terms. Analyses were performed separately using PLINK v2.00a3LM (45) in each sex strata.

### Sleep phenotypes

Sleep phenotypes were defined as previously reported (16). In detail, OSA was defined based on the VA electronic health record using a multimodal automated phenotyping (MAP) algorithm (46). The MAP algorithm was applied to predict general sleep apnea and OSA separately. For each trait, it resulted in a score, roughly mapping to a likelihood of having these condition. The final OSA definition combined both scores. Insomnia status was inferred using a MAP algorithm applied to relevant ICD codes and elements extracted using natural language processing. Insomnia status was inferred at the same age as was used in OSA analysis, i.e., a participant was considered to have insomnia if their age of first insomnia ICD code was on or before their age in the OSA analysis. All other sleep phenotypes were self-reported based on the baseline questionnaire administered to program participants. Long sleep was defined as sleep duration > 9, and short sleep as sleep duration < 6, where the sleep duration was defined based on the response to the question about hours of sleep in a typical day, with responses ranging from “5 or less” to “10 or more”, with increments of 0.5. The excessive daytime sleepiness phenotype was based on the questionnaire item “Feeling excessively sleepy during the day (does not include regular naps),” with yes or no answers. All five sleep phenotypes are binary.

### Association analysis with CVD-related outcomes in AoU

We used short-read whole-genome sequencing (srWGS) data (version 7) from the AoU study to conduct associations analysis with CVD-related outcomes. To reduce memory storage requirements, we focused on genomics data pre-filtered by the following criteria: population-specific allele frequency ≥ 1% or population-specific allele count > 100 (data from: gs://fc-aou-datasets-controlled/v7/wgs/short_read/snpindel/acaf_threshold_v7.1). To align with summary statistics computed from the White HARE group in the MVP, we included only White individuals (as determined by self-reported race and ethnicity) in the AoU analysis. Related individuals were excluded based on information available in gs://fc-aou-datasets-controlled/v7/wgs/short_read/snpindel/aux/relatedness/relatedness_flagged_samples.tsv. We also restricted the analysis to adults aged 18 to 95, with BMI values ranging from 17 to 55.

Following data preprocessing, approximately 114,000 White individuals were included, consisting of 67,600 females and 46,400 males. The exact sample size varied slightly depending on the phenotype analyzed. Six binary CVD-related phenotypes were considered: atrial fibrillation (AF), coronary artery disease (CAD), cardiovascular disease (CVD), heart failure (HF), hypertension (HTN), and type 2 diabetes mellitus (T2DM). Detailed selection criteria for these clinical outcomes and the corresponding SNOMED codes and OMOP Concept IDs in the AoU study, are provided in Supplementary Table 6. More details are summarized in Supplementary Note 4.

After matching the variants identified as IVs according to the exposure dataset (MVP), we conducted single-variant association analyses with the six binary outcomes. We used logistic regression, adjusting for age and 16 genetic principal components (PCs) as covariates in the BMI-unadjusted analyses. For the BMI-adjusted analyses, the models were further adjusted for BMI, including both linear and quadratic terms. The effect sizes of the variants, i.e., log(OR) with the corresponding SD, were then extracted for use as summary statistics in the outcome GWAS.

### Simulation studies

We performed simulation studies to evaluate the performance of exposure-outcome causal effect (*β*_*F*_ and *β*_*M*_) estimation, using shrinkage variant-exposure effect estimates (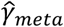 and 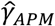) as input to existing two-sample MR approaches. We then compared these to the use of un-calibrated (“raw”) effect estimates 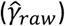.We employed three settings to simulate sex-specific variant-exposure effect sizes (*γ*_*F*_ and *γ*_*M*_) and considered different levels of sex difference in causal effect within each setting.

### Simulation data generation

We simulated data via the structural model presented in Figure 1a, where we use *S* (for sleep phenotype) to represent the exposure variable and *C* (for CVD phenotype) to represent the outcome variable. We first generated individual-level genetic data for two datasets, corresponding to two populations used in an exposure GWAS (one population) and an outcome GWAS (second population). In each of the datasets, the data-generating process was carried out independently in the females and males. We set the female sample size of the exposure dataset to 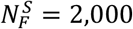 and the sample size of the male stratum to 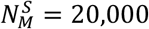 matching the sex sample size proportions in the MVP. In the outcome dataset, the female and male sample sizes were set to be the same: 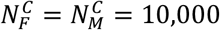.For each genetic variant, allele counts were generated independently from *Binomial*(2,0.3) distribution, assuming that all variants are in linkage equilibrium with allele frequencies of 0.3.

In the exposure dataset, we generated the exposure variable according to the following linear model:

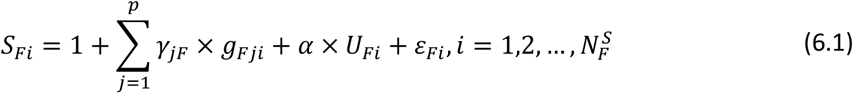

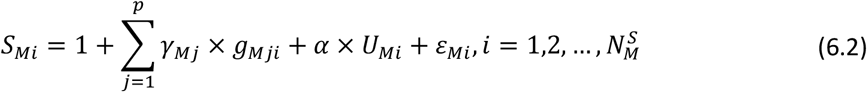

With *p* = 100 independent variants. *U* represents the unknown confounder, which we generated from Normal distribution with mean 0 but different variances in female and male populations: *U*_*Fi*_∼*N*(0,1) and *U*_*Mi*_∼*N*(0,0.5). The random errors *ε*_*Fi*_ and *ε*_*Mi*_ were generated from standard Normal distribution for both the female and male populations. The *γ*_*jF*_ and *γ*_*Mj*_ are the underlying sex-specific variant-exposure effect sizes. The effect size of the unknown confounder to the exposure association was set as *α* = 0.1 for both the female and male populations.

Next, we use the same strategy to generate the genetic data, exposure variable, and unmeasured confounder for the outcome population,

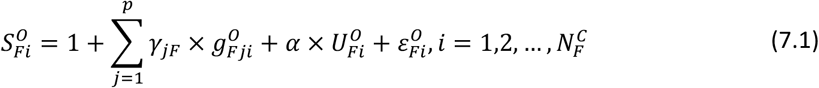

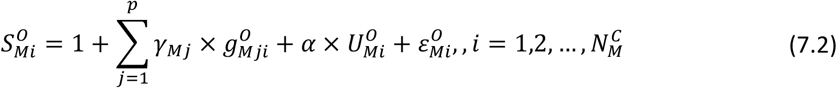

The superscript *O* is used to distinguish the exposure simulated in the outcome population from the exposure simulated in the exposure population. Based on the generated exposure variables 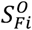 and 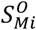, we then generate the outcome variable through the following linear equation:

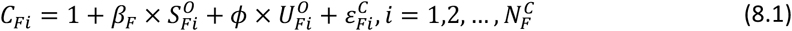

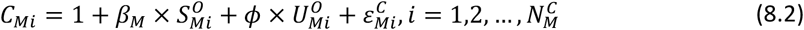

where *ϕ* denotes the effect sizes of the unknown confounder on the outcome, which we set to *ϕ* = 0.1. The *β*_*F*_ and *β*_*M*_ are the underlying true female and male exposure-outcome causal effects, respectively.

After generating the individual level data, we get the exposure GWAS and outcome GWAS summary statistics by fitting marginal linear regression models. The exposure data summary statistics 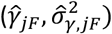 and 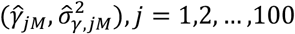 are obtained by fitting the following regression using the data generated from equations (6.1) and (6.2):

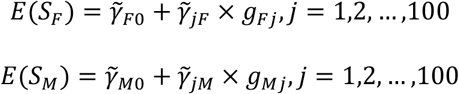

And the same procedure is applied for the outcome population

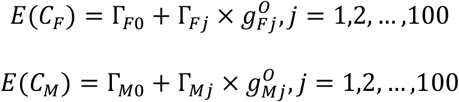

The outcome GWAS summary statistics are 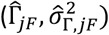 and 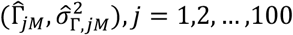.

We performed simulations in three simulation settings that differed in the way that variant-exposure effect sizes were simulated. In all settings, we considered a few models for the exposure-outcome effect sizes: no sex differences in exposure-outcome causal effects, with *β*_*F*_ = *β*_*M*_ = 0.1; and sex differences in exposure-outcome causal effect with *β*_*M*_ = 0.1, and *β*_*F*_ ∈ {0,0.05,0.15}.

### Simulation settings 1: Fixed sex differences in variant-exposure effects

Here, we simulated from settings in which the sex-difference in *γ*_*F*_ and *γ*_*M*_, when there were such differences, were weak and fixed across variants. Male-specific variant-exposure effect sizes were *γ*_*jM*_ = 0.1, *j* = 1,2, …, 100 for all variants. For the female population, some variants had the same effect size as in the male population (*γ*_*jF*_ = 0.1), and a subset of variants (sized 10, 50, or 90 out of 100) had female-specific variant-exposure effect size *γ*_*jF*_ = 0.05. For the sex differences in causal effect setting, we fixed 50% of variants to have sex differences in variant-exposure effect sizes.

### Simulation settings 2: Random sex differences in variant-exposure effects

Here, we simulated from settings in which the sex-difference in variant-exposure effect sizes, when there were such differences, were strong. For each variant *j* = 1,2, …, 100, we generated a pair of female and male variant-exposure effect size from a bi-variate normal distribution 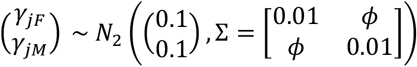, where we considered different covariance value *ϕ* to set the correlations 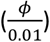 between (*γ*_*F*_, *γ*_*M*_) equaling to either 0.5, 0.7, and 0.9 when the exposure-outcome causal effect was the same in males and females (*β*_*F*_ = *β*_*M*_ = 0.1). When *β*_*M*_ ≠ *β*_*F*_, the correlation between (*γ*_*F*_, *γ*_*M*_) was set to 0.7. Finally, the variant exposure effect sizes *γ*_*F*_, *γ*_*M*_ were randomly generated in each simulation replicate.

### Simulation settings 3: Summary statistics from MVP OSA GWAS guide variant-exposure effect sizes

Here, we used the summary statistics from the MVP OSA GWAS (without BMI adjustment) to guide the *γ*_*F*_ and *γ*_*M*_ values. First, we conducted p-value thresholding and clumping for the male-specific summary statistics. The p-value threshold was set at 10^−5^, with clumping window of 10,000 kb and a correlation threshold of 0.001, using the European population reference panel from 1000 genome. This resulted in a list of 110 variants. We randomly selected 100 variants used their male- and female-specific effect size estimates from the MVP GWAS in simulations.

### Two-sample MR methods

We applied the following two-sample MR methods, described in Table 4, over the raw and calibrated estimated variant effect sizes.

## Data availability

Summary statistics from sex-specific sleep trait GWASs will become available on the dbGaP repository “Veterans Administration (VA) Million Veteran Program (MVP) Summary Results from Omics Studies”, study accession phs001672. Data from the NIH All of Us study are available via institutional data access for researchers who meet the criteria for access to confidential data. To register as a researcher with All of Us, researchers may use the following URL and complete the laid-out steps: https://www.researchallofus.org/register/. The srWGS genomic data were available on: gs://fc-aou-datasets-controlled/v7/wgs/short_read/. The R code used to implement the proposed semi-empirical Bayes framework, two-sample MR approaches, and simulation studies is available on the GitHub repository: https://github.com/Gene-Huang/sex-specific-MR. The harmonized summary statistics for both exposure and outcome GWASs used in our data analysis are also provided in the repository.

## Acknowledgements

We are grateful to the Million Veteran Program participants and staff. This research is based on data from the Million Veteran Program, Office of Research and Development, Veterans Health Administration, and was supported by the Million Veteran Program-MVP000 and BX004821.

This publication does not represent the views of the Department of Veteran Affairs or the United States Government. The Million Veteran Program core acknowledgements for publications are stated in Supplementary Note 5. We gratefully acknowledge All of Us participants for their contributions and also thank the National Institutes of Health’s All of Us Research Program for making available the participant data examined in this study. The All of Us Research Program is supported by the National Institutes of Health, Office of the Director: Regional Medical Centers: 1 OT2 OD026549; 1 OT2 OD026554; 1 OT2 OD026557; 1 OT2 OD026556; 1 OT2 OD026550; 1 OT2 OD 026552; 1 OT2 OD026553; 1 OT2 OD026548; 1 OT2 OD026551; 1 OT2 OD026555; IAA #: AOD 16037; Federally Qualified Health Centers: HHSN 263201600085U; Data and Research Center: 5 U2C OD023196; Biobank: 1 U24 OD023121; The Participant Center: U24 OD023176; Participant Technology Systems Center: 1 U24 OD023163; Communications and Engagement: 3 OT2 OD023205; 3 OT2 OD023206; and Community Partners: 1 OT2 OD025277; 3 OT2 OD025315; 1 OT2 OD025337; 1 OT2 OD025276. The of Us Research Program would not be possible without the partnership of its participants. This research was further supported by National Institute on Aging grant R01AG080598 and by the National Heart, Lung, and Blood Institute grant R01HL161012.

## Author contributions

Conceptualization: TS. Methodology: Y-JH, TS. Formal analysis: Y-JH, NK. Resources, Project administration, Funding acquisition: KC, PWFW. Data Curation: KC, JEH. Writing – original draft: Y-JH, TS. Writing – review & editing: NK, DFL, JG, JEH, KC, PWFW, DJG, KMR. Visualization: Y-JH. Supervision: TS.

