## Supplementary information for "A semi-empirical Bayes approach for calibrating weak instrumental bias in sex-specific Mendelian randomization studies"

### Supplementary Note 1: Primary simulation studies

#### Overview of simulation settings and summary of primary results

To mimic the female-to-male sample size proportions in the Million Veteran Program (MVP), where we used the corresponding GWAS summary statistics as the exposure GWAS in two-sample Mendelian randomization (MR) analyses, we set the female exposure population sample size to 2,000 and the male exposure population to 20,000. For simplicity, we balanced the sample sizes for the outcome phenotype population between females and males, setting both to 10,000. In these simulation studies, the smaller sample size for females may introduce weak instrumental variable (IV) bias, potentially affecting the accuracy of female-specific exposure-outcome causal effect estimates ($\hat{\beta}_F$). As in the main manuscript, in what follows, we use $\gamma$ to represent “variant-exposure” effect size and $\Gamma$ to represent “variant-outcome” effect size. We evaluated using the shrinkage variant-exposure effect estimates ($\hat{\gamma}_{meta}, \hat{\gamma}_{APM}, \hat{\gamma}_{AW}$) for estimating the causal effect of interest ($\beta_F$), and compared it to the use of uncalibrated, raw, estimates ($\hat{\gamma}_{raw}$). In all the simulations, we considered 100 independent variants as IVs, all with allele frequency equal to 0.3. The results from simulation studies are summarized over 1000 simulation replicates.

We conducted simulations in a few settings. First, we compared sex differences in the exposure-outcome causal effect settings: no sex differences ($\beta_F = \beta_M$) and sex differences ($\beta_F \neq \beta_M$). For the sex difference setting, we fixed $\beta_M = 0.1$ and varied the strength of $\beta_F$ ($\beta_F = 0, 0.05, 0.15$). In settings with no sex differences in the exposure-outcome causal effects, we set $\beta_F = \beta_M = 0.1$, and considered different numbers of variants having sex differences in $\gamma$ (notation $|D_\gamma|$ in main manuscript), as well as different strengths of sex differences effect in $\gamma$. The details of the simulation settings are summarized in Supplementary Tables 1 and 2.

#### Summary of female-specific exposure-outcome causal effect estimation

We used boxplot to display the distribution of $\hat{\beta}_F$ using either shrinkage estimates ($\hat{\gamma}_{meta}$, $\hat{\gamma}_{APM}$, $\hat{\gamma}_{AW}$) or $\hat{\gamma}_{raw}$ as input for MR analysis. The results are presented in Supplementary Figure 3. The boxplot illustrates $\hat{\beta}_F$ across 1000 simulation replicates, with the horizontal dashed red line indicating the underlying $\beta_F$ value.

In the simulation with fixed strength sex differences in $\gamma$ (Supplementary Figure 3, panel a and b), most MR methods achieved unbiased estimation of $\beta_F$, except the MR-Egger approach. The shrinkage estimates resulted in smaller variance in causal effect estimates than the use of $\hat{\gamma}_{raw}$. In the settings where 90% of the variants exhibit sex differences in $\gamma$, only the cML and MR-RAPS methods resulted in unbiased estimates using $\hat{\gamma}_{raw}$. In contrast, while cML and MR-RAPS resulted in slightly lower average causal effect estimates when using $\hat{\gamma}_{APM}$, they achieved smaller variance in $\hat{\beta}_F$ compared to using $\hat{\gamma}_{raw}$. Therefore, the overall MSE of the causal effect estimate was smaller than that of using $\hat{\gamma}_{raw}$. (Figure 2a in the main manuscript).

In simulations with random strengths of sex differences in $\gamma$ (Supplementary Figure 3, panel c and d), the $\hat{\gamma}_{meta}$ approach produced biased estimates when the correlation between $\gamma_F$ and $\gamma_M$ was low or moderate (correlation = 0.5 and 0.7). However, the $\hat{\gamma}_{APM}$ and $\hat{\gamma}_{AW}$ approaches still produced unbiased or nearly unbiased estimates in these settings. The $\hat{\gamma}_{meta}$ approach produced unbiased estimates for all MR methods in high-correlation setting. The improvement from using the $\hat{\gamma}_{APM}$ approach was more significant in the weighted median and IVW these two methods, which produced estimates closer to the underlying true causal effect on average compared to using $\hat{\gamma}_{raw}$. The cML and MR-RAPS methods produced unbiased estimates in all settings, regardless of whether the shrinkage or raw estimate was used.

Of the simulation using MVP OSA GWAS to guide $\gamma$ effect size (Supplementary Figure 3, panel e and f), all methods provided unbiased estimates in the null causal effect setting ($\beta_F = 0$), regardless of $\beta_M$. As the true causal effect increased, the bias also increased with the $\hat{\gamma}_{meta}$ approach, but this had less influence on the methods using the $\hat{\gamma}_{APM}$ or $\hat{\gamma}_{AW}$ approaches. The $\hat{\gamma}_{APM}$ approach significantly reduced the variance of causal effect estimates compared to the use of $\hat{\gamma}_{raw}$, especially in the cML and MR-RAPS methods, which typically showed smaller MSE in all settings.

#### Supplementary Note 2: Secondary simulation studies

We consider additional simulation studies to examine the usefulness of the proposed shrinkage estimate framework in (i) improving the estimation of $\gamma$ and (ii) comparing the performance of $\beta$ estimation using both shrinkage and raw estimates as input in MR analysis. We first investigated whether the shrinkage estimate achieves better estimation performance for $\gamma$, mainly focusing on populations with smaller sample sizes (female population). Next, we use the same settings as in simulation 1 (fixed sex differences strength in $\gamma$) to conduct two additional analyses. Specifically, we first examined the situation where some variants directly affect the outcome (i.e., a horizontal pleiotropy effect), which results in some variants being invalid IVs for the analysis. Second, we compared the performance of $\beta$ estimation between two strategies: (i) applying the APM shrinkage approach only to $\gamma$ or (ii) applying it to both $\gamma$ and $\Gamma$ estimates.

We performed simulations with increasing sample sizes for the female population so that balanced sample sizes between sex groups in the exposure GWAS was also considered. This analysis aims to evaluate the performance of the shrinkage approaches in a setting where less information can be transferred between sex groups. Finally, we conducted a test of differences in the sex-specific causal effects obtained from the two-sample MR approaches (i.e., $H_0: \beta_F = \beta_M$), assessing the type 1 error rate and power of each MR method.

#### Secondary simulation study 1: Examining variant-exposure effect estimation performance

Given that the accuracy of the $\hat{\gamma}$ might highly impacts the precision of the $\beta$ estimation, especially in the presence of weak IVs in the population with smaller sample sizes, we first evaluated the estimation performance of $\hat{\gamma}$ when using shrinkage approaches ($\hat{\gamma}_{meta}, \hat{\gamma}_{APM}, \hat{\gamma}_{AW}$) and $\hat{\gamma}_{raw}$. Our rationale was that if shrinkage estimates demonstrate better performance, then more accurate and stable $\hat{\beta}$ using these shrinkage estimates can be anticipated. The results are summarized in Supplementary Figures 4, 5, and 6. When fewer variants have sex differences in $\gamma$ or when these differences are minor, the $\hat{\gamma}_{meta}$ estimate has lower MSE compared to other estimates. The $\hat{\gamma}_{APM}$ and $\hat{\gamma}_{AW}$ estimates consistently outperform the $\hat{\gamma}_{raw}$ across all simulations. When more variants having sex differences in $\gamma$ ($|D_\gamma| = 50$ and $|D_\gamma| = 90$ settings) or the correlation between $\gamma_F$ and $\gamma_M$ is not strong (correlation = 0.5 setting), $\hat{\gamma}_{APM}$ and $\hat{\gamma}_{AW}$ perform better than $\hat{\gamma}_{meta}$. All methods performed similarly in the male population, with the $\hat{\gamma}_{meta}$ showing slightly higher MSE than the others.

#### Secondary simulation study 2: Some IVs are associated with the outcome (pleiotropy)

We consider the settings with fixed sex differences in $\gamma$, while randomly selecting some variants as being pleiotropic. Specifically, we randomly selected 20 variants to have a direct effect (not mediated by the exposure) on the outcome. We generated the outcome variable by the following model:

$$O_i = 1 + \sum_{j \in K} \lambda_j \times g_{ij} + \beta \times E_i + \alpha \times U_i + \varepsilon_i$$

where $K$ denotes the set of pleiotropic variants, and $\lambda_j$ represents their direct effect sizes. The direct effects $\lambda_j$ were independently generated from a normal distribution $\lambda_j \sim N(0,0.1)$. We used the same procedure described in the Method section in the main manuscript to generate genetic variants ($g_{ij}$), exposure ($E_i$), unknown confounder ($U_i$), and random error term ($\varepsilon_i$). The variants with pleiotropic effects were the same in the two sex groups, and the direct effects were generated from the normal distribution. Both settings with and without sex differences in the exposure-outcome causal effect were considered. The female and male exposure GWAS sample sizes were 2,000 and 20,000, respectively. Results were summarized across 1000 simulation replicates.

Simulation results are displayed in Supplementary Figures 7 and 8. The conclusions align with the findings from the primary simulation studies. Overall, the shrinkage approaches have smaller MSE than estimates using $\hat{\gamma}_{raw}$. Estimates using $\hat{\gamma}_{meta}$ usually have the smallest MSE when the sex differences in $\gamma$ are not weak. Both the APM and AW methods have more robust performance in terms of MSE and 95% confidence interval coverage rate, regardless of the magnitude of sex differences in $\gamma$. When having sex differences in causal effect (Supplementary Figures 7b and 8b), using $\hat{\gamma}_{APM}$ for MR analysis significantly improves causal effect estimation performance and the 95% confidence interval coverage rate when the female population has a higher underlying causal effect. When comparing robust methods to traditional versions (robust IVW vs. IVW and robust MR-Egger vs. MR-Egger), the robust versions have much lower MSEs, demonstrating the advantages of using robust approaches when invalid IVs are present. For the male population, with less information transferable from females, the shrinkage approaches perform similarly to the use of $\hat{\gamma}_{raw}$.

#### Secondary simulation study 3: Calibrating both variant-exposure and variant-outcome effect estimates

In the primary simulation studies, we only applied the proposed semi-empirical Bayes framework for calibrating $\hat{\gamma}$. There are two main reasons for this: (i) the motivating example, the MVP sleep phenotype, serves as the exposure variable in our real data MR analysis; (ii) unstable $\hat{\gamma}$ have more severe impact on causal effect estimation than variant-outcome effect estimates ($\hat{\Gamma}$). However, the proposed framework is generalizable and can be applied to any trait's role in an MR analysis. Therefore, we examined the performance of calibrating both $\hat{\gamma}$ and $\hat{\Gamma}$ in a two-sample MR analysis. We used the fixed sex differences in $\gamma$ simulation settings to examine the results. To simplify the comparison, we focus on the raw and APM approached in variant-exposure and variant-outcome effect estimate, which already form four different combinations.

Supplementary Figures 9 and 10 summarize the results of this simulation study. For the settings with no sex differences in $\beta$ (Supplementary Figures 9a and 10a), calibrating both $\hat{\gamma}$ and $\hat{\Gamma}$ performs the best (smaller MSE) for all MR methods except the MR-Egger approach. A slight improvement in causal effect estimation performance was also observed in the male population. However, calibrating both estimates did not improve the causal effect estimation in some of the sex difference causal effect settings (Supplementary Figures 9b and 10b). Specifically, calibrating $\hat{\Gamma}$ may result in higher MSE for causal effect estimation in both female and male populations in the null female-specific causal effect setting ($\beta_F = 0, \beta_M \neq 0$). In this setting, the 95% coverage rate is much lower than the raw or only calibrating for $\hat{\gamma}$ in both sex groups. One possible reason could be that the shrinkage estimates significantly reduce the standard error estimate for both $\hat{\gamma}$ and $\hat{\Gamma}$, which may lead to an underestimation of the variance of causal effect estimate. In brief, although the proposed framework can generally be applied to calibrate any variant-phenotype association effect estimate in MR analysis, simulation results suggest its practical use is particularly evident in calibrating $\hat{\gamma}$ to address weak instrument bias issue.

#### Secondary simulation study 4: Balanced sex groups sample sizes in exposure GWAS

In this simulation study, we perform all three simulation settings in primary analysis again but increase the sample size of the female exposure GWAS population from 2,000 to 20,000, matching the male population sample size. Therefore, in these simulations, in all calibrated variant-exposure estimates we expect the weight of the female-specific estimates to increase relative to the male-specific weight, and, consequently, we expect less improvement in $\beta_F$ estimation from shrinkage approaches compared to the raw approach.

The results from this simulation study are summarized in Supplementary Figures 11 and 12. Considering MSE, estimation of $\beta_F$ while relying to $\gamma_{APM}$ performs nearly the same as the use of $\hat{\gamma}_{raw}$ in both female and male analyses. When strong sex differences in $\gamma$ exist, particularly in simulation 3, $\hat{\gamma}_{meta}$ had the highest MSE

#### Secondary simulation study 5: Sex-differences test

We evaluated the type 1 error and power of the test for sex-differences in causal effect across considered MR methods. The following test statistics is used

$$t_{\beta_F,\beta_M} = \frac{\hat{\beta}_F - \hat{\beta}_M}{\sqrt{\hat{\sigma}^2_{\beta_F} + \hat{\sigma}^2_{\beta_M}}} \qquad (1)$$

where $\hat{\beta}_F$ and $\hat{\beta}_M$ are the sex-specific causal estimates from a given MR method, and $\hat{\sigma}_{\beta_F}$, $\hat{\sigma}_{\beta_M}$ are their corresponding estimated standard errors. The test statistic in equation (1) assumes that $\hat{\beta}_F$ and $\hat{\beta}_M$ are independent, so we only include the uncalibrated approach ($\hat{\gamma}_{raw}$) in this analysis. The p-value is computed through $\chi^2_{(1)}$ distribution, and the rejection rate is calculated as the proportion of p-value $< 0.05$ across 1000 simulation replicates. In settings with no sex differences in causal effects ($\beta_F = \beta_M$), the rejection rate is the type I error rate; while the rejection rate estimates power when $\beta_F \neq \beta_M$.

Simulation results are shown in Supplementary Figure 13. Overall, the penalized and robust IVW (IVW* in the figure) has an inflated type I error rate, so a higher power of detecting sex differences is expected. In contrast, the weighted median and MR-RAPS are conservative, having type I error rates lower than 0.05. Still, MR-RAPS often has comparable or even higher power, compared to other methods, in our simulations.

Supplementary Figure 14 presents results evaluating the sex-differences test in the secondary simulation studies 4 (equivalent female-male sample size, and overall higher sample size). Overall, most of the MR methods controlled the type I error rate around the nominal threshold for the sex-differences test. The robust and penalized IVW and cML yielded a slightly inflated type I error rate, while MR-RAPs usually produced more conservative results. In terms of power, the robust and penalized IVW, cML, and MR-RAPs have higher power to detect sex differences in the causal effect compared to weighted median and penalized and robust MR-Egger.

#### Supplementary Note 3: Secondary data analyses

The sex-specific causal effect estimates using $\hat{\gamma}_{raw}$, $\hat{\gamma}_{AW}$, and $\hat{\gamma}_{meta}$, as well as comparisons between male and female-specific causal effect, are shown in Supplementary Figures 15-18.

Results based on $\hat{\gamma}_{raw}$ for IV selection are shown in Supplementary Figures 15 and 16, while those using $\hat{\gamma}_{APM}$ for IV selection are provided in Supplementary Figure 17 and 18.

#### Comparing results between MR-RAPS and MR-PRESSO

In all analyses, we set $10^{-5}$ as the p-value threshold to select IVs. This decision is due to the fact that no variants were available in the female population when a stricter p-value threshold was considered. This lower p-value threshold, however, may increase the risk of including weak IVs as well as the possibility of variants having horizontal pleiotropy effect due to the larger number of variants used as IVs. Therefore, we compared the findings from primary analyses, the causal estimation from MR-RAPS with $\hat{\gamma}_{APM}$, to MR-PRESSO, which is designed to detect the violation of horizontal pleiotropy assumption in MR analysis. The comparisons are shown in Supplementary Figures 19 and 20. Overall, the causal effect estimates are consistent between two methods, especially for those statistically significant findings identified from MR-RAPS. These results increase the reliability of our findings in primary analyses.

#### Comparing results between raw and APM-derived IV selection strategies

We compared estimated causal effects when using IV selected based on $\hat{\gamma}_{raw}$ and $\hat{\gamma}_{APM}$, as shown in Supplementary Figure 21. As expected, the resulting estimates were generally consistent with each other in the male population, as it has substantially larger sample size than the female population, so that $\hat{\gamma}_{M,APM}$ tend to be similar to $\hat{\gamma}_{M,raw}$. In contrast, using $\hat{\gamma}_{F,APM}$ identifies several additional IVs, compared to $\hat{\gamma}_{F,raw}$. Therefore, some causal effects were statistically significant only when using $\hat{\gamma}_{F,APM}$ for IV selection.

#### Results of sex-combined causal effect estimation

We estimated sex-combined causal effects in secondary analysis, using $\hat{\gamma}_{meta}$ estimates. In this case, we also used $\hat{\gamma}_{meta}$ for IV selection. The number of selected IVs are listed in Supplementary Table 2. For the outcome phenotype, we conduct sex-combined association analysis for both BMI adjustment and un-adjustment analysis, but further adjust sex variable in the analysis. The results are shown in Supplementary Figure 22. In this analysis, several causal effects between OSA and CVD-related outcomes, such as CKD, HF, and T2DM, were identified as statistically significant associations. The causal effect of OSA on CKD and on HF were also identified in male population in sex-specific analysis (primary analysis). However, for example, the significant female-specific causal effect of OSA on HTN were not identified in sex-combined analysis. This may be due to the fact that $\hat{\gamma}_{meta}$ is closer to $\hat{\gamma}_{M}$ estimates, caused by the predominance male sample size in MVP.

### Supplementary Note 4: All of Us genomics and phenotype data

We used short-read whole-genome sequencing (srWGS) data (version 7) from the All of Us (AoU) study to estimate single variant association analysis with CVD-related outcomes. Report of sequencing and quality control methods are provided in this link: https://support.researchallofus.org/hc/article_attachments/19370367115796. AoU provided genetics PCs and performed relatedness analysis. The genetic PCs information for each individual can be found in: gs://fc-aou-datasets-controlled/v7/wgs/short_read/snpindel/aux/ancestry/ancestry_preds.tsv. The relatedness kinship score can be found in gs://fc-aou-datasets-controlled/v7/wgs/short_read/snpindel/aux/relatedness/relatedness.tsv. We use the information provided in gs://fc-aou-datasets-controlled/v7/wgs/short_read/snpindel/aux/relatedness/relatedness_flagged_samples.tsv to remove related individuals in our analysis. The data were accessed on October, 2024.

#### Preprocess of clinical variables and six binary CVD-related outcomes in AoU

We first loaded basic clinical information into a Jupyter notebook, including BMI, date of birth, sex assigned at birth, and race/ethnicity for all 245,394 individuals with srWGS data. For the analysis, we focused on six binary CVD-related outcomes: atrial fibrillation (AF), coronary artery disease (CAD), cardiovascular disease (CVD), heart failure (HF), hypertension (HTN), and type 2 diabetes mellitus (T2DM). The outcome data were loaded by selecting the corresponding SNOMED and OMOP Concept IDs for each phenotype, as detailed in Supplementary Table 5. Individuals without a diagnosis of the specified outcomes were classified as controls. For individuals diagnosed with a CVD-related condition, we defined their age based on the first recorded diagnosis date and used the closest BMI measurement to that date as the BMI value for further analysis. For control individuals, we define their age as “age at 2024”, and BMI as the value from their earliest recorded measurement.

We included only individuals who self-reported their sex assigned at birth as either “Female” or “Male”. To align with summary statistics computed for the White HARE group in the MVP, we restricted our analysis to individuals self-identified as White (from race and ethnicity information), however we did not use genetic ancestry. We further limited the analysis to adults aged 18 to 95 with BMI values between 17 and 55. Additionally, individuals with documented deaths in the EHR record were excluded. After the preprocessing step, approximately 114,000 White individuals remained, consisting of 67,600 females and 46,400 males, with slight variations in sample size depending on the specific phenotype analyzed. The characteristics of the samples used in the association analysis for the AoU study are summarized in Supplementary Table 4.

#### Association analysis with CVD-related outcomes in AoU

We extracted the variants identified as IVs in the exposure GWAS (MVP) using the Hail table from the AoU srWGS data. Variants with a minor allele frequency (MAF) <1% in the AoU dataset were excluded. The resulting Hail table was then converted into a PLINK file. To obtain individual-level genetic data for association analysis, we used the R package "BEDMatrix" to import the .bed file using the function "BEDMatrix". Single-variant association analyses were performed for six binary outcomes using logistic regression, adjusting for age and 16 genetic principal components (PCs) in the BMI-unadjusted models. For BMI-adjusted analyses, the models were further adjusted for BMI, incorporating both linear and quadratic terms. The effect sizes of the variants (log(OR) with standard deviation) were extracted and used as summary statistics for the outcome GWAS. For the sex-combined analysis, we additionally adjusted for sex assigned at birth in the association models

#### Ethics statement

The All of Us research program was approved by a single IRB, the "All of Us IRB", which is charged with reviewing the protocol, informed consent, and other participant-facing materials for the All of Us Research Program. The IRB follows the regulations and guidance of the Office for Human Research Protections (https://www.hhs.gov/ohrp/index.html) for all studies, ensuring that the rights and welfare of research participants are overseen and protected uniformly. More information is provided online https://allofus.nih.gov/about/who-we-are/institutional-review-board-irb-of-all-of-us-research-program and in the All of Us design paper.

#### Acknowledgements

We gratefully acknowledge All of Us participants for their contributions and also thank the National Institutes of Health's All of Us Research Program for making available the participant data examined in this study. The All of Us Research Program is supported by the National Institutes of Health, Office of the Director: Regional Medical Centers: 1 OT2 OD026549; 1 OT2 OD026554; 1 OT2 OD026557; 1 OT2 OD026556; 1 OT2 OD026550; 1 OT2 OD 026552; 1 OT2 OD026553; 1 OT2 OD026548; 1 OT2 OD026551; 1 OT2 OD026555; IAA #: AOD 16037; Federally Qualified Health Centers: HHSN 263201600085U; Data and Research Center: 5 U2C OD023196; Biobank: 1 U24 OD023121; The Participant Center: U24 OD023176; Participant Technology Systems Center: 1 U24 OD023163; Communications and Engagement: 3 OT2 OD023205; 3 OT2 OD023206; and Community Partners: 1 OT2 OD025277; 3 OT2 OD025315; 1 OT2 OD025337; 1

OT2 OD025276. The All of Us Research Program would not be possible without the partnership of its participants.

#### Supplementary Note 5: Million Veteran Program: Core Acknowledgements for Publications

**MVP Program Office**

- Sumitra Muralidhar, Ph.D., Program Director

  US Department of Veterans Affairs, 810 Vermont Avenue NW, Washington, DC 20420

- Jennifer Moser, Ph.D., Associate Director, Scientific Programs

  US Department of Veterans Affairs, 810 Vermont Avenue NW, Washington, DC 20420

- Jennifer E. Deen, B.S., Associate Director, Cohort & Public Relations

  US Department of Veterans Affairs, 810 Vermont Avenue NW, Washington, DC 20420

**MVP Executive Committee**

- Co-Chair: Philip S. Tsao, Ph.D.

  VA Palo Alto Health Care System, 3801 Miranda Avenue, Palo Alto, CA 94304

- Co-Chair: Sumitra Muralidhar, Ph.D.

  US Department of Veterans Affairs, 810 Vermont Avenue NW, Washington, DC 20420

- J. Michael Gaziano, M.D., M.P.H.

  VA Boston Healthcare System, 150 S. Huntington Avenue, Boston, MA 02130

- Elizabeth Hauser, Ph.D.

Durham VA Medical Center, 508 Fulton Street, Durham, NC 27705

- Amy Kilbourne, Ph.D., M.P.H.

  VA HSR&D, 2215 Fuller Road, Ann Arbor, MI 48105

- Michael Matheny, M.D., M.S., M.P.H.

  VA Tennessee Valley Healthcare System, 1310 24th Ave. South, Nashville, TN 37212

- Dave Oslin, M.D.

  Philadelphia VA Medical Center, 3900 Woodland Avenue, Philadelphia, PA 19104

- Deepak Voora, MD

  Durham VA Medical Center, 508 Fulton Street, Durham, NC 27705

**MVP Co-Principal Investigators**

- J. Michael Gaziano, M.D., M.P.H.

  VA Boston Healthcare System, 150 S. Huntington Avenue, Boston, MA 02130

- Philip S. Tsao, Ph.D.

  VA Palo Alto Health Care System, 3801 Miranda Avenue, Palo Alto, CA 94304

**MVP Core Operations**

- Jessica V. Brewer, M.P.H., Director, MVP Cohort Operations

  VA Boston Healthcare System, 150 S. Huntington Avenue, Boston, MA 02130

- Mary T. Brophy M.D., M.P.H., Director, VA Central Biorepository

  VA Boston Healthcare System, 150 S. Huntington Avenue, Boston, MA 02130

- Kelly Cho, M.P.H, Ph.D., Director, MVP Phenomics

VA Boston Healthcare System, 150 S. Huntington Avenue, Boston, MA 02130

- Lori Churby, B.S., Director, MVP Regulatory Affairs

  VA Palo Alto Health Care System, 3801 Miranda Avenue, Palo Alto, CA 94304

- Scott L. DuVall, Ph.D., Director, VA Informatics and Computing Infrastructure (VINCI)

  VA Salt Lake City Health Care System, 500 Foothill Drive, Salt Lake City, UT 84148

- Saiju Pyarajan Ph.D., Director, Data and Computational Sciences

  VA Boston Healthcare System, 150 S. Huntington Avenue, Boston, MA 02130

- Robert Ringer, Pharm.D., Director, VA Albuquerque Central Biorepository

  New Mexico VA Health Care System, 1501 San Pedro Drive SE, Albuquerque, NM 87108

- Luis E. Selva, Ph.D., Director, MVP Biorepository Coordination

  VA Boston Healthcare System, 150 S. Huntington Avenue, Boston, MA 02130

- Shahpoor (Alex) Shayan, M.S., Director, MVP PRE Informatics

  VA Boston Healthcare System, 150 S. Huntington Avenue, Boston, MA 02130

- Brady Stephens, M.S., Principal Investigator, MVP Information Center

  Canandaigua VA Medical Center, 400 Fort Hill Avenue, Canandaigua, NY 14424

- Stacey B. Whitbourne, Ph.D., Director, MVP Cohort Development and Management

  VA Boston Healthcare System, 150 S. Huntington Avenue, Boston, MA 02130

**MVP Publications and Presentations Committee**

- Co-Chair: Themistocles L. Assimes, M.D., Ph. D

  VA Palo Alto Health Care System, 3801 Miranda Avenue, Palo Alto, CA 94304

- Co-Chair: Adriana Hung, M.D.; M.P.H

VA Tennessee Valley Healthcare System, 1310 24th Ave. South, Nashville, TN 37212

- Co-Chair: Henry Kranzler, M.D.

  Philadelphia VA Medical Center, 3900 Woodland Avenue, Philadelphia, PA 19104

### Supplementary Tables

#### Supplementary Table 1: Summary of primary simulation results

| | | MSE results | 95% CI coverage results |
|---|---|---|---|
| Estimation of $\beta_F$ | | | |
| | No sex differences in causal effect settings | • $\hat{\gamma}_{meta}$ performs the best when (i): there are few variants with sex differences in $\gamma$ ($\lvert D_\gamma \rvert = 10$); (ii) the correlation between $\gamma_F$ and $\gamma_M$ is strong (cor = 0.9)<br>• $\hat{\gamma}_{APM}$ outperforms $\hat{\gamma}_{raw}$ in most of the settings<br>• MR-RAPS and cML have smaller MSEs compared to other MR methods in most settings | • $\hat{\gamma}_{APM}$ has a similar coverage rate compared to $\hat{\gamma}_{raw}$<br>• $\hat{\gamma}_{APM}$ improves the coverage rate in settings with large causal effect settings ($\beta_M = \beta_F = 0.2$)<br>• The coverage rate of MR-RAPS is closest to the nominal level (95%) among all MR methods |
| | Sex differences in causal effect settings | • $\hat{\gamma}_{APM}$ improves the estimation performance over $\gamma_{raw}$ in the non-null causal effect setting ($\beta_F \neq 0$), and performs similarly to $\gamma_{raw}$ in the null causal effect setting ($\beta_F = 0$)<br>• MR-RAPS and cML have smaller MSEs compare to other MR methods in most settings | • $\hat{\gamma}_{APM}$ improved the coverage rate in the settings where the causal effect is higher in females than males ($\beta_F = 0.15;\ \beta_M = 0.1$)<br>• The coverage rate of MR-RAPS is closest to the nominal level (95%) among all MR methods |
| Estimation of $\beta_M$ | | | |
| | No sex differences in causal effect setting | • Shrinkage estimates have similar performance compared to the $\hat{\gamma}_{raw}$<br>• MR-RAPS and cML perform slightly better than other MR methods | • Shrinkage estimates have a similar coverage rate compared to $\hat{\gamma}_{raw}$<br>• The coverage rate of MR-RAPS is closest to the nominal level (95%) among all MR methods |
| | Sex differences in causal effect setting | • Shrinkage estimates have similar performance compared to the $\hat{\gamma}_{raw}$<br>• MR-RAPS and cML perform slightly better than other MR methods | • Shrinkage estimates have a similar coverage rate compared to $\hat{\gamma}_{raw}$<br>• The coverage rate of MR-RAPS is closest to the nominal level (95%) among all MR methods |

Note: For simulation 1, $D_\gamma$ denotes the set containing variants defined as sex differences in variant-exposure effect sizes.

Abbreviation: MSE: mean square error; CI: confidence interval; cor: correlation; MR: Mendelian randomization; APM: adaptive posterior mean.

#### Supplementary Table 2: Summary of primary simulation settings

| | | | | | | |
|---|---|---|---|---|---|---|
| **Simulation 1: fixed sex differences in variant-exposure effect sizes** | | | | | | |
| | No sex differences in causal effect setting | | | Sex differences in causal effect setting | | |
| exposure-outcome causal effect | $\beta_F = \beta_M = 0.1$ | | | $\beta_F = 0$ $\beta_M = 0.1$ | $\beta_F = 0.05$ $\beta_M = 0.1$ | $\beta_F = 0.15$ $\beta_M = 0.1$ |
| variant-exposure effect sizes | $\gamma_{jM} = 0.1, j = 1,2,\ldots,100$ $\gamma_{jF} = \begin{cases} 0.05, j \in D_\gamma \\ 0.1, j \notin D_\gamma \end{cases}$ | | | $\gamma_{jM} = 0.1, j = 1,2,\ldots,100$ $\gamma_{jF} = \begin{cases} 0.05, j \in D_\gamma \\ 0.1, j \notin D_\gamma \end{cases}$ | | |
| Number of variants having sex differences in variant-exposure effect sizes | $\lvert D_\gamma \rvert = 10$ | $\lvert D_\gamma \rvert = 50$ | $\lvert D_\gamma \rvert = 90$ | $\lvert D_\gamma \rvert = 50$ | | |
| **Simulation 2: random sex differences in variant-exposure effect sizes** | | | | | | |
| | No sex differences in causal effect setting | | | Sex differences in causal effect setting | | |
| exposure-outcome causal effect | $\beta_F = \beta_M = 0.1$ | | | $\beta_F = 0$ $\beta_M = 0.1$ | $\beta_F = 0.05$ $\beta_M = 0.1$ | $\beta_F = 0.15$ $\beta_M = 0.1$ |
| variant-exposure effect sizes | $\begin{pmatrix} \gamma_{jF} \\ \gamma_{jM} \end{pmatrix} \sim N_2\left(\begin{pmatrix} 0.1 \\ 0.1 \end{pmatrix}, \Sigma^2\right), j = 1,2,\ldots 100$ | | | $\begin{pmatrix} \gamma_{jF} \\ \gamma_{jM} \end{pmatrix} \sim N_2\left(\begin{pmatrix} 0.1 \\ 0.1 \end{pmatrix}, \Sigma^2\right), j = 1,2,\ldots 100$ | | |
| Correlation of $(\gamma_{jF}, \gamma_{jM})$ | 0.5 | 0.7 | 0.9 | 0.7 | | |
| **Simulation 3: Use MVP OSA GWAS summary statistics to guide variant-exposure effect sizes** | | | | | | |
| | No sex differences in causal effect setting | | | Sex differences in causal effect setting | | |
| exposure-outcome causal effect | $\beta_F = \beta_M = 0$ | $\beta_F = \beta_M = 0.1$ | $\beta_F = \beta_M = 0.2$ | $\beta_F = 0$ $\beta_M = 0.1$ | $\beta_F = 0.05$ $\beta_M = 0.1$ | $\beta_F = 0.15$ $\beta_M = 0.1$ |
| variant-exposure effect sizes | From MVP OSA bmi-unadjusted GWAS | | | From MVP OSA bmi-unadjusted GWAS | | |

Note: For simulation 1, $D_\gamma$ denotes the set containing variants defined as sex differences in variant-exposure effect sizes. For simulation 2, the sex-specific variant-exposure effect sizes were simulated from a bivariate normal distribution with the mean value set at 0.1 and the variance set at 0.01 for both sex groups.

#### Supplementary Table 3: The number of variants passing p-value thresholding and clumping procedure in each MVP sleep GWAS

| | **male selection** | **female selection** | **APM male selection** | **APM female selection** | **FE meta selection** |
|---|---|---|---|---|---|
| OSA bmi-unadjusted | 122 | 17 | 155 | 51 | 134 |
| OSA bmi-adjusted | 70 | 14 | 86 | 19 | 73 |
| Insomnia bmi-unadjusted | 23 | 15 | 30 | 12 | 22 |
| Insomnia bmi-adjusted | 21 | 15 | 29 | 13 | 22 |
| Sleepiness bmi-unadjusted | 31 | 12 | 42 | 19 | 29 |
| Sleepiness bmi-adjusted | 26 | 13 | 34 | 15 | 20 |
| Short sleep bmi-unadjusted | 41 | 12 | 55 | 16 | 47 |
| Short sleep bmi-adjusted | 41 | 13 | 47 | 18 | 43 |
| Long sleep bmi-unadjusted | 25 | 18 | 34 | 15 | 27 |
| Long sleep bmi-adjusted | 25 | 19 | 36 | 14 | 29 |

Note: The p-value threshold was $10^{-5}$.

#### Supplementary table 4: Sample sizes of MVP sleep-related GWASs

| | **BMI-unadjusted** | | **BMI-adjusted** | |
|---|---|---|---|---|
| | **female** | **male** | **female** | **male** |
| OSA | 29,795 | 380,473 | 29,325 | 372,890 |
| Insomnia | 29,793 | 380,450 | 29,323 | 372,867 |
| sleepiness | 15,346 | 212,574 | 15,033 | 206,861 |
| Short sleep | 15,262 | 210,589 | 14,951 | 204,913 |
| Long sleep | 15,262 | 210,589 | 14,951 | 204,913 |

Supplementary table 5: Characteristic of AoU individuals used in variant-outcome association analysis

| AF | Female (N=67,610) | Male (N=46,411) | Overall (N=114,021) |
|---|---|---|---|
| age | | | |
| Mean (SD) | 58.5 (16.7) | 62.0 (16.3) | 59.9 (16.6) |
| Median [Min, Max] | 61.0 [18.0, 95.0] | 65.0 [20.0, 95.0] | 63.0 [18.0, 95.0] |
| BMI | | | |
| Mean (SD) | 28.7 (7.13) | 28.8 (5.63) | 28.8 (6.56) |
| Median [Min, Max] | 27.1 [17.0, 55.0] | 27.9 [17.0, 55.0] | 27.5 [17.0, 55.0] |
| AF = No | 64,644 (95.6%) | 42,130 (90.8%) | 106,774 (93.6%) |
| AF = Yes | 2,966 (4.4%) | 4,281 (9.2%) | 7,247 (6.4%) |
| **CAD** | **Female (N=67,604)** | **Male (N=46,408)** | **Overall (N=114,012)** |
| age | | | |
| Mean (SD) | 58.6 (16.7) | 62.0 (16.3) | 59.9 (16.7) |
| Median [Min, Max] | 61.0 [20.0, 95.0] | 65.0 [20.0, 95.0] | 63.0 [20.0, 95.0] |
| BMI | | | |
| Mean (SD) | 28.7 (7.14) | 28.8 (5.63) | 28.8 (6.57) |
| Median [Min, Max] | 27.1 [17.0, 55.0] | 27.9 [17.0, 55.0] | 27.5 [17.0, 55.0] |
| CAD = No | 65,984 (97.6%) | 43,158 (93.0%) | 109,142 (95.7%) |
| CAD = Yes | 1,620 (2.4%) | 3,250 (7.0%) | 4,870 (4.3%) |
| **CKD** | **Female (N=67,573)** | **Male (N=46,391)** | **Overall (N=113,964)** |
| age | | | |
| Mean (SD) | 58.6 (16.8) | 62.3 (16.5) | 60.1 (16.8) |
| Median [Min, Max] | 61.0 [20.0, 95.0] | 66.0 [19.0, 95.0] | 63.0 [19.0, 95.0] |
| BMI | | | |
| Mean (SD) | 28.7 (7.14) | 28.8 (5.63) | 28.8 (6.57) |
| Median [Min, Max] | 27.1 [17.0, 55.0] | 27.9 [17.0, 55.0] | 27.5 [17.0, 55.0] |
| CKD = No | 65,464 (96.9%) | 43,526 (93.8%) | 108,990 (95.6%) |
| CKD = Yes | 2,109 (3.1%) | 2,865 (6.2%) | 4,974 (4.4%) |
| **HF** | **Female (N=67,582)** | **Male (N=46,380)** | **Overall (N=113,962)** |
| age | | | |
| Mean (SD) | 58.7 (16.8) | 62.5 (16.6) | 60.2 (16.8) |
| Median [Min, Max] | 61.0 [20.0, 95.0] | 66.0 [20.0, 95.0] | 63.0 [20.0, 95.0] |
| BMI | | | |
| Mean (SD) | 28.7 (7.14) | 28.8 (5.63) | 28.8 (6.57) |
| Median [Min, Max] | 27.1 [17.0, 55.0] | 27.9 [17.0, 55.0] | 27.5 [17.0, 55.0] |

| | | | |
|---|---|---|---|
| HF = No | 65,871 (97.5%) | 44,152 (95.2%) | 110,023 (96.5%) |
| HF = Yes | 1,711 (2.5%) | 2,228 (4.8%) | 3,939 (3.5%) |

| **HTN** | **Female (N=67,680)** | **Male (N=46,490)** | **Overall (N=114,170)** |
|---|---|---|---|
| age | | | |
| Mean (SD) | 55.9 (15.9) | 58.6 (15.5) | 57.0 (15.8) |
| Median [Min, Max] | 58.0 [18.0, 95.0] | 61.0 [18.0, 95.0] | 59.0 [18.0, 95.0] |
| BMI | | | |
| Mean (SD) | 28.8 (7.16) | 28.9 (5.65) | 28.8 (6.59) |
| Median [Min, Max] | 27.2 [17.0, 55.0] | 27.9 [17.0, 55.0] | 27.5 [17.0, 55.0] |
| HTN = No | 46,877 (69.3%) | 27,359 (58.8%) | 74,236 (65.0%) |
| HTN = Yes | 20,803 (30.7%) | 19,131 (41.2%) | 39,934 (35.0%) |

| **T2DM** | **Female (N=67,564)** | **Male (N=46,361)** | **Overall (N=113,925)** |
|---|---|---|---|
| age | | | |
| Mean (SD) | 58.5 (16.8) | 62.2 (16.5) | 60.0 (16.8) |
| Median [Min, Max] | 61.0 [18.0, 95.0] | 66.0 [20.0, 95.0] | 63.0 [18.0, 95.0] |
| BMI | | | |
| Mean (SD) | 28.7 (7.14) | 28.8 (5.63) | 28.8 (6.57) |
| Median [Min, Max] | 27.1 [17.0, 55.0] | 27.9 [17.0, 55.0] | 27.5 [17.0, 55.0] |
| T2DM = No | 64,872 (96.0%) | 43,043 (92.8%) | 107,915 (94.7%) |
| T2DM = Yes | 2,692 (4.0%) | 3,318 (7.2%) | 6,010 (5.3%) |

Sex was based on sex assigned at birth.
Abbreviations: AF: atrial fibrillation; CAD: coronary artery disease; CKD: chronic kidney disease; HF: heart failure; HTN: hypertension; T2DM: type 2 diabetes mellitus.

#### Supplementary Table 6: Standard concept names used to define outcome phenotypes in All of Us study

| **Outcome** | **OMOP concept ID** | **SNOMED ID** | **Number of cases** |
|---|---|---|---|
| Atrial fibrillation (AF) | 313217 | 49436004 | 7,247 |
| Coronary arteriosclerosis (CAD) | 317576 | 53741008 | 4,870 |
| Chronic Kidney disease (CKD) | 46271022 | 709044004 | 4,974 |
| Essential hypertension (HTN) | 320128 | 59621000 | 39,934 |
| Heart failure (HF) | 316139 | 84114007 | 3,939 |
| Type 2 diabetes mellitus (T2DM) | 201826 | 44054006 | 6,010 |

Note: number of cases are computed only from White individuals

### Supplementary Figures

#### Supplementary Figure 1: MSE of sex-specific causal effect estimates from primary simulation studies

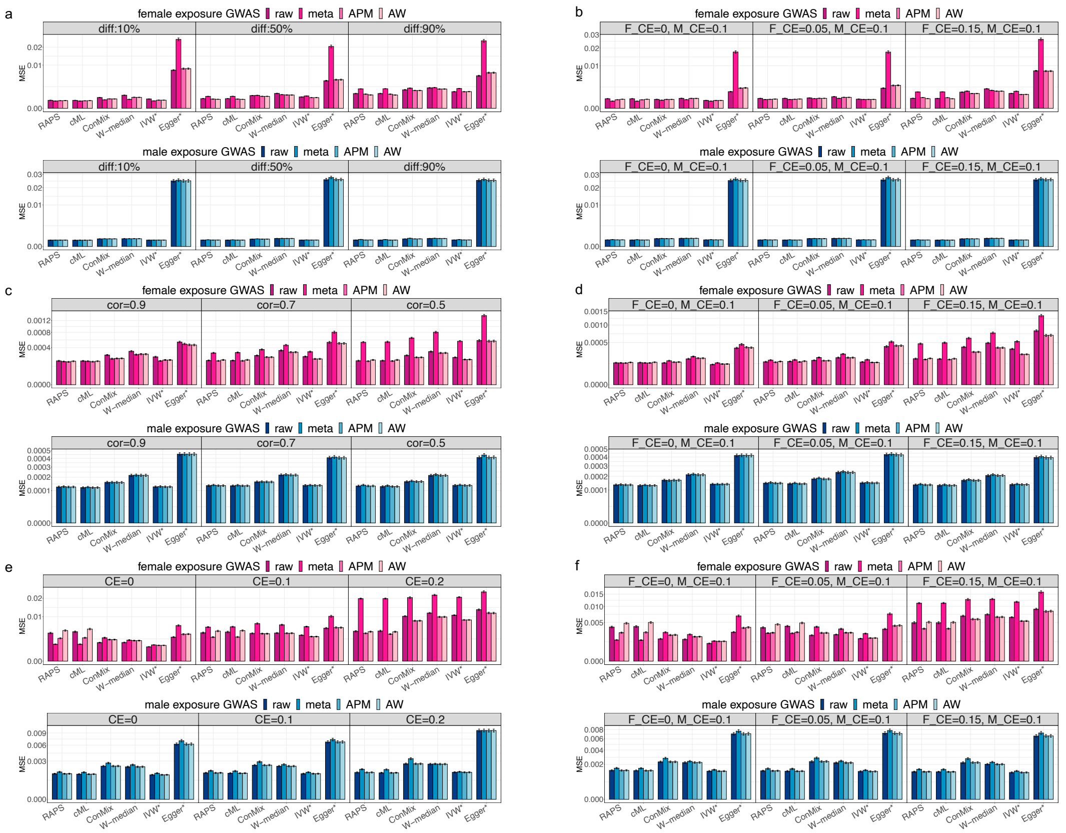

This figure shows the MSE of estimated $\beta_F$ and $\beta_M$. Female results are in pink, and male results are in blue. Two-sample MR methods are annotated as x-axis labels. Shade of bars correspond to the type of variant-exposure effect estimate (raw, meta, APM, AW), as labeled. Panels a and b provide results from simulation settings with fixed sex differences in $\gamma$, panels c and d correspond to settings with random sex differences in $\gamma$, and panels e and f correspond to settings with MVP OSA GWAS-guided sex-specific $\gamma$ effect sizes. Left panels (a, c, e) corresponds to settings with $\beta_F = \beta_M$ (CE), while the right panels (b, d, f) correspond to settings with $\beta_F \neq \beta_M$, with values denoted by F_CE and M_CE for females and males, respectively. MSEs were computed over 1000 simulation replicates. Intervals around the estimated MSE correspond to the MSE +/- one estimated standard error.

Abbreviations: MSE: mean square error; MR: Mendelian randomization; APM: adaptive posterior mean; AW: adaptive weight; diff: different level of sex differences in variant-exposure effects; Cor: correlation between female and male variant-exposure effect; CE: causal effect; W-median: weighted median; IVW*: penalized and robust IVW; Egger*: penalized and robust MR-Egger; ConMix: contaminated mixture; cML: constrained maximum likelihood; RAPS: MR-RAPS.

#### Supplementary Figure 2: 95% CI coverage rates of the true causal effect from primary simulation studies

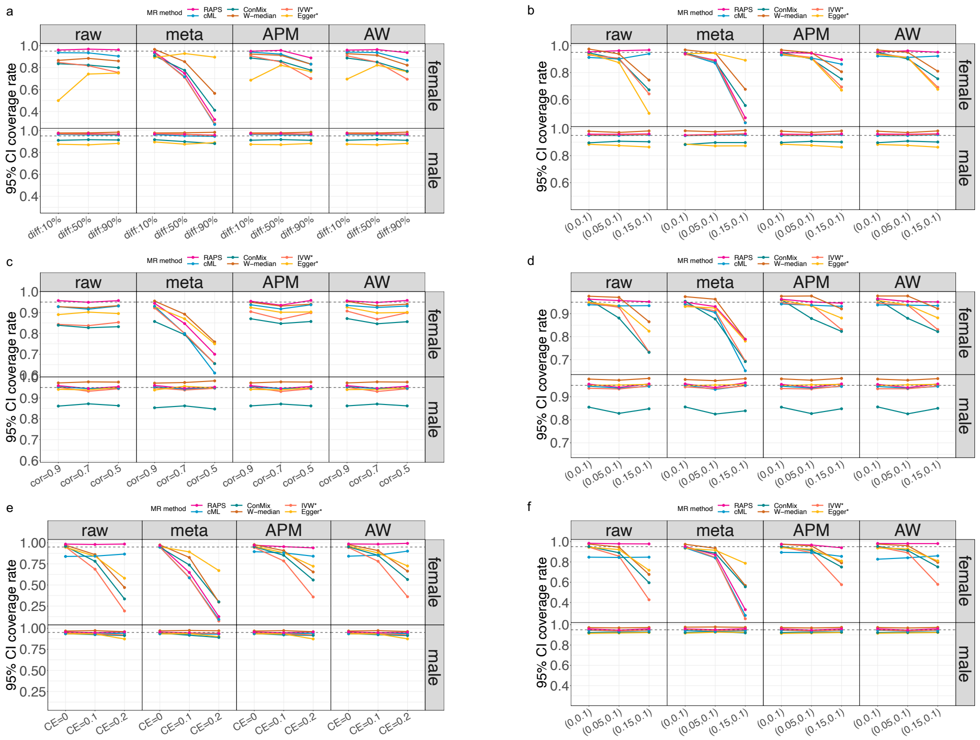

Coverage rates for the simulated true $\beta_F$ and $\beta_M$ values are shown, using 95% CI computed by each MR method across 1000 simulation replicates. In each result grid, female and male results are indicated by row labels, and methods used to estimate the variant-exposure effect sizes are indicated by column labels. Two-sample MR methods used are indicated by colors as labeled. Panels a and b provide results from simulations setting with fixed sex differences in $\gamma$, panels c and d correspond to settings with random sex differences in $\gamma$, and panels e and f correspond to settings with MVP OSA GWAS summary guided $\gamma$ effect sizes. Left panels (a, c, e) corresponds to settings with $\beta_F = \beta_M$ (CE), while the right panels (b, d, f) correspond to settings with $\beta_F \neq \beta_M$. In panel a and c, we set $\beta_F = \beta_M = 0.1$. In panel e, the underlying causal effect is shown at the bottom of the figure. Results from tests of sex differences in causal effect are provided in the right panels (b, d, f), with the underlying sex-specific causal effect shown in parentheses below (x-axis labels) in the form $(\beta_F, \beta_M)$.

Abbreviations: CI: confidence interval; MR: Mendelian randomization; APM: adaptive posterior mean; AW: adaptive weight; diff: different level of sex differences in exposure effects; Cor: correlation between female and male exposure effect; CE: causal effect; W-median: weighted median; IVW*: penalized and robust IVW; Egger*: penalized and robust MR-Egger; ConMix: contaminated mixture; cML: constrained maximum likelihood; RAPS: MR-RAPS.

#### Supplementary Figure 3: Boxplots of female-specific exposure-outcome causal effect estimates from primary simulation studies

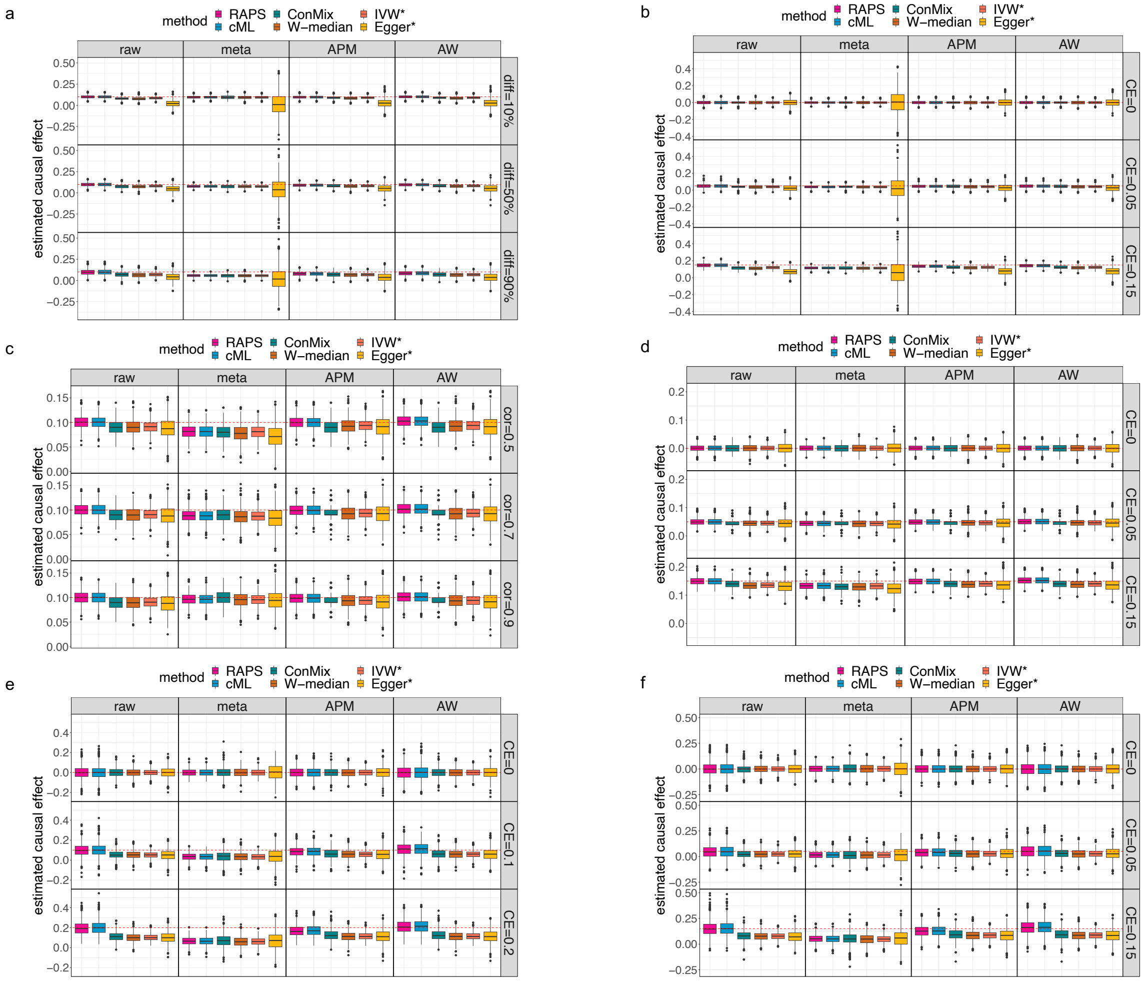

This figure shows the female-specific exposure-outcome causal effect estimates using boxplots from each MR method. Each boxplot represents $\beta_F$ estimates across 1,000 simulation replicates. We evaluated six two-sample MR methods for estimating the causal effect: W-median, IVW*, Egger*, ConMix, cML, and RAPS. In each panel, columns are labeled according to the type of variant-exposure effect estimate used. Panels a and b illustrate results from simulation settings with fixed sex differences in $\gamma$. Panels c and d show results from settings with random sex differences in $\gamma$. Panels e and f display the results from settings using MVP OSA GWAS summary statistics to guide sex-specific $\gamma$ effect sizes. The left panels (a, c, e) present the results of no sex differences in causal effect settings ($\beta_F = \beta_M$), while the right panels (b, d, f) show results from settings with $\beta_F \neq \beta_M$. In the latter settings, $\beta_M = 0.1$, and simulated $\beta_F$ values are indicated in the legend on the right-hand side of each panel (CE). The horizontal dashed redline highlight the simulated, true, $\beta_F$.

Abbreviations: MR: Mendelian randomization; APM: adaptive posterior mean; AW: adaptive weight; diff: different level of sex differences in variant-exposure effects; Cor: correlation between female and male variant-exposure effects; CE: causal effect; W-median: weighted median; IVW*: penalized and robust IVW; Egger*:

penalized and robust MR-Egger; ConMix: contaminated mixture; cML: constrained maximum likelihood; RAPS: MR-RAPS.

#### Supplementary Figure 4: Boxplots for MSE of variant-exposure effect estimates in secondary simulation studies 1

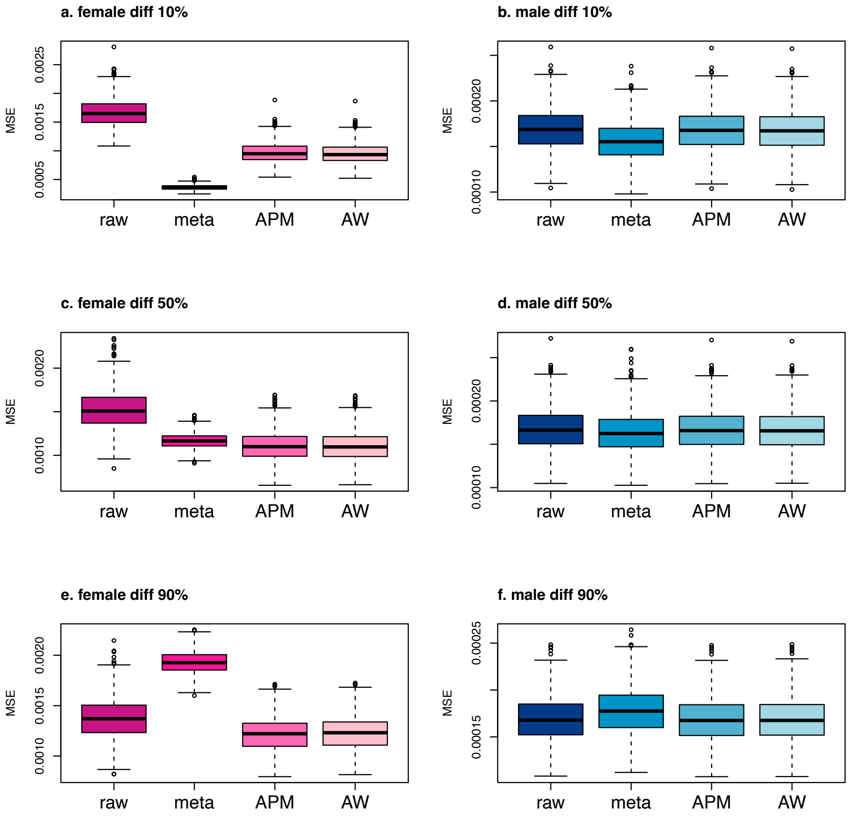

The figure shows the estimation performance of the variant-exposure ($\gamma$) effect estimate using shrinkage approaches (meta, APM, and AW) and the uncalibrated (raw) estimates from secondary simulation studies 1. For each simulation replicate, MSE values were computed averaging all 100 variants, while the boxplots use the resulting MSEs across the 1000 simulation replicates. Results of the female-specific and male-specific variant-exposure estimate are shown on the left and right panels, respectively.

Abbreviations: MSE: mean square error; APM: adaptive posterior mean; AW: adaptive weight; diff: different level of sex differences in variant-exposure effects.

#### Supplementary Figure 5: Boxplots of MSE of variant-exposure effect estimates in secondary simulation study 2

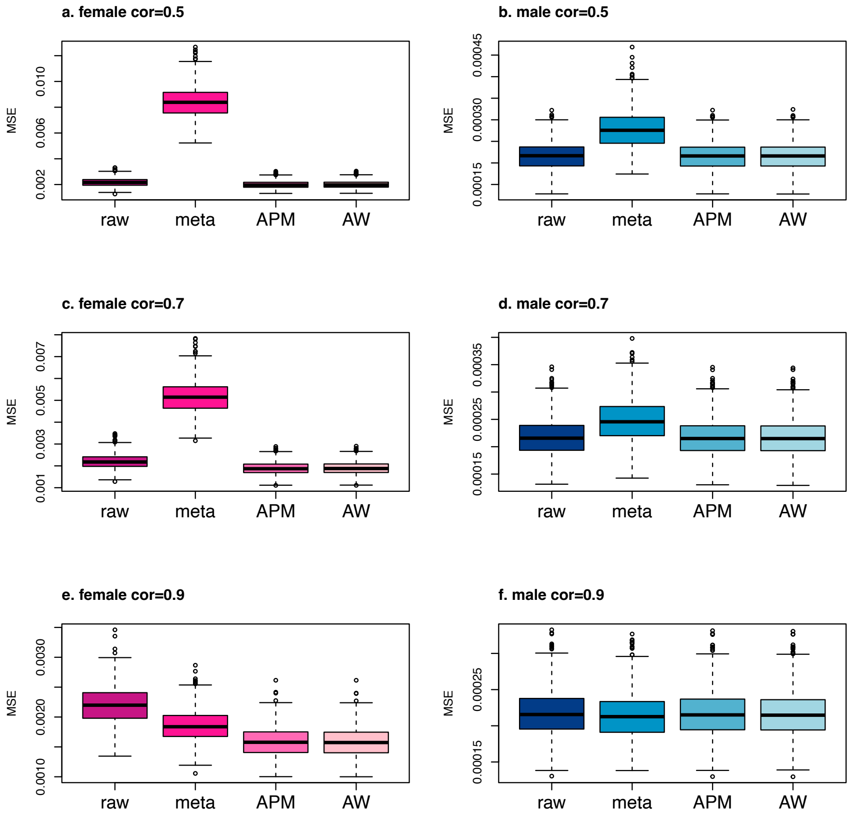

This figure shows the estimation performance of the variant-exposure effects using shrinkage approaches (meta, APM, and AW) and the uncalibrated (raw) estimates in secondary simulation studies 2. The correlation between $\gamma_F$ and $\gamma_M$ are shown in the subtitle of each panel (e.g., cor = 0.5, 0.7, 0.9). For each simulation replicate, MSE values were computed averaging all 100 variants, while the boxplots use the resulting MSEs across the 1000 simulation replicates. Results of the female-specific and male-specific variant-exposure estimate are shown on the left and right panels, respectively.

Abbreviations: MSE: mean square error; APM: adaptive posterior mean; AW: adaptive weight; cor: correlation between $\gamma_F$ and $\gamma_M$.

#### Supplementary Figure 6: Boxplot for MSE of variant-exposure effect estimates in secondary simulation study 3

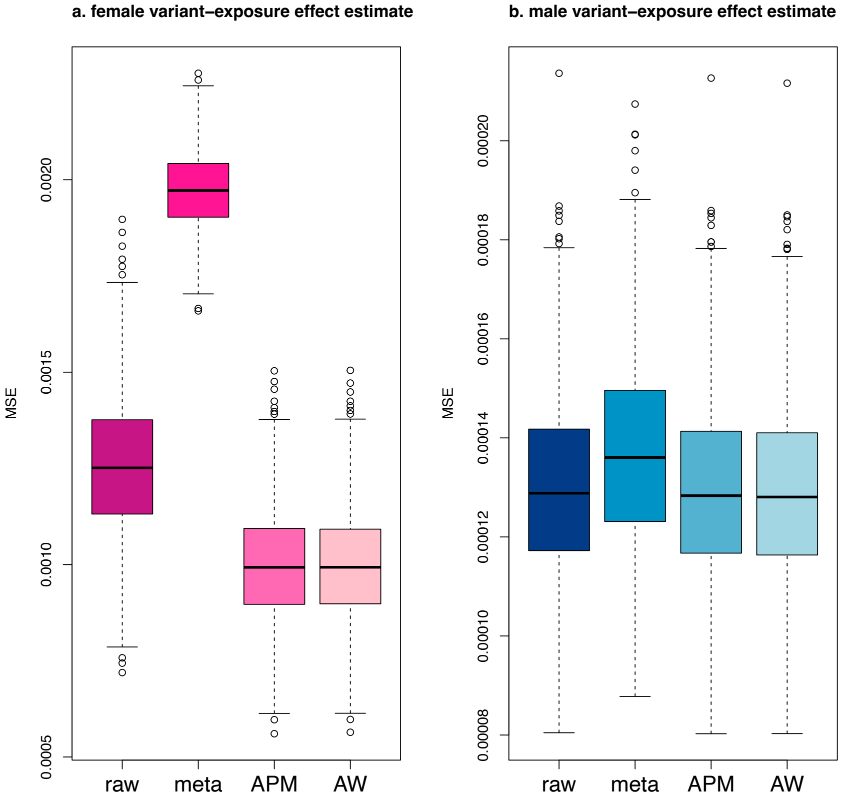

MSE of variant-exposure effect estimate using shrinkage approaches (meta, APM, and AW) and the uncalibrated (raw) estimates from secondary simulations 3. For each simulation replicate, MSE values were computed averaging all 100 variants, while the boxplots use the resulting MSEs across the 1000 simulation replicates. The results of the female-specific variant-exposure estimate are shown on the left panel, and the male-specific variant-exposure estimate results are shown on the right panel.

Abbreviations: MSE: mean square error; APM: adaptive posterior mean; AW: adaptive weight.

#### Supplementary Figure 7: MSE of sex-specific causal effect estimates from simulations with pleiotropic variants

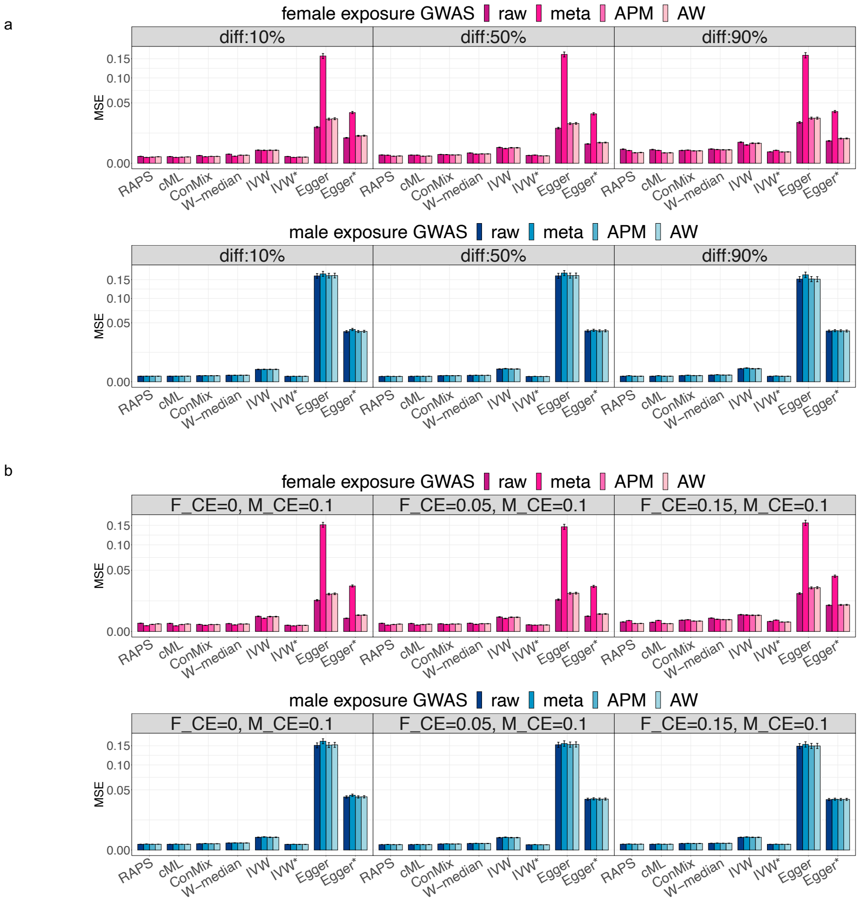

This figure shows the MSE of sex-specific causal effect estimates. Female results are in color pink, and male results are in color blue. We considered eight two-sample MR methods to estimate the causal effect: W-median, IVW, IVW*, MR-Egger, Egger*, ConMix, cML, and RAPS. The color level of the bars corresponds to type of variant-exposure estimate used. The results from simulation settings with no sex differences in causal effect ($\beta_F = \beta_M = 0.1$) are shown in panel a, and MSEs from settings with sex differences in causal effect are shown in panel b. in panel b, the $\beta_F$ and $\beta_M$ values are labeled as F_CE and M_CE, representing the causal effects for females and males, respectively. MSEs were computed over 1000 simulation replicates. Intervals around the estimated MSE correspond to the MSE +/- one estimated standard error.

Abbreviations: MSE: mean square error; MR: Mendelian randomization; APM: adaptive posterior mean; AW: adaptive weight; diff: different level of sex differences in exposure effects; CE: causal effect; W-median: weighted median; IVW: inverse-variance weighted; IVW*: penalized and robust IVW; Egger: MR-Egger; Egger*: penalized and robust MR-Egger; ConMix: contaminated mixture; cML: constrained maximum likelihood; RAPS: MR-RAPS.

#### Supplementary Figure 8: Confidence interval coverage rate of the true causal effect from pleiotropy effect simulation studies

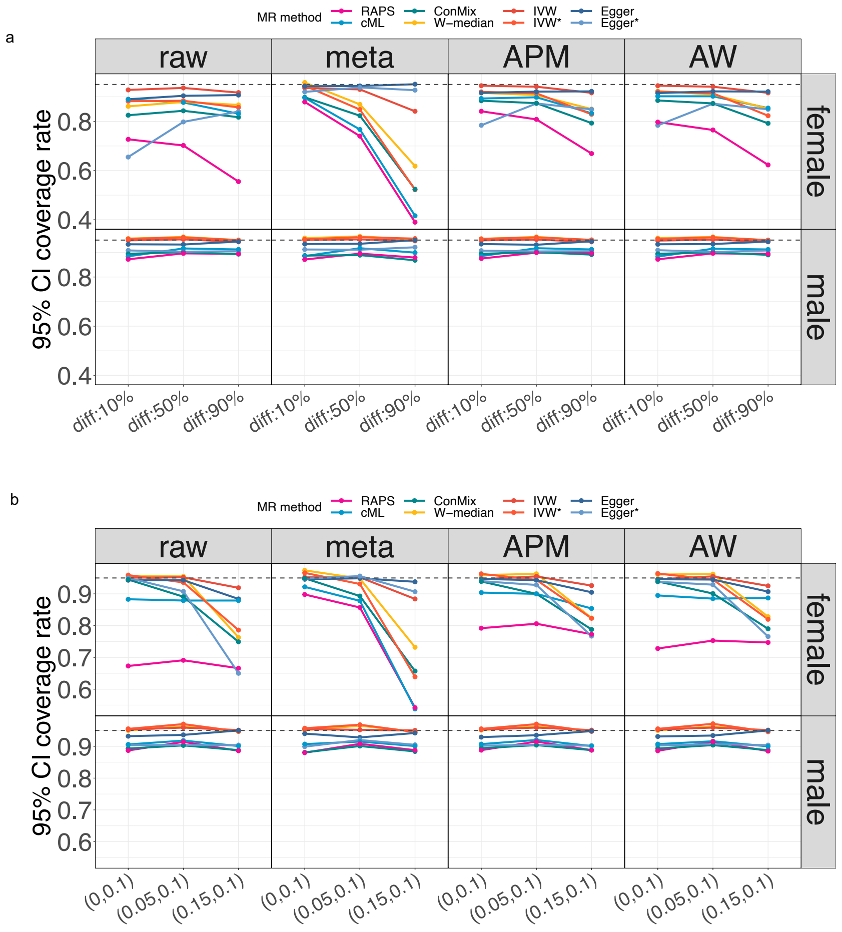

Coverage rates of the underlying true sex-specific causal effect using 95% CI computed by each MR method across 1000 simulation replicates. We considered eight two-sample MR methods to estimate the causal effect: W-median, IVW, IVW*, MR-Egger, Egger*, ConMix, cML, and RAPS. Column titles describe which variant-exposure effect estimate approach was used. Results from simulation settings with $\beta_F = \beta_M = 0.1$ are illustrated in panel a. Panel b provides results from simulations with $\beta_F \neq \beta_M$, with the underlying sex-specific causal effect shown in the parentheses below (x-axis) in the form $(\beta_F, \beta_M)$.

Abbreviations: CI: confidence interval; MR: Mendelian randomization; APM: adaptive posterior mean; AW: adaptive weight; diff: different level of sex differences in exposure effects; W-median: weighted median; IVW: inverse-variance weighted; IVW*: penalized and robust IVW; Egger: MR-Egger; Egger*: penalized and robust MR-Egger; ConMix: contaminated mixture; cML: constrained maximum likelihood; RAPS: MR-RAPS.

#### Supplementary Figure 9: MSE of sex-specific causal effect estimates from simulations calibrating both variant-exposure and outcome analyses

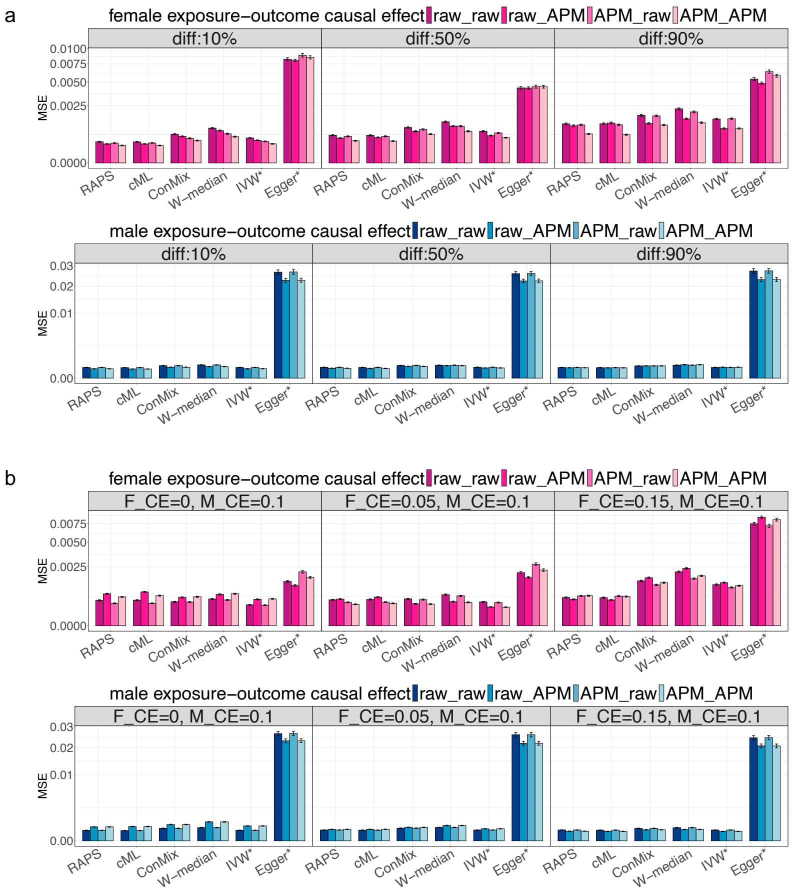

This figure shows the MSE of sex-specific causal effect estimates in secondary simulation study 3. The fixed sex differences in $\gamma$ setting is applied in this analysis. Female results are in color pink, and male results are in color blue. We considered six two-sample MR methods to estimate the causal effect: W-median, IVW*, Egger*, ConMix, cML, and RAPS. Four combinations of variant-phenotype effect estimates are considered: no calibration for both variant-exposure and variant-outcome effect estimate (raw_raw), calibration only for variant-outcome effect estimate using APM (raw_APM), calibration only for variant-exposure effect estimate using APM (APM_raw), and calibration for both variant-exposure and variant-outcome effect estimate (APM_APM). The results of no sex differences in causal effect settings ($\beta_F = \beta_M = 0.1$) are shown in panel a, and MSEs of sex differences in causal effect settings are shown in panel b, with $\beta_F$ and $\beta_M$ values denoted by F_CE and M_CE for females and males, respectively. MSEs were computed over 1000 simulation replicates. Intervals around the estimated MSE correspond to the MSE +/- one estimated standard error.

Abbreviations: MSE: mean square error; MR: Mendelian randomization; APM: adaptive posterior mean; AW: adaptive weight; diff: different level of sex differences in exposure effects; CE: causal effect; W-median: weighted median; IVW: inverse-variance weighted; IVW*: penalized and robust IVW; Egger: MR-Egger; Egger*: penalized and robust MR-Egger; ConMix: contaminated mixture; cML: constrained maximum likelihood; RAPS: MR-RAPS.

#### Supplementary Figure 10: Confidence interval coverage rate of the true causal effect in simulations calibrating both variant-exposure and outcome analyses

a

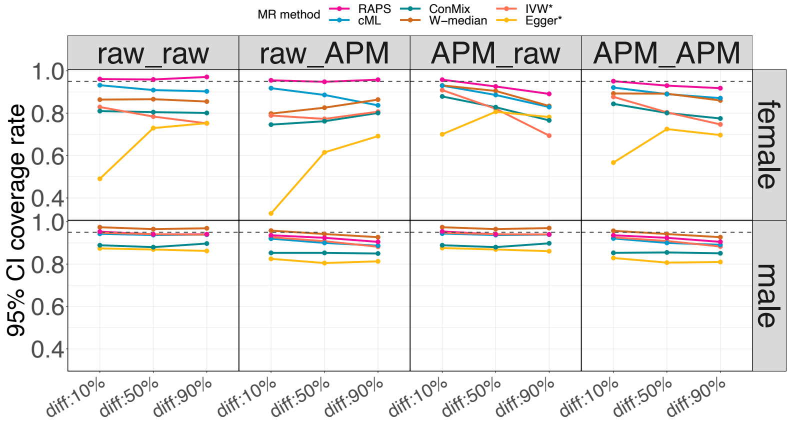

b

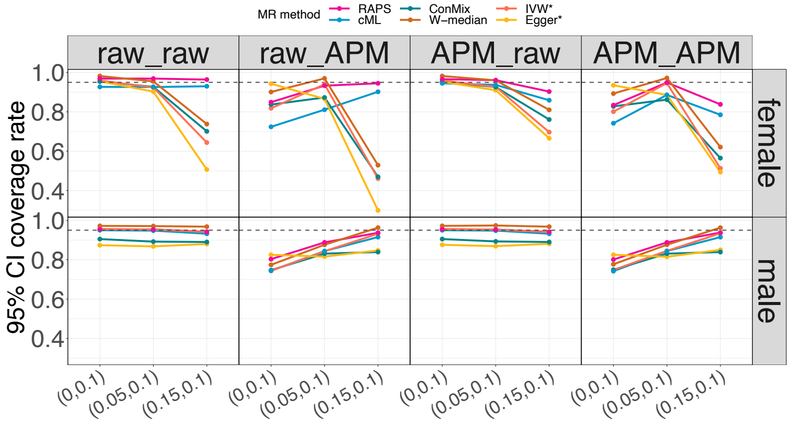

The figure provides coverage rates of the underlying true sex-specific causal effect using 95% CI computed by each MR method across 1000 simulation replicates. Female results are shown at the top, and male results are shown at the bottom. We considered six two-sample MR methods to estimate causal effect: W-median, IVW*, Egger*, ConMix, cML, and RAPS. Four combinations of variant-phenotype effect estimates are considered: no calibration for both variant-exposure and variant-outcome effect estimate (raw_raw), calibration only for variant-outcome effect estimate using APM (raw_APM), calibration only for variant-exposure effect estimate using APM (APM_raw), and calibration for both variant-exposure and variant-outcome effect estimate (APM_APM). The results of the same causal effect settings are illustrated in panel a, in which the causal effects are set as 0.1 for both females and males. Panel b provides results from simulations with $\beta_F \neq \beta_M$, with the underlying sex-specific causal effect shown in parentheses below (x-axis) in the form $(\beta_F, \beta_M)$.

Abbreviations: CI: confidence interval; MR: Mendelian randomization; APM: adaptive posterior mean; AW: adaptive weight; diff: different level of sex differences in exposure effects; W-median: weighted median; IVW: inverse-variance weighted; IVW*: penalized and robust IVW; Egger: MR-Egger; Egger*: penalized and robust MR-Egger; ConMix: contaminated mixture; cML: constrained maximum likelihood; RAPS: MR-RAPS.

### Supplementary Figure 11: MSE of sex-specific causal effect estimates from secondary simulation 4 (equivalent female-male sample size)

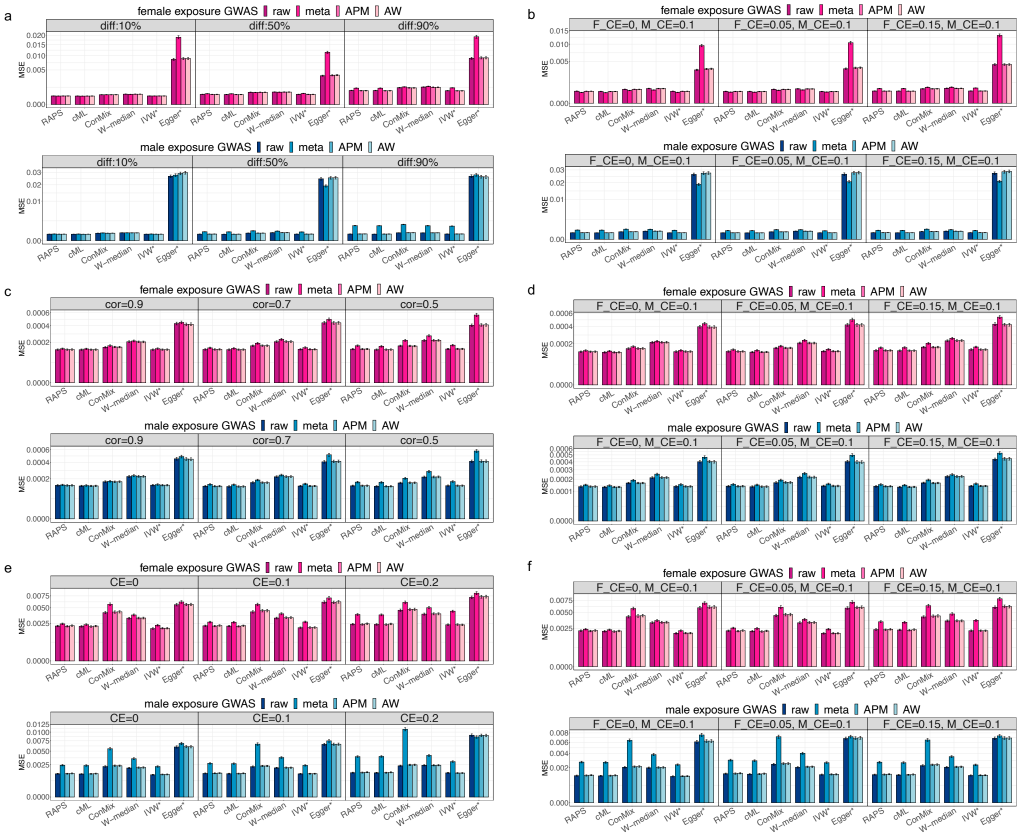

This figure shows the MSE of estimated $\beta_F$ and $\beta_M$ in secondary simulation 4. Female results are in pink, and male results are in blue. Two-sample MR methods are annotated as x-axis labels. Shade of bars correspond to the type of variant-exposure effect estimate (raw, meta, APM, AW), as labeled. Panels a and b provide results from simulation settings with fixed sex differences in $\gamma$, panels c and d correspond to settings with random sex differences in $\gamma$, and panels e and f correspond to settings with MVP OSA GWAS-guided sex-specific $\gamma$ effect sizes. Left panels (a, c, e) corresponds to settings with $\beta_F = \beta_M$ (CE), while the right panels (b, d, f) correspond to settings with $\beta_F \neq \beta_M$, with values denoted by F_CE and M_CE for females and males, respectively. MSEs were computed over 1000 simulation replicates. Intervals around the estimated MSE correspond to the MSE +/- one estimated standard error.

Abbreviations: MSE: mean square error; MR: Mendelian randomization; APM: adaptive posterior mean; AW: adaptive weight; diff: different level of sex differences in variant-exposure effects; Cor: correlation between female and male variant-exposure effect; CE: causal effect; W-median: weighted median; IVW*: penalized and robust IVW; Egger*: penalized and robust MR-Egger; ConMix: contaminated mixture; cML: constrained maximum likelihood; RAPS: MR-RAPS.

#### Supplementary Figure 12: Confidence interval coverage rate of the true causal effect from secondary simulation 4 (equivalent female-male sample size)

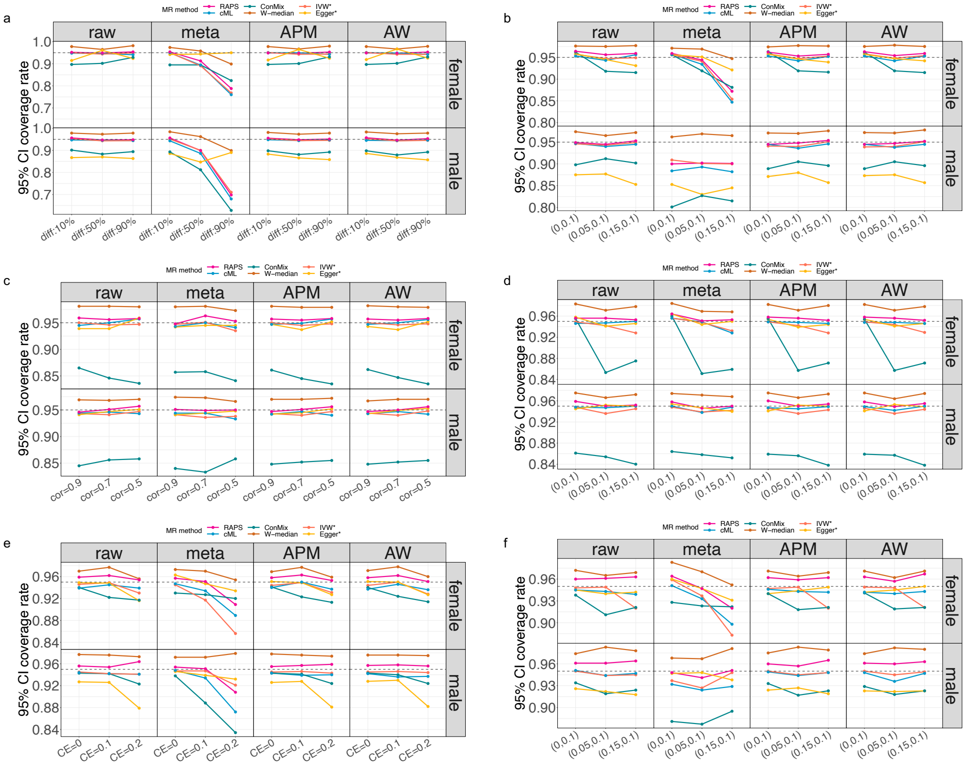

Coverage rates for the simulated true $\beta_F$ and $\beta_M$ values are shown, using 95% CI computed by each MR method across 1000 simulation replicates. In each result grid, female and male results are indicated by row labels, and methods used to estimate the variant-exposure effect sizes are indicated by column labels. Two-sample MR methods used are indicated by colors as labeled. Panels a and b provide results from simulations setting with fixed sex differences in $\gamma$, panels c and d correspond to settings with random sex differences in $\gamma$, and panels e and f correspond to settings with MVP OSA GWAS summary guided $\gamma$ effect sizes. Left panels (a, c, e) corresponds to settings with $\beta_F = \beta_M$ (CE), while the right panels (b, d, f) correspond to settings with $\beta_F \neq \beta_M$. In panel a and c, we set $\beta_F = \beta_M = 0.1$. In panel e, the underlying causal effect is shown at the bottom of the figure. Results from tests of sex differences in causal effect are provided in the right panels (b, d, f), with the underlying sex-specific causal effect shown in parentheses below (x-axis labels) in the form $(\beta_F, \beta_M)$.

Abbreviations: CI: confidence interval; MR: Mendelian randomization; APM: adaptive posterior mean; AW: adaptive weight; diff: different level of sex differences in exposure effects; Cor: correlation between female and male exposure effect; CE: causal effect; W-median: weighted median; IVW*: penalized and robust IVW; Egger*: penalized and robust MR-Egger; ConMix: contaminated mixture; cML: constrained maximum likelihood; RAPS: MR-RAPS.

#### Supplementary Figure 13: Rejection rates of sex-differences test from primary simulation studies

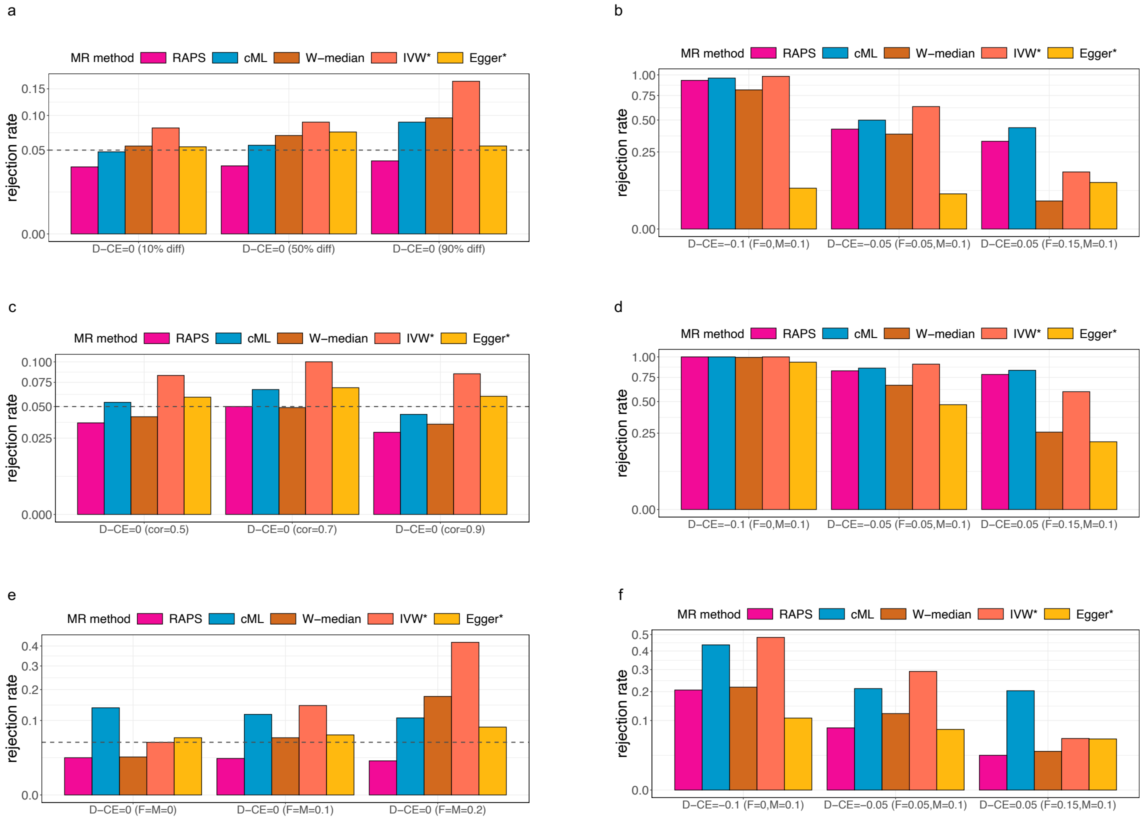

The figure shows the rejection rates of the sex-differences test computed by each MR method across 1000 simulation replicates in primary simulation studies. The left panels (a, c, e) provide results from settings with no sex differences in the causal effect (i.e., measuring type 1 error), while the panels on the right (b, d, f) provide results from settings with sex differences in the causal effects (i.e., measuring power). Both female and male causal effects are set as 0.1 in panel a and c. In panel e, the underlying causal effect is shown in parentheses below (x-axis labels) in the form (F ($\beta_F$) = M ($\beta_M$) = value). The values of female and male-specific causal effects in sex-differences settings are shown at the bottom of each sub-panel (sub-panel b, d, and f) and are specified as D-CE=value. The horizontal dashed line at 0.05 in sub-panel a, c, and e indicates the desired type 1 error rate. We used five two-sample MR methods to estimate sex-specific causal effect: W-median, IVW*, Egger*, cML, and RAPS. The contaminated mixture approach was not included in this analysis because the algorithm does output the standard error of the estimated causal effect. All methods used the raw variant-exposure effect estimate to conduct sex differences tests.

Abbreviations: D-CE: differences in causal effect; CE: causal effect; MR: Mendelian randomization; diff: different level of sex differences in exposure effects; Cor: correlation between female and male exposure effect; W-median: weighted median; IVW*: penalized and robust IVW; Egger*: penalized and robust MR-Egger; cML: constrained maximum likelihood; RAPS: MR-RAPS; F: female; M: Male.

#### Supplementary Figure 14: Rejection rates of sex-differences test from secondary simulation 4 (equivalent female-male sample size)

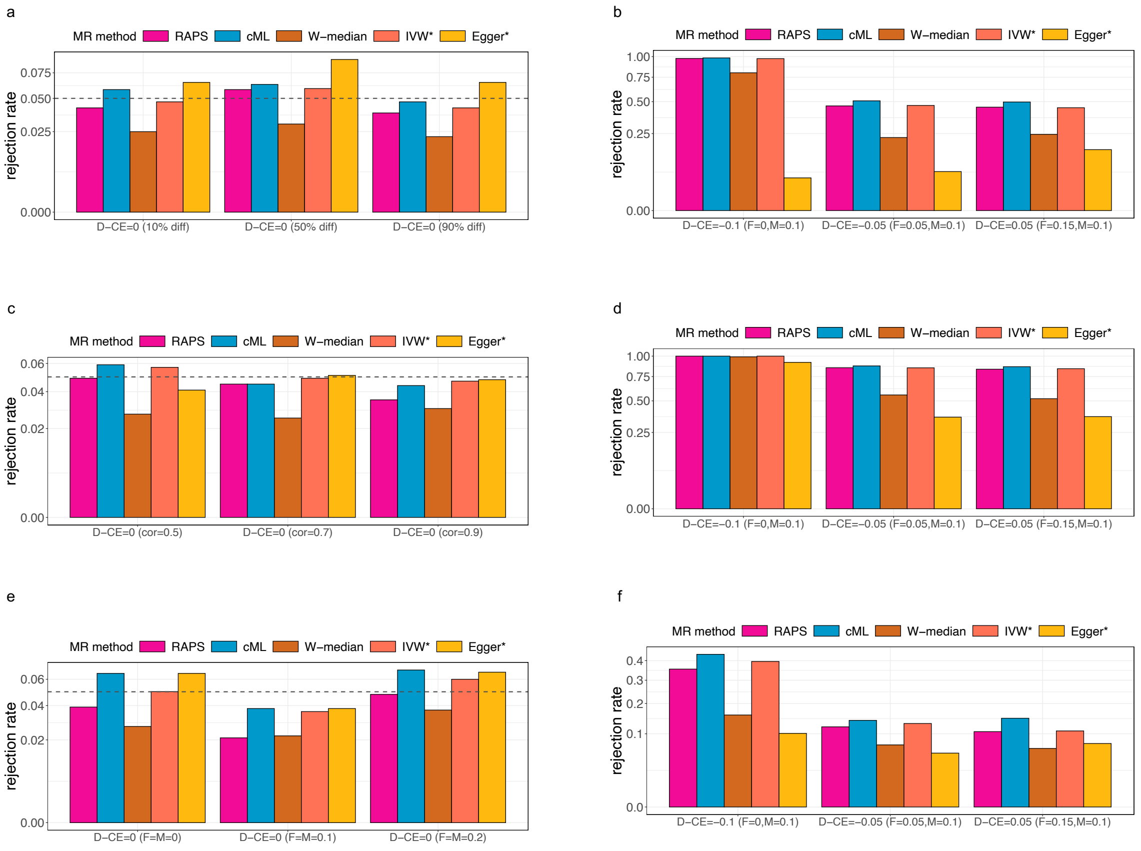

The figure shows the rejection rates of the sex-differences test computed by each MR method across 1000 simulation replicates in increasing female sample size settings. The left panels (a, c, e) provide results from settings with no sex differences in the causal effect (i.e., measuring type 1 error), while the panels on the right (b, d, f) provide results from settings with sex differences in the causal effects (i.e., measuring power). Both female and male causal effects are set as 0.1 in panel a and c. In panel e, the underlying causal effect is shown in parentheses below (x-axis labels) in the form (F ($\beta_F$) = M ($\beta_M$) = value). The values of female and male-specific causal effects in sex-differences settings are shown at the bottom of each sub-panel (sub-panel b, d, and f) and are specified as D-CE=value. The horizontal dashed line at 0.05 in sub-panel a, c, and e indicates the desired type 1 error rate. We used five two-sample MR methods to estimate sex-specific causal effect: W-median, IVW*, Egger*, cML, and RAPS. The contaminated mixture approach was not included in this analysis because the algorithm does output the standard error of the estimated causal effect. All methods used the raw variant-exposure effect estimate to conduct sex differences tests.

Abbreviations: D-CE: differences in causal effect; CE: causal effect; MR: Mendelian randomization; diff: different level of sex differences in exposure effects; Cor: correlation between female and male exposure effect; W-median: weighted median; IVW*: penalized and robust IVW; Egger*: penalized and robust MR-Egger; cML: constrained maximum likelihood; RAPS: MR-RAPS; F: female; M: Male.

#### Supplementary Figure 15: Results from sex-specific causal effect estimation using BMI-unadjusted sleep GWASs

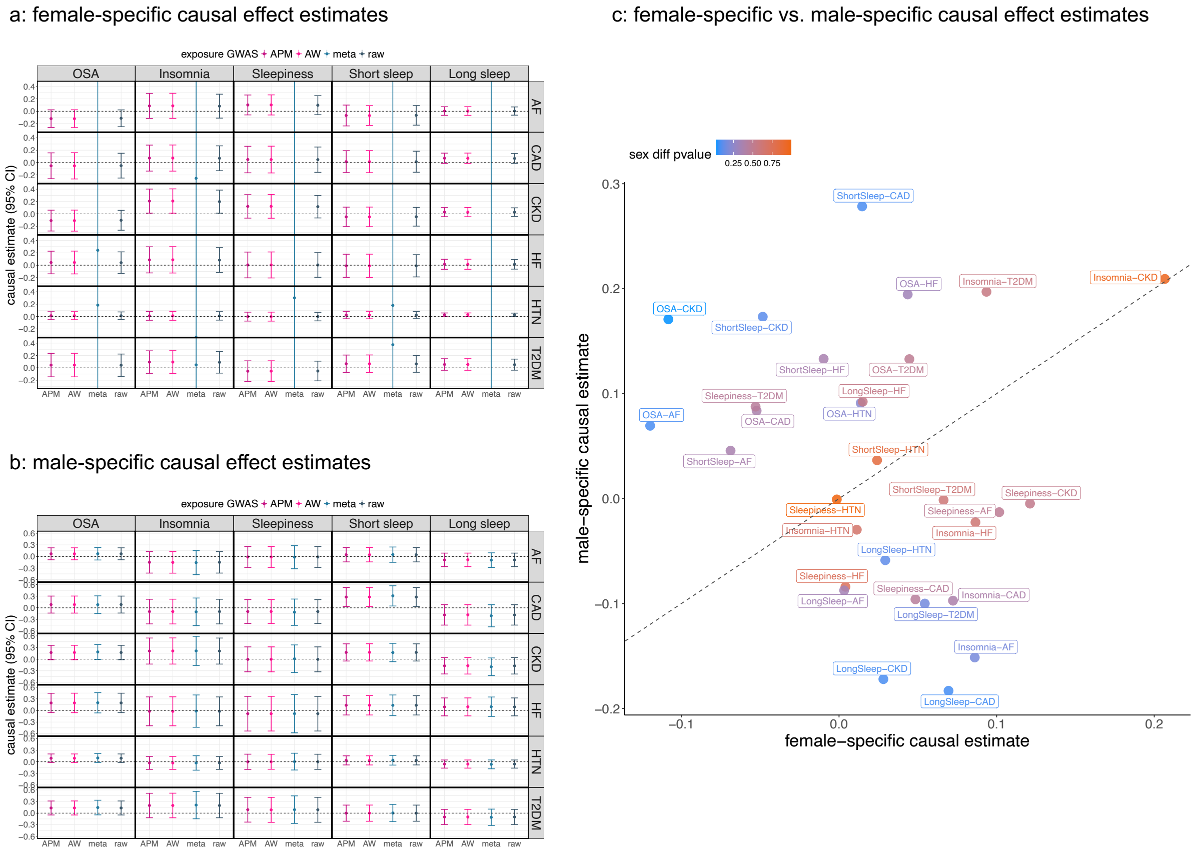

Panels a (female-specific) and b (male-specific) provide sex-specific causal effect ($\beta_F, \beta_M$) estimates with the corresponding 95% CIs. The causal effects were estimated using MR-RAPS and are displayed on the log(OR) scale. Horizontal dashed lines indicate the null value. These analyses used summary statistics from BMI-unadjusted sleep GWASs. IVs were selected based on raw estimates ($\hat{\gamma}_{raw}$). The columns represent the exposure variables, while the rows show the outcome variables. Due to the large standard errors of the $\hat{\beta}_{F,meta}$ estimates (estimates that used $\hat{\gamma}_{meta}$), the corresponding CIs were truncated. Panel c compares estimated $\beta_F$ (x-axis) and $\beta_M$ (y-axis) obtained when using APM estimates ($\hat{\gamma}_{APM}$). The color of each point in panel c is proportional to the p-value of the sex-differences test.

Abbreviations: CI: confidence interval; IV: instrumental variable; OR: odds ratio; MR: Mendelian randomization; MR-RAPS: MR using robust adjusted profile score method; APM: adaptive posterior mean; AW: adaptive weight; meta: fixed-effect meta estimates; sex diff p-value: p-value of sex differences test; OSA: obstructive sleep apnea; sleepiness: excessive daytime sleepiness; AF: atrial fibrillation; CAD: coronary artery disease; CKD: chronic kidney disease; HF: heart failure; HTN: hypertension; T2DM: type 2 diabetes mellitus.

#### Supplementary Figure 16: Results from sex-specific causal effect estimation using BMI-adjusted sleep GWASs

a: female-specific causal effect estimates

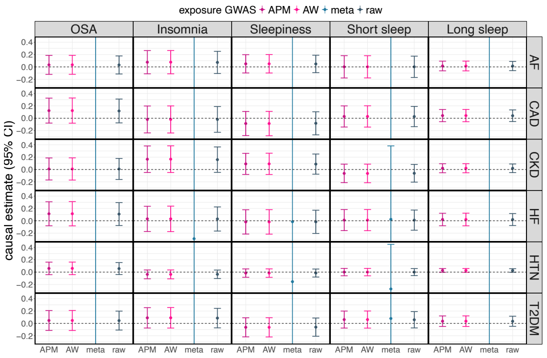

c: female-specific vs. male-specific causal effect estimates

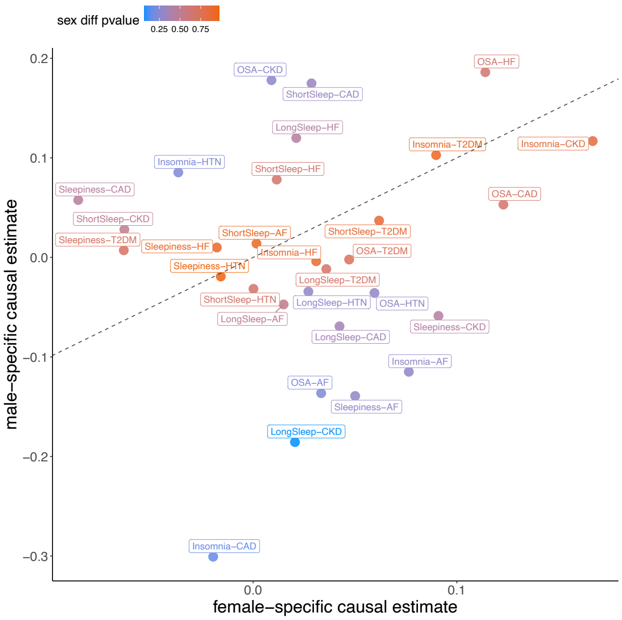

b: male-specific causal effect estimates

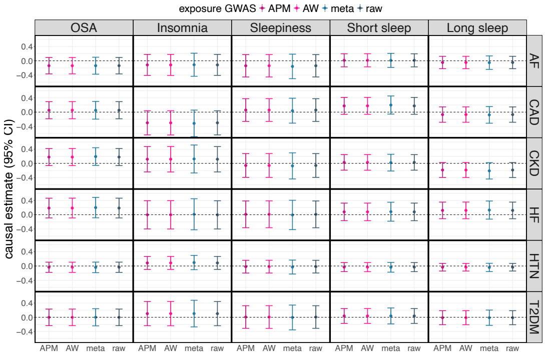

Panels a (female-specific) and b (male-specific) provide sex-specific causal effect ($\beta_F, \beta_M$) estimates with the corresponding 95% CIs. The causal effects were estimated using MR-RAPS and are displayed on the log(OR) scale. Horizontal dashed lines indicate the null value. These analyses used summary statistics from BMI-adjusted sleep GWASs. IVs were selected based on raw estimates ($\hat{\gamma}_{raw}$). The columns represent the exposure variables, while the rows show the outcome variables. Due to the large standard errors of the $\hat{\beta}_{F,meta}$ estimates (estimates that used $\hat{\gamma}_{meta}$), the corresponding CIs were truncated. Panel c compares estimated $\beta_F$ (x-axis) and $\beta_M$ (y-axis) obtained when using APM estimates ($\hat{\gamma}_{APM}$). The color of each point in panel c is proportional to the p-value of the sex-differences test.

Abbreviations: CI: confidence interval; IV: instrumental variable; OR: odds ratio; MR: Mendelian randomization; MR-RAPS: MR using robust adjusted profile score method; APM: adaptive posterior mean; AW: adaptive weight; meta: fixed-effect meta estimates; sex diff p-value: p-value of sex differences test; OSA: obstructive sleep apnea; sleepiness: excessive daytime sleepiness; AF: atrial fibrillation; CAD: coronary artery disease; CKD: chronic kidney disease; HF: heart failure; HTN: hypertension; T2DM: type 2 diabetes mellitus.

#### Supplementary Figure 17: Results from sex-specific causal effect estimation using variants selected by APM estimates (BMI-unadjusted sleep GWASs)

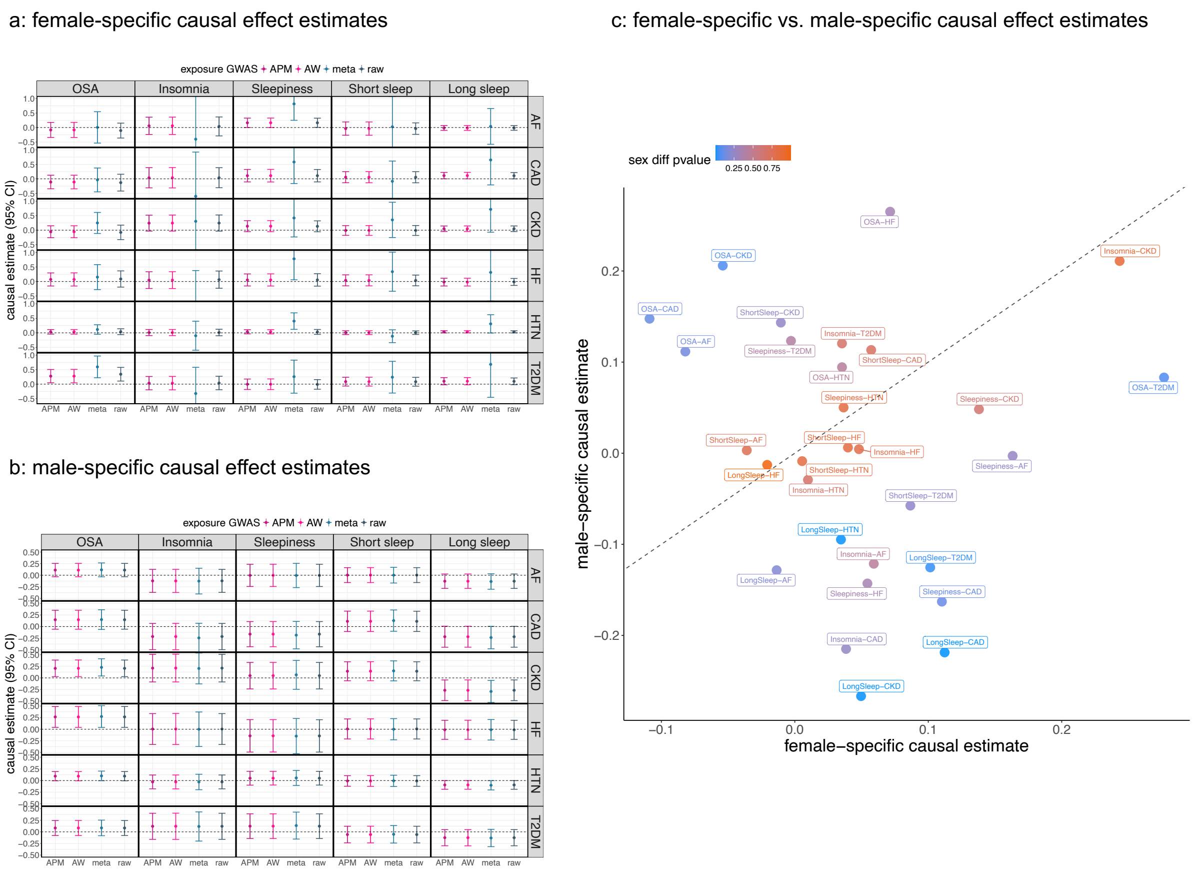

Panels a (female-specific) and b (male-specific) provide sex-specific causal effect ($\beta_F, \beta_M$) estimates with the corresponding 95% CIs. The causal effects were estimated using MR-RAPS and are displayed on the log(OR) scale. Horizontal dashed lines indicate the null value. These analyses used summary statistics from BMI-unadjusted sleep GWASs. IVs were selected based on APM estimates ($\hat{\gamma}_{APM}$). The columns represent the exposure variables, while the rows show the outcome variables. Due to the large standard errors of the $\hat{\beta}_{F,meta}$ estimates (estimates that used $\hat{\gamma}_{meta}$), the corresponding CIs were truncated. Panel c compares estimated $\beta_F$ (x-axis) and $\beta_M$ (y-axis) obtained when using APM estimates ($\hat{\gamma}_{APM}$). The color of each point in panel c is proportional to the p-value of the sex differences test.

Abbreviations: CI: confidence interval; IV: instrumental variable; OR: odds ratio; MR: Mendelian randomization; MR-RAPS: MR using robust adjusted profile score method; APM: adaptive posterior mean; AW: adaptive weight; meta: fixed-effect meta estimates; sex diff p-value: p-value of sex differences test; OSA: obstructive sleep apnea; sleepiness: excessive daytime sleepiness; AF: atrial fibrillation; CAD: coronary artery disease; CKD: chronic kidney disease; HF: heart failure; HTN: hypertension; T2DM: type 2 diabetes mellitus.

#### Supplementary Figure 18: Results from sex-specific causal effect estimation using variants selected by APM estimates (BMI-adjusted sleep GWASs)

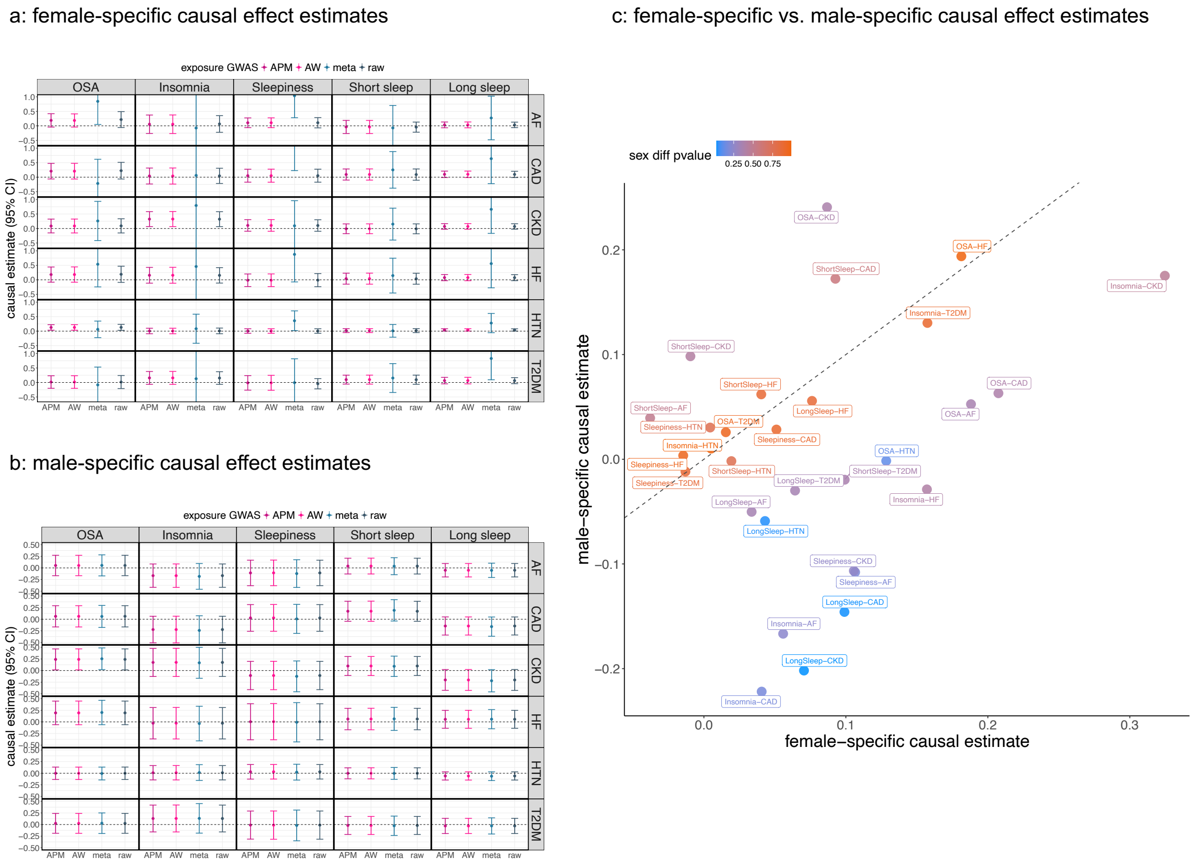

Panels a (female-specific) and b (male-specific) provide sex-specific causal effect ($\beta_F, \beta_M$) estimates with the corresponding 95% CIs. The causal effects were estimated using MR-RAPS and are displayed on the log(OR) scale. Horizontal dashed lines indicate the null value. These analyses used summary statistics from BMI-adjusted sleep GWASs. IVs were selected based on APM estimates ($\hat{\gamma}_{APM}$). The columns represent the exposure variables, while the rows show the outcome variables. Due to the large standard errors of the $\hat{\beta}_{F,meta}$ estimates (estimates that used $\hat{\gamma}_{meta}$), the corresponding CIs were truncated. Panel c compares estimated $\beta_F$ (x-axis) and $\beta_M$ (y-axis) obtained when using APM estimates ($\hat{\gamma}_{APM}$). The color of each point in panel c is proportional to the p-value of the sex differences test.

Abbreviations: CI: confidence interval; IV: instrumental variable; OR: odds ratio; MR: Mendelian randomization; MR-RAPS: MR using robust adjusted profile score method; APM: adaptive posterior mean; AW: adaptive weight; meta: fixed-effect meta estimates; sex diff p-value: p-value of sex differences test; OSA: obstructive sleep apnea; sleepiness: excessive daytime sleepiness; AF: atrial fibrillation; CAD: coronary artery disease; CKD: chronic kidney disease; HF: heart failure; HTN: hypertension; T2DM: type 2 diabetes mellitus.

#### Supplementary Figure 19: Comparison of causal effect estimates from MR-RAPS and MR-PRESSO using IVs selected by raw variant-exposure effect estimates

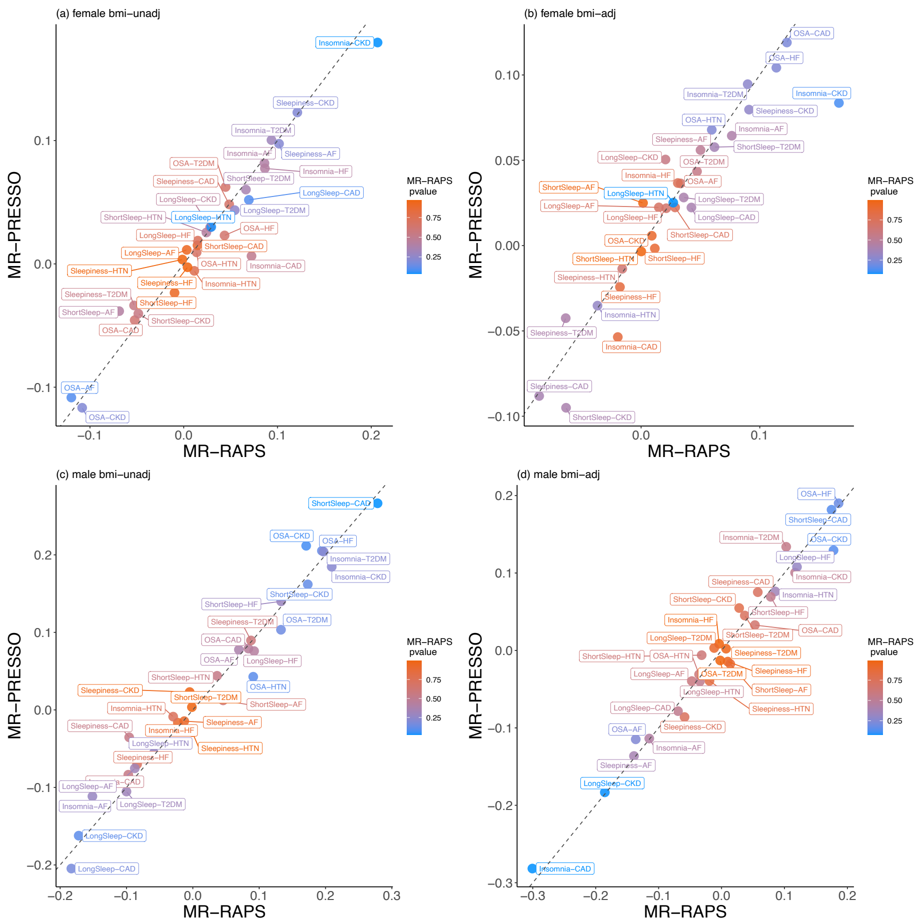

The figure compares of exposure-outcome causal effect ($\beta_F, \beta_M$) estimates between MR-RAPS (x-axis) and MR-PRESSO (y-axis). The IVs were selected based on raw sex-specific variant-exposure estimates ($\hat{\gamma}_{raw}$). The color of each point is proportional to the p-value of the causal effect estimate from MR-RAPS. Panels (a) and (b) show the female-specific results using summary statistics from MVP sleep GWASs without adjustment (panel a) and with adjustment for BMI (panel b), while panels (c) and (d) present the male-specific results without (panel c) and with (panel d) BMI adjustment.

Abbreviation: MR: Mendelian randomization; MR-RAPS: MR using robust adjusted profile score method; MR-PRESSO: Mendelian randomization pleiotropy residual sum and outlier; IV: instrumental variable; OR: odds ratio; MVP: Million Veteran Program; GWAS: Genome-wide association study; bmi-unadj: BMI unadjusted; bmi-adj: BMI adjusted; OSA: obstructive sleep apnea; sleepiness: excessive daytime sleepiness; AF: atrial fibrillation; CAD: coronary artery disease; CKD: chronic kidney disease; HF: heart failure; HTN: hypertension; T2DM: type 2 diabetes mellitus.

#### Supplementary Figure 20: Comparison of causal effect estimates from MR-RAPS and MR-PRESSO when using IVs selected by APM estimates

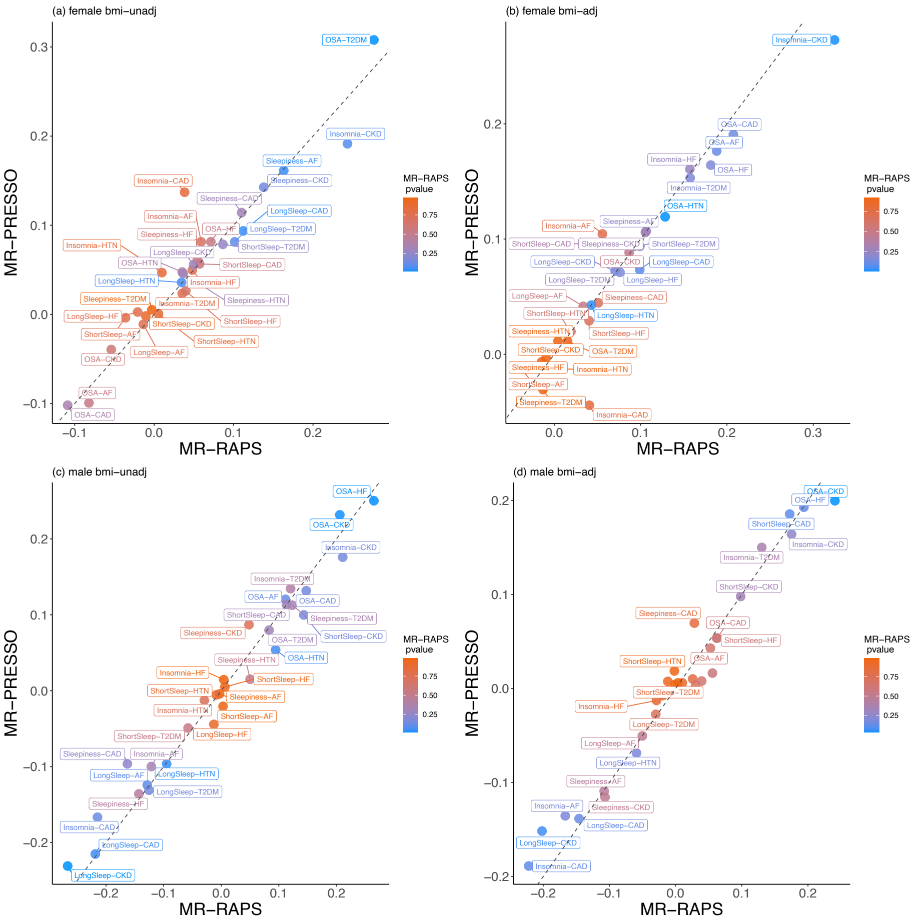

The figure compares exposure-outcome causal effect ($\beta_F, \beta_M$) estimates between MR-RAPS (x-axis) and MR-PRESSO (y-axis). The IVs were selected based on sex-specific APM estimates ($\hat{\gamma}_{APM}$). Estimated effects are displayed on the log(OR) scale. The color of each point is proportional to the p-value of the causal effect estimate from MR-RAPS. Panels (a) and (b) show the female-specific results using summary statistics from MVP sleep GWASs without adjustment (panel a) and with adjustment for BMI (panel b), while panels (c) and (d) present the male-specific results without (panel c) and with (panel d) BMI adjustment.

Abbreviations: MR: Mendelian randomization; MR-RAPS: MR using robust adjusted profile score method; MR-PRESSO: Mendelian randomization pleiotropy residual sum and outlier; IV: instrumental variable; APM: adaptive posterior mean; OR: odds ratio; MVP: Million Veteran Program; GWAS: Genome-wide association study; bmi-unadj: BMI unadjusted; bmi-adj: BMI adjusted; OSA: obstructive sleep apnea; sleepiness: excessive daytime sleepiness; AF: atrial fibrillation; CAD: coronary artery disease; CKD: chronic kidney disease; HF: heart failure; HTN: hypertension; T2DM: type 2 diabetes mellitus.

#### Supplementary 21: Comparison of causal effect estimates using raw and APM-derived IV selection strategies

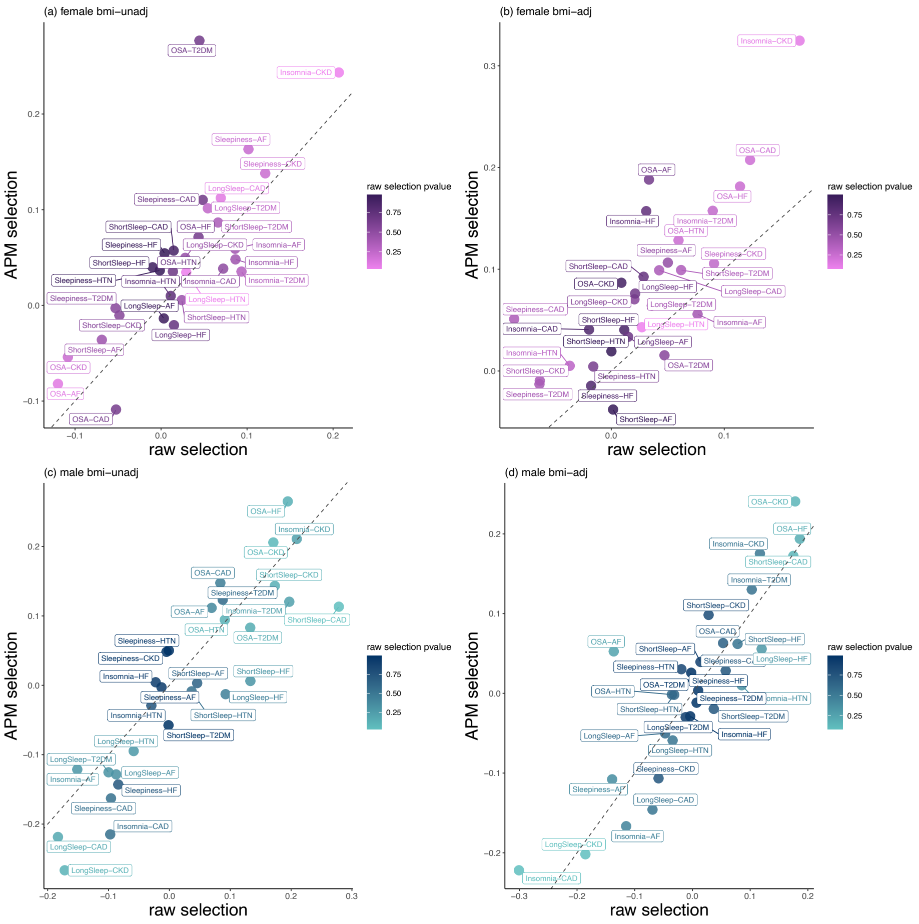

The figure compares exposure-outcome causal effect ($\beta_F, \beta_M$) estimates from using (i) raw estimate ($\hat{\gamma}_{raw}$) and (ii) APM ($\hat{\gamma}_{APM}$) estimate for IV selection. Causal effects were estimated using MR-RAPS with $\hat{\gamma}_{APM}$ and are displayed on a log(OR) scale. The x-axis shows causal effect estimates based on IVs selected using $\hat{\gamma}_{raw}$, while y-axis shows the results for IV selected by $\hat{\gamma}_{APM}$ estimates. Panels (a) and (b) show the female-specific results. Panel (a) used the summary statistics from BMI-unadjusted sleep GWASs, while panel (b) used the summary statistics from BMI-adjusted sleep GWASs. Panels (c) and (d) display the male-specific results, with panel (c) using the summary statistics from BMI-unadjusted sleep GWASs and panel (d) using the summary statistics from BMI-adjusted sleep GWASs. The color of each point is proportional to the p-value of the causal effect estimate from MR-RAPS in primary analysis.

Abbreviations: APM: adaptive posterior mean; IV: instrumental variable; MR-RAPS: MR using robust adjusted profile score method; OR: odds ratio; GWAS: Genome-wide association study; bmi-unadj: BMI unadjusted; bmi-adj: BMI adjusted; OSA: obstructive sleep apnea; sleepiness: excessive daytime sleepiness; AF: atrial fibrillation; CAD: coronary artery disease; CKD: chronic kidney disease; HF: heart failure; HTN: hypertension; T2DM: type 2 diabetes mellitus.

#### Supplementary Figure 22: Results from sex-combined causal effect estimation using variants selected by fixed-effect meta estimates

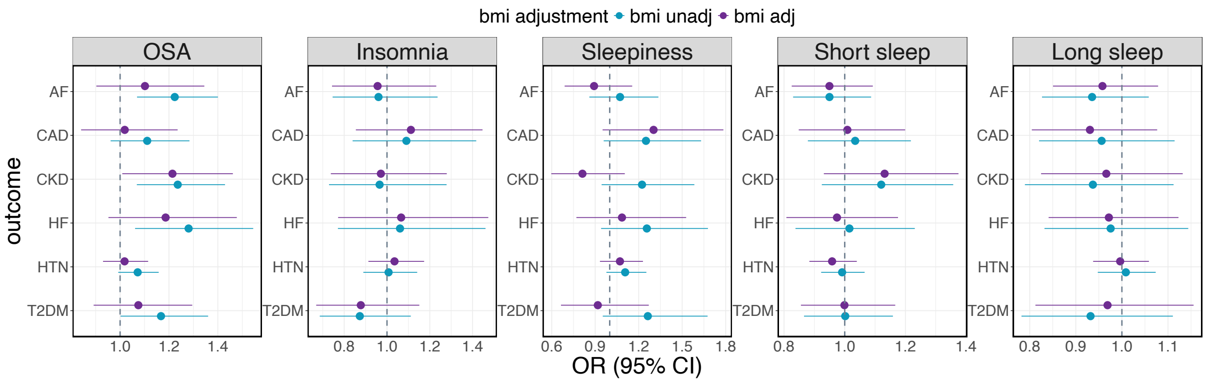

This figure presents sex-combined causal effect estimates with corresponding 95% CIs, based on IVs selected using $\hat{\gamma}_{meta}$. Variant-outcome association effect sizes were computed from a sex-combined dataset in the AoU study. The estimated causal effects, obtained via MR-RAPS using $\hat{\gamma}_{meta}$ variant-exposure effect estimates, are displayed on an OR scale. In each figure, variant-phenotype estimates without BMI adjustment are shown in blue, while those with BMI adjustment are shown in purple. Vertical dashed lines represent the null causal effect. Exposure variables are labeled at the top of each figure, with outcome variables displayed across the rows.

Abbreviations: CI: confidence interval; IV: instrumental variable; AoU: All of Us; MR-RAPS: MR using robust adjusted profile score method; MR: Mendelian randomization; OR: odds ratio; BMI: body mass index; bmi unadj: BMI-unadjusted; bmi adj: BMI-adjusted; OSA: obstructive sleep apnea; AF: atrial fibrillation; CAD: coronary artery disease; CKD: chronic kidney disease; HF: heart failure; HTN: hypertension; T2DM: type 2 diabetes mellitus.

#### Supplementary Figure 23: Miami plot of sex-specific OSA GWAS without BMI adjustment

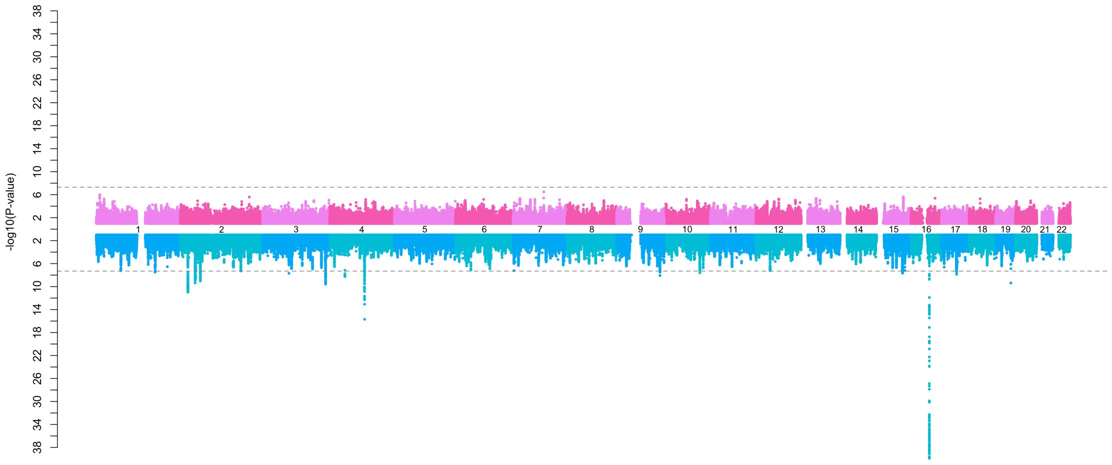

Female-specific results are shown at the top of the plot (pink color), and male-specific results are shown at the bottom (blue color). The y-axis provides the negative log p-values. The horizontal dashed line represents the genome-wide significance threshold ($5 \times 10^{-8}$).
Abbreviations: GWAS: genome-wide association study; BMI: body mass index.

#### Supplementary Figure 24: Miami plot of sex-specific OSA GWAS with BMI adjustment

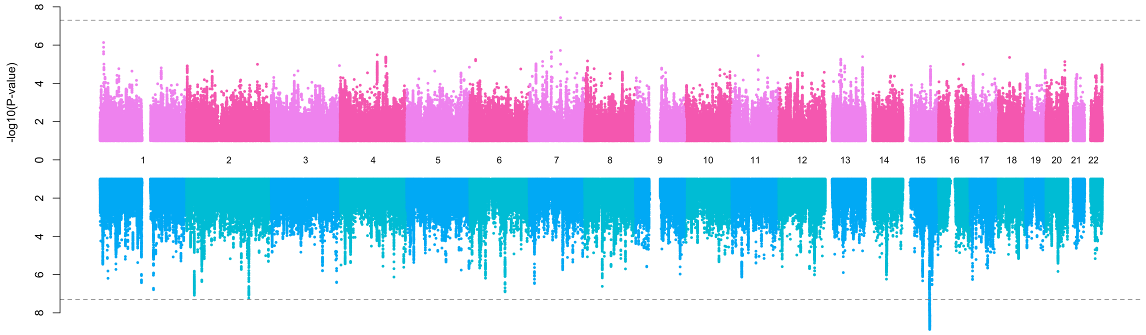

Female-specific results are shown at the top of the plot (pink color), and male-specific results are shown at the bottom (blue color). The y-axis provides the negative log p-values. The horizontal dashed line represents the genome-wide significance threshold ($5 \times 10^{-8}$).
Abbreviations: GWAS: genome-wide association study; BMI: body mass index.

#### Supplementary Figure 25: Miami plot of sex-specific insomnia GWAS without BMI adjustment

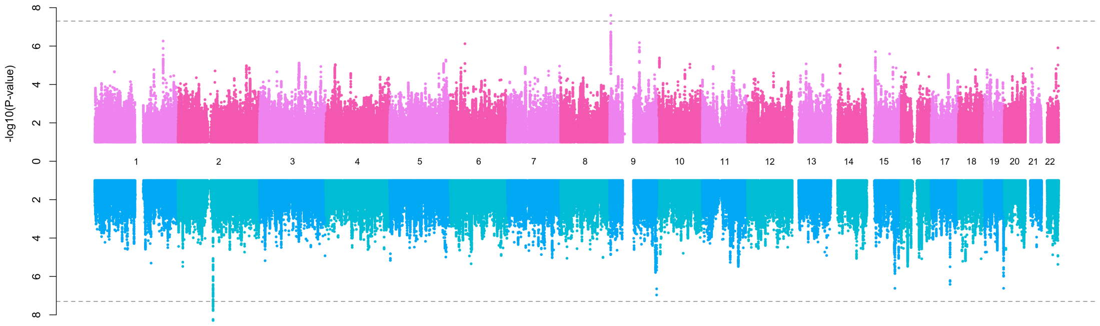

Female-specific results are shown at the top of the plot (pink color), and male-specific results are shown at the bottom (blue color). The y-axis provides the negative log p-values. The horizontal dashed line represents the genome-wide significance threshold ($5 \times 10^{-8}$).
Abbreviations: GWAS: genome-wide association study; BMI: body mass index.

#### Supplementary Figure 26: Miami plot of sex-specific insomnia GWAS with BMI adjustment

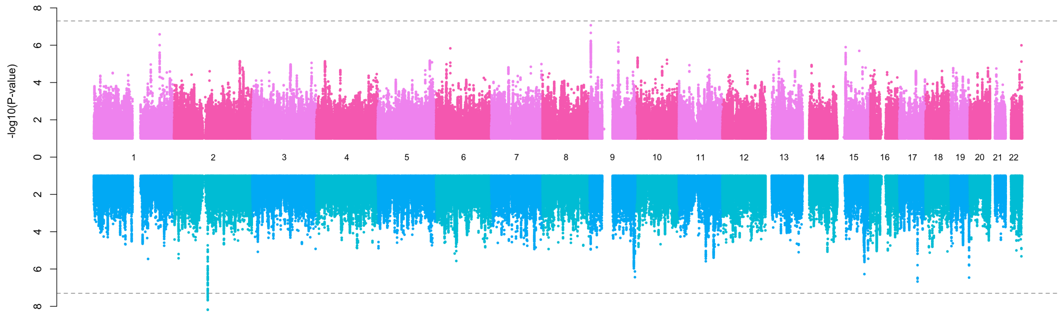

Female-specific results are shown at the top of the plot (pink color), and male-specific results are shown at the bottom (blue color). The y-axis provides the negative log p-values. The horizontal dashed line represents the genome-wide significance threshold ($5 \times 10^{-8}$).
Abbreviations: GWAS: genome-wide association study; BMI: body mass index.

#### Supplementary Figure 27: Miami plot of sex-specific GWAS of excessive daytime sleepiness GWAS without BMI adjustment

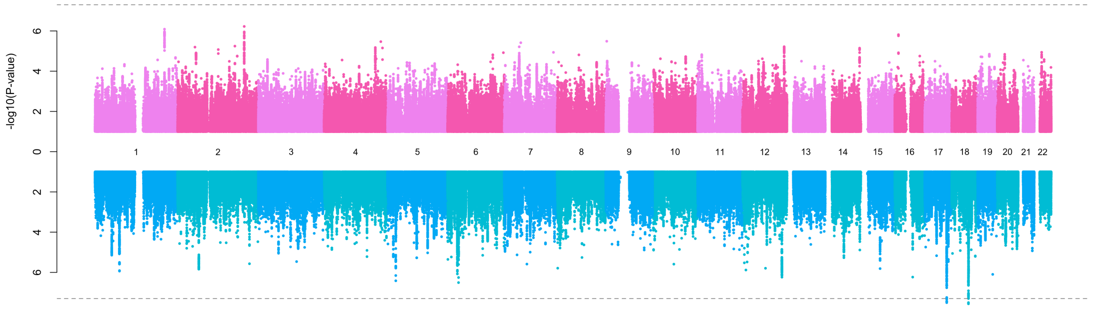

Female-specific results are shown at the top of the plot (pink color), and male-specific results are shown at the bottom (blue color). The y-axis provides the negative log p-values. The horizontal dashed line represents the genome-wide significance threshold ($5 \times 10^{-8}$).
Abbreviations: GWAS: genome-wide association study; BMI: body mass index.

#### Supplementary Figure 28: Miami plot of sex-specific GWAS of excessive daytime sleepiness GWAS with BMI adjustment

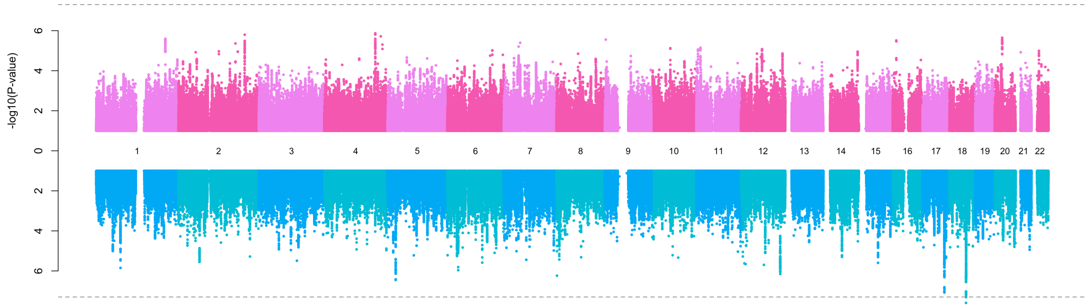

Female-specific results are shown at the top of the plot (pink color), and male-specific results are shown at the bottom (blue color). The y-axis provides the negative log p-values. The horizontal dashed line represents the genome-wide significance threshold ($5 \times 10^{-8}$).
Abbreviations: GWAS: genome-wide association study; BMI: body mass index.

#### Supplementary Figure 29: Miami plot of sex-specific GWAS of short sleep duration, without BMI adjustment

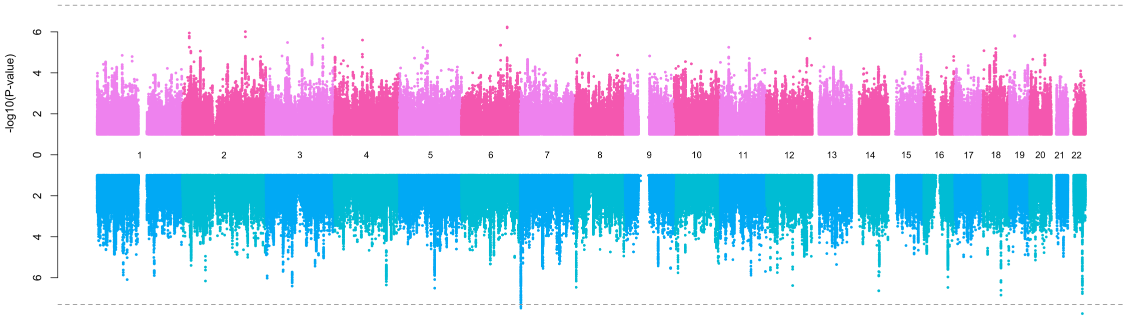

Female-specific results are shown at the top of the plot (pink color), and male-specific results are shown at the bottom (blue color). The y-axis provides the negative log p-values. The horizontal dashed line represents the genome-wide significance threshold ($5 \times 10^{-8}$).
Abbreviations: GWAS: genome-wide association study; BMI: body mass index.

#### Supplementary Figure 30: Miami plot of sex-specific GWAS of short sleep duration GWAS, BMI adjusted

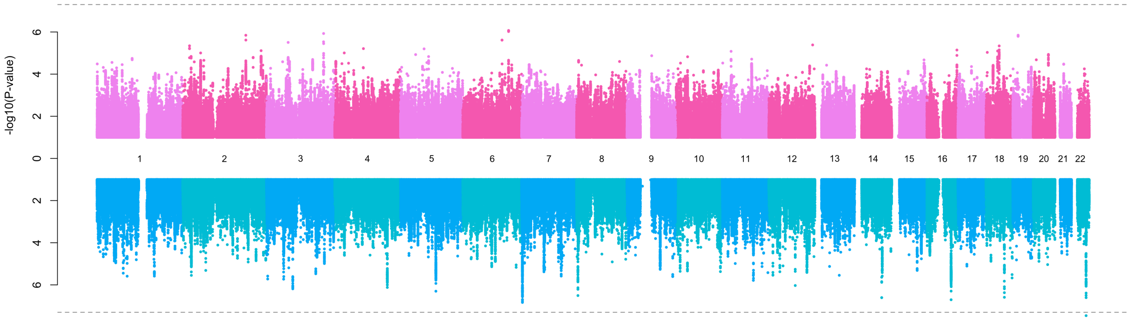

Female-specific results are shown at the top of the plot (pink color), and male-specific results are shown at the bottom (blue color). The y-axis provides the negative log p-values. The horizontal dashed line represents the genome-wide significance threshold ($5 \times 10^{-8}$).
Abbreviations: GWAS: genome-wide association study; BMI: body mass index.

#### Supplementary Figure 31: Miami plot of sex-specific GWAS of long sleep duration, unadjusted to BMI

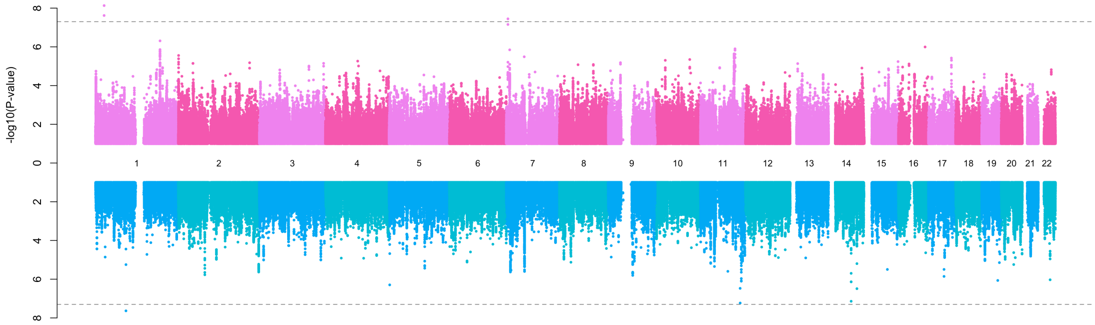

Female-specific results are shown at the top of the plot (pink color), and male-specific results are shown at the bottom (blue color). The y-axis provides the negative log p-values. The horizontal dashed line represents the genome-wide significance threshold ($5 \times 10^{-8}$).
Abbreviations: GWAS: genome-wide association study; BMI: body mass index.

#### Supplementary Figure 32: Miami plot of sex-specific GWAS of long sleep duration, BMI adjusted

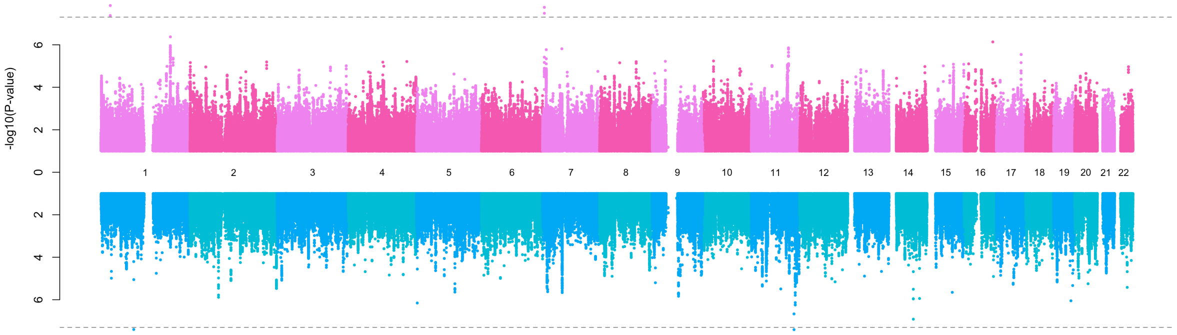

Female-specific results are shown at the top of the plot (pink color), and male-specific results are shown at the bottom (blue color). The y-axis provides the negative log p-values. The horizontal dashed line represents the genome-wide significance threshold ($5 \times 10^{-8}$).
Abbreviations: GWAS: genome-wide association study; BMI: body mass index.
